# Health and budget impact, and price threshold for cost-effectiveness of lenacapavir for PrEP in Eastern and Southern Africa: a modeling analysis

**DOI:** 10.1101/2024.08.20.24312137

**Authors:** Linxuan Wu, David Kaftan, Rachel Wittenauer, Cory Arrouzet, Nishali Patel, Arden L. Saravis, Brian Pfau, Edinah Mudimu, Anna Bershteyn, Monisha Sharma

**Author notes:** Corresponding author: Monisha Sharma, University of Washington, Box 359931, Seattle, WA 98195, United States. Co-last authors (Anna Bershteyn, Monisha Sharma).

## Abstract

**Background:** Six-monthly injectable lenacapavir is a promising product for HIV pre-exposure prophylaxis (PrEP). However, health and budget impacts and threshold price at which lenacapavir could be cost-effective in Eastern and Southern Africa is unknown.

**Methods:** We adapted an agent-based model, EMOD-HIV, to simulate lenacapavir scale-up in Zimbabwe, South Africa, and western Kenya from 2026-2036; uptake assumptions were informed by a literature review of PrEP product preferences. In the main analysis, we varied lenacapavir coverage by subgroup: female sex workers (32%), male clients of FSWs (30%), adolescent girls/young women >1 partner (32%), older females with >1 partner (36%), and males >1 partner (32%). We also assessed a higher coverage scenario (64-76% across subgroups) and scenarios of expanding lenacapavir use, varying from concentrated among those with highest HIV risk to broader coverage. We estimated maximum per-dose lenacapavir price that achieved cost-effectiveness (<US$500/disability-adjusted life-year averted) and 5-year budget impact, compared to daily oral PrEP only.

**Findings:** In the main analysis, lenacapavir was projected to achieve 1.4-3.5% population coverage across settings and avert 12.3-18.0% of infections over 10 years. Maximum per-dose price was highest in South Africa ($106.30), followed by Zimbabwe ($21.10), and lowest in western Kenya ($16.60). The 5-year budget impact (in millions) was US$507.25 in South Africa, US$16.80 in Zimbabwe, and US$4.09 in western Kenya. Lenacapavir provision costs made up >90% of the budget impact. In the higher coverage scenario, lenacapavir distribution reached 2.7-6.9% population coverage and averted 21-33% of HIV infections across setting; price thresholds were 10-18% lower: $88.34 in South Africa, $17.71 in Zimbabwe, and $14.78 in western Kenya. Expanding lenacapavir coverage resulted in higher HIV incidence reductions but lower price thresholds.

**Interpretation:** Lenacapavir can avert substantial HIV incidence; price thresholds and budget impacts vary by setting and coverage. Results can inform policy deliberations regarding lenacapavir pricing and resource planning.

**RESEARCH IN CONTEXT:** *Evidence before this study:* We searched PubMed for modelling studies published through July 31, 2024 that assessed the health or economic impact of long-acting PrEP scale-up in Africa using the terms: “HIV” AND “lenacapavir” OR “pre-exposure prophylaxis” OR “PrEP” AND (a list of terms indicating health impact), “cost*”, “budget impact”, “economic evaluation” and (a list of countries in sub-Saharan Africa), “sub-Saharan” AND “model*”, OR “mathematical model*”. We did not find modeling studies evaluating the maximum cost or budget impact of lenacapavir for PrEP. However, several modelling studies evaluated the cost-effectiveness of other long-acting injectable PrEP products in South Africa. Two studies found that CAB-LA was cost-effective when used by populations at substantial HIV risk, while another found it was not cost-effective when delivered to heterosexual men. Five more recent modelling studies evaluating CAB-LA (four parameterized to South Africa and one to sub-Saharan Africa), found that CAB-LA is not cost-effective when targeted to those at highest risk unless the price is reduced. One compartment model evaluated the price threshold at which CAB-LA would be similarly cost-effective to oral PrEP in South Africa and found a per-dose cost ranging from $9.05-$14.47. We found a lack of studies evaluating LA-PrEP in other African settings aside from South Africa, which has a considerably higher gross domestic product than other countries in the region.

*Added value of this study:* We evaluate the health impact and budget impact and maximum price threshold of lenacapavir in three African settings (South Africa, Zimbabwe, and Kenya) using an individual-based network transmission model. We find that lenacapavir scale up to 1.4-3.5% population coverage across settings can avert 12.3-18.0% of infections and can be cost-effectively implemented at a price per-dose of $106.30 (South Africa), $21.10 (Zimbabwe), and $16.60 (western Kenya) in our main analysis based on PrEP preference data. Price threshold, volume of doses needed, and budget impact, varied by setting and coverage.

*Implications of all the available evidence:* LA PrEP formulations have the potential to substantially reduce HIV burden in low- and middle-income countries, but costs will likely need to be reduced to enable equitable implementation. Our findings can inform price negotiations and public health planning regarding adoption of novel PrEP products.

## INTRODUCTION

HIV incidence remains unacceptably high in Eastern and Southern Africa (ESA) causing substantial morbidity, mortality, and financial strain on health systems(1). Daily oral pre-exposure prophylaxis (PrEP) is effective at preventing HIV acquisition and available in public clinics in ESA, but uptake is far below UNAIDS targets(2,3); additionally, among those initiating PrEP, adherence and persistence are suboptimal(4). Barriers to oral PrEP use include pill burden and stigma associated with taking daily antiretrovirals(3). Studies among key groups and the general population indicate a preference for long-acting (LA) PrEP compared to daily oral pills, with injectables preferred over implants and a strong desire for longer duration products(5). LA PrEP can overcome barriers associated with uptake, adherence, persistence, by providing a more convenient and discreet alternative to daily pill taking and mitigating adherence challenges.

Lenacapavir has emerged as a particularly promising investigational LA product for HIV prevention, with interim phase 3 clinical trial results demonstrating 100% efficacy in cisgender females age 16 to 25 years in Brazil, Peru, South Africa, and the United States(6). Lenacapavir is an antiviral that disrupts the HIV-1 capsid, interfering with several stages of the HIV replication cycle(7–9). Administered through twice yearly subcutaneous injections, lenacapavir has an extended half-life which provides a 6-month duration of HIV protection(9). As a first-in-class antiretroviral, lenacapavir is not expected to have overlapping drug resistance mutations with other antiretrovirals(8–10) and shows a favorable safety profile, suggesting potential for widespread use(7,11–13).

While lenacapavir has the potential to greatly expand PrEP coverage in ESA, its impact is contingent on its affordability, particularly in resource constrained settings which are disproportionately impacted by HIV. Evaluating realistic scenarios of lenacapavir uptake (informed by preference data) is crucial for informing price thresholds that enable cost-effective implementation, estimating product volume needed, and projecting health impact. We sought to estimate the health and economic impact of lenacapavir scale-up in South Africa, western Kenya, and Zimbabwe, countries that were early adopters of oral PrEP, suggesting favorable regulator environments for lenacapavir adoption. As the per-dose price of lenacapavir is still uncertain, we also estimated the maximum price threshold that achieves cost-effectiveness in each setting. Results can inform price negotiations and policy planning regarding lenacapavir implementation in ESA.

## METHODS

### Mathematical model

We augmented a previously developed agent-based model created by Institute for Disease Modeling to simulate lenacapavir scale-up(14–16). EMOD-HIV is an open-source microsimulation model, integrating population demography, HIV disease progression, and heterosexual network-based transmission of HIV, designed to match age- and sex-specific propensities of sexual partnership formation. The model simulates HIV transmission and the impact of HIV treatment and prevention interventions on the epidemic. HIV interventions are incorporated via configurable healthcare modules, including an HIV care continuum with HIV testing, linkage and retention on antiretroviral therapy (ART), and a PrEP continuum with uptake, adherence, persistence, and re-engagement(17). The model tracks health outcomes including HIV infections, HIV-related deaths, and healthcare utilization, enabling calculation of disability adjusted life years (DALYs) and health-related costs.

The model was parameterized using epidemiological data from South Africa (adult HIV prevalence in 2020: 19.1%), western Kenya—Homa Bay, Kisii, Kisumu, Migori, Nyamira, and Siaya counties, (11.3%), and Zimbabwe (11.9%) including age-specific fertility, mortality, voluntary male circumcision coverage, number of persons on oral PrEP, and healthcare-seeking behavior(17–20) . We calibrated the model to primary data from each setting on HIV prevalence by age and sex, number of persons on ART, population size and age/sex structure, and sizes of key populations including female sex workers (FSWs) and male clients of FSWs. The model was calibrated using an optimization algorithm that maximizes the likelihood of matching observed data. We selected 100 good-fitting model parameter sets using roulette resampling in proportion to the goodness-of-fit of each simulation to calibration data. Details on parameterization and calibration data are available in the Supplemental Appendix I (**Tables S1a-S6c, Figures S1a-S2c**).

### Modelled scenarios

In the baseline (no lenacapavir) scenario, we assumed availability of only daily oral PrEP which was utilized by both males and females age 16-49 years with HIV risk indication and scaled up to currently observed levels in each country(21). We assumed oral PrEP decreased HIV acquisition risk by 75% based on clinical trials accounting for average adherence(22); individuals had a persistence of 3 months based on observed continuation in real-world implementation(23–25). In intervention scenarios, lenacapavir was introduced in 2026 and scaled linearly to the target coverage by 2029. We assumed lenacapavir implementation ends in 2035 to evaluate the impact of a ten-year commitment of lenacapavir delivery and utilized a 35-year analytic time horizon (2026 to 2060) to capture long term outcomes. For the main analysis, we estimated realistic lenacapavir uptake using a comprehensive literature review of PrEP preferences in ESA among different population subgroups with HIV risk and an evaluation of healthcare accessibility across groups (see Supplemental Appendix I pages 4-6 for additional details). Based on our literature review, we developed the following lenacapavir coverage estimates across subgroups: FSWs (40%), male clients of FSWs (40%), adolescent girls and young women (AGYW) age 16-24 years with >1 partner (32%), women ≥age 25 years with >1 partner (36%), and men age ≥18 years with >1 partner (32%). We assumed lenacapavir decreased risk of HIV acquisition among both sexes by 95% for 6 months, conservatively lower than interim trial results of 100% (6). Persons who discontinued lenacapavir were eligible to re-initiate if they met eligibility criteria, based on current PrEP guidelines. Leveraging insights from the family planning literature which demonstrates an increase in total contraceptive use with the introduction of new methods, we assumed oral PrEP uptake would remain at currently observed levels and the impact of lenacapavir would be largely additive (26). This assumption aligns with qualitative findings on PrEP preferences which indicates some portion of the population would still choose oral PrEP despite availability of LA products(5).

### Sensitivity analyses

Due to the uncertainty surrounding product volume and populations reached by lenacapavir scale-up, we conducted extensive sensitivity analyses varying lenacapavir coverage. We assessed a higher uptake scenario informed by the upper bound estimates of our PrEP preferences literature review and assumed the following lenacapavir uptake across subgroups: FSWs (72%), male clients of FSWs (72%), AGYW age 16-24 years with >1 partner (76%), women over age 25 years with >1 partner (72%), and men age ≥18 years with >1 partner (64%).

We also assessed a higher oral PrEP scenario in the setting of South Africa in which background daily oral PrEP was scaled up to 3-times currently observed levels by 2026 in both the baseline and intervention scenarios using uptake assumptions from our main analysis.

Further, we conduct a set of sensitivity analyses in which we varied distribution of lenacapavir to females with expanding degrees of HIV risk (with and without coverage among males). To categorize HIV risk in the model, we modified an empirically validated risk scoring tool (Vaginal and Oral Interventions to Control the Epidemic (VOICE) score) developed to identify African females with high likelihood of HIV acquisition based on demographic, behavioral and partnership-level factors(27). We evaluated the following scenarios from highest to lowest HIV risk (with higher VOICE score indicating greater risk): 1) FSWs, 2) VOICE score ≥ 5, defined as 3 of the following 4 factors: age 15-24 years; unmarried, ≥ 1 male sexual partner who has other partners; medium sexual activity category, 3) VOICE score ≥ 3, defined as sexually active with at least 2 of the previously mentioned 4 factors; 4) VOICE score ≥ 1, defined as sexually active with at least 1 of the previously mentioned 4 factors. Each of the above scenarios was evaluated alone and in combination with males who are clients of FSWs and males with >1 partner (See Supplement Appendix I page 52 for details).

### Model outcomes

For each scenario, we estimated the number of HIV infections, HIV-related deaths, disability-adjusted life years (DALYs) and percentage of each outcome averted compared to the counterfactual scenario of oral PrEP only. We also estimated total doses of lenacapavir distributed and doses per HIV infection averted. We calculated 95% credible intervals across 100 good-fitting parameter sets to assess parameter uncertainty. Analysis of model outputs was performed using R version 4.2.2(28).

### Price threshold analysis

We calculated the maximum price per dose for lenacapavir to achieve cost-effectiveness in each scenario using a commonly referenced supply-side cost-effectiveness threshold of $500 USD per DALY averted.(29) Costs (2021 $USD) included HIV testing, ART, HIV-related hospitalizations, and costs related to lenacapavir provision including personnel, consumables, product wastage and demand generation activities (assumed to be 10% of the per dose price) (Table 1; Supplemental Appendix I pages 49-51). We additionally evaluated a lower cost threshold of $200 per DALY averted for all settings and a higher threshold ($1,175 per DALY averted) for South Africa based on country-specific estimates which are higher in South Africa compared to the rest of ESA(29).

**Table 1.** Model parameters, assumptions, and cost inputs^¥^.

| **Model assumptions** | **Value** | **Source** |
| --- | --- | --- |
| Oral PrEP effectiveness | 75% | Baeten 2012 (22) |
| Lenacapavir effectiveness | 95% | Bekker 2024 (37) |
| Lenacapavir scale up period | 2026 – 2029 | Assumption |
| Lenacapavir implementation period | 2026 – 2035 | Assumption |
| Analytic time horizon | 2026 – 2060 | Assumption |
| **Costs (2021 USD)** |  |  |
| Lenacapavir demand generation | 10% per dose | Assumption based on VMMC Torres-Rueda 2018 (38) |
| Lenacapavir provision cost* | \$8.55 | Mangale 2022 (39) 16/08/2024 15:12:00 |
| Lenacapavir wastage | 5% | Assumption |
| **Western Kenya** |  |  |
| **Annual health care costs (among those not on ART)** |  |  |
| HIV-positive CD4 < 200 | \$110.30 | Eaton 2024 (40) |
| HIV-positive CD4 200 - 349 | \$30.38 | Eaton 2024 (40) |
| HIV-positive CD4 > 350 | \$8.59 | Eaton 2024 (40) |
| End of life care | \$105.68 | Eaton 2024 (40) |
| ART provision (annual) | \$196.85 | Larson 2018 (41), Long 2010 (42), The Global Fund (43), and Haas 2015 (44) |
| Oral PrEP per person month | \$10.88 | Wanga 2019 (45) |
| Facility-based HIV-positive test | \$3.68 | Meisner 2021(46) 16/08/2024 15:12:00 |
| Facility-based HIV-negative test | \$2.64 | Meisner 2021(46) |
| **South Africa** |  |  |
| **Annual health care costs (among those not on ART)** |  |  |
| HIV-positive CD4 < 200 | \$374.08 | Eaton 2024 (40) |
| HIV-positive CD4 200 - 349 | \$102.95 | Eaton 2024 (40) |
| HIV-positive CD4 > 350 | \$29.10 | Eaton 2024 (40) |
| End of life care | \$358.10 | Eaton 2024 (40) |
| ART provision (annual) | \$189.56 | Larson 2018 (41), Long 2010 (42), The Global Fund (43), and Haas 2015 (44) |
| Oral PrEP per person month | \$15.20 | (34,47) |
| Facility-based HIV-positive test | \$5.62 | Meyer-Rath 2019 (48) |
| Facility-based HIV-negative test | \$3.62 | Meyer-Rath 2019 (48) |
| **Zimbabwe** |  |  |
| **Annual health care costs (among those not on ART)** |  |  |
| HIV-positive CD4 < 200 | \$93.98 | Eaton 2024 (40) |
| HIV-positive CD4 200 - 349 | \$25.89 | Eaton 2024 (40) |
| HIV-positive CD4 > 350 | \$7.32 | Eaton 2024 (40) |
| End of life care | \$89.83 | Eaton 2024 (40) |
| ART provision (annual) | \$176.01 | Larson 2018 (41), Long 2010 (42), The Global Fund (43), and Haas 2015 (44) |
| Oral PrEP per person month | \$9.25 | Wanga 2019 (45) |
| Facility-based HIV-positive test | \$3.13 | Meisner 2021(46) |
| Facility-based HIV-negative test | \$2.24 | Meisner 2021(46) |

<sup>‡</sup>PrEP: pre-exposure prophylaxis, ART: antiretroviral therapy. VMMC: voluntary medical male circumcision. Costs are adjusted for inflation and GDP per capita ratio where applicable. ART costs are of delivery costs and assume 3% of patients receive 2nd-line ART. Lenacapavir demand generation costs are adapted from the literature for voluntary medical male circumcision demand generation. Additional details on costing methods are available in Supplemental Appendix 1

### Budget impact analysis

Using the per-dose price threshold calculated in each scenario as well as delivery and program costs, we estimated the undiscounted five-year budget impact of lenacapavir scale up in each scenario from the healthcare payer perspective. We disaggregated the annual costs incurred and averted by cost category over the time horizon.

## RESULTS

In the main scenario, lenacapavir scale up was estimated to avert 18.0% of HIV infections in western Kenya, 17.0% in Zimbabwe, and 12.3% in South Africa during 10 years of implementation compared to the reference scenario of oral PrEP only **(Table 2)**. Although the percentage of infections and deaths averted was lowest in South Africa, the absolute number of DALYs averted was greater due to higher HIV prevalence and population size (Supplemental Appendix 2: **Table S3**). Population-level lenacapavir coverage ranged from 1.4% in South Africa, 2.5% in western Kenya and 3.5% in Zimbabwe, with number of doses required over the first 5 years of implementation highest in South Africa (3,942,509), followed by Zimbabwe (501,406), and western Kenya (145,454). Utilizing a threshold of $500 per DALY averted, the maximum per-dose price to achieve cost-effectiveness was $106.28 in South Africa, $21.15 in Zimbabwe, and $16.58 in western Kenya. Under a lower threshold of $200 per DALY averted, per-dose prices decreased to $74.87 (South Africa), $11.68 (Zimbabwe), and $11.32 (western Kenya). Assuming a higher cost threshold ($1,175 per DALY averted for South Africa only) resulted in higher maximum per-dose prices: $177.09 for the baseline coverage and $146.67 for the higher coverage scenario. Assuming higher oral PrEP availability (3-times observed levels), did not impact the maximum price threshold: $108.21 in South Africa vs. $106.28 in the main analysis (Supplemental Appendix: **Table S3**).

**Table 2:**
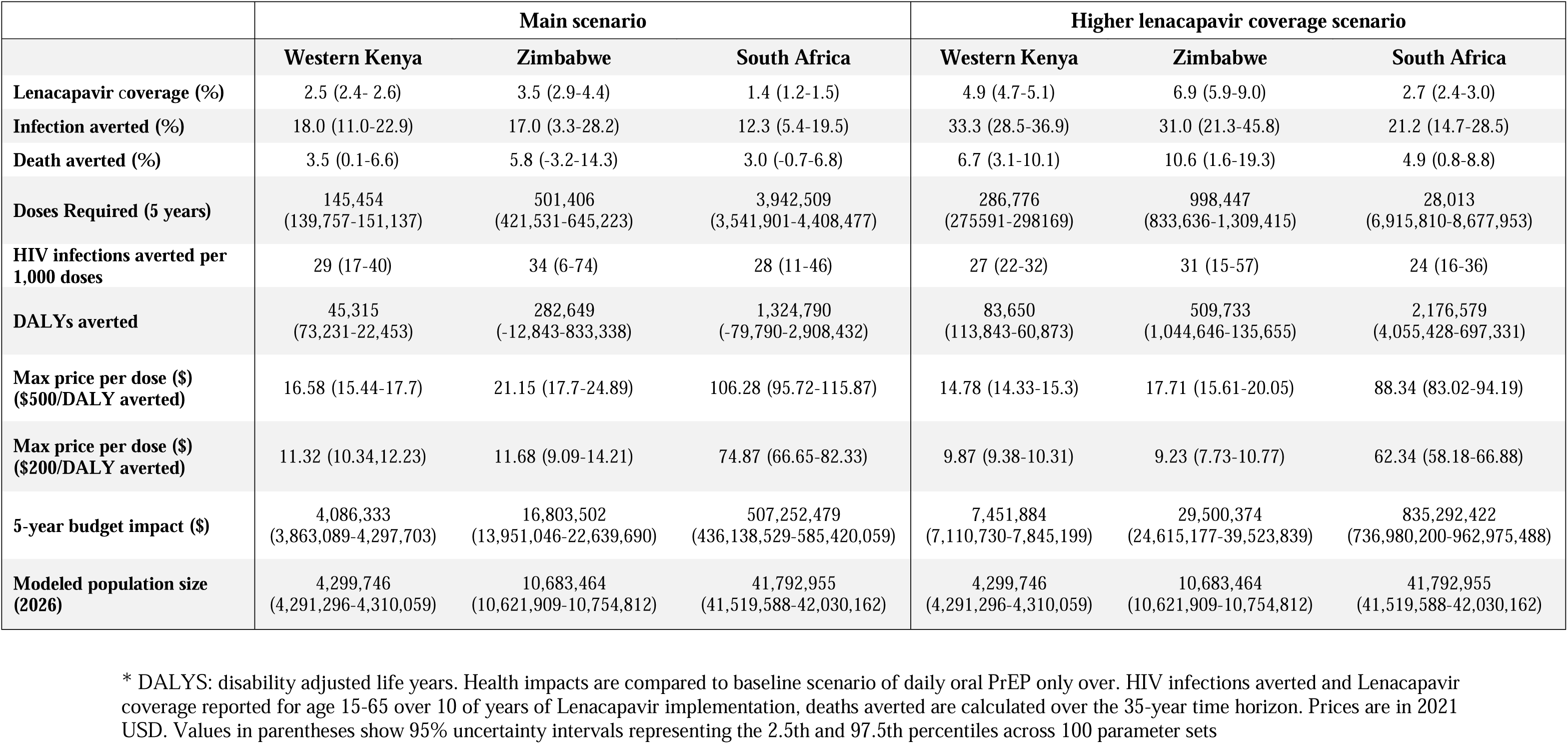
Health and budget impact and maximum price threshold for Lenacapavir scale up*.

Utilizing the per-dose price calculated in the main analysis (based on the $500 threshold), the 5-year budget impact of lenacapavir implementation was $4,086,333 in western Kenya, $16,803,502 in Zimbabwe and $507,252,479 in South Africa (**Table 2**). Costs were highest in South Africa which had the largest modeled population size and highest per-dose price. Costs increased with implementation year as lenacapavir scaled to targeted levels; for example, year one costs in Kenya were $108,500 vs. $1,284,802 in year 5; >90% of the budget impact was due to lenacapavir provision (**Figure 1a and** Appendix II **Table S1**). Cost savings due to HIV-related illness and ART averted increased over time but accounted for <2% of costs across settings. Although lenacapavir provision made up the largest portion of the budget impact, the highest total costs were due to ART provision (approximately 70% of total) (**Figure 1b**).

**Figure 1.**
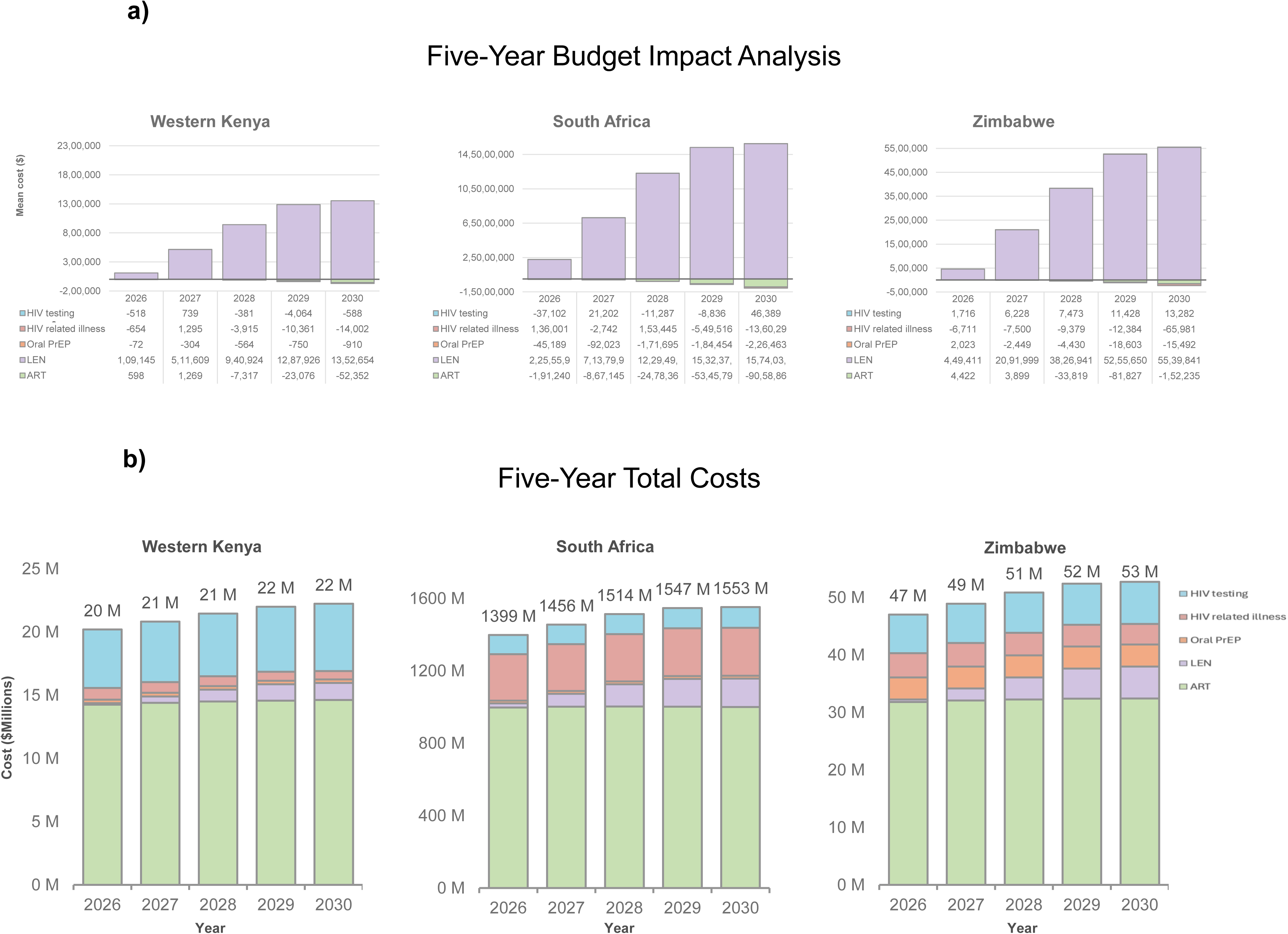

In the higher coverage scenario, lenacapavir distribution reached 2.7-6.9% population coverage across settings and averted 33% of HIV infections in western Kenya, 31% in Zimbabwe, and 21% in South Africa over 10 years (**Table 2**). Both doses required and DALYs averted were nearly double that of the main scenario and the price threshold at $500/DALY averted was 10-18% lower across settings: $88.34 in South Africa, $17.71 in Zimbabwe, and $14.78 in western Kenya. The budget impact followed a similar pattern to the main scenario with lenacapavir provision accounting for most of the costs incurred. Cost savings from HIV-related illness and ART averted were higher but still represented a small proportion of the budget impact (5-8%) (**Supplemental Appendix II Figures S3-5)**.

In sensitivity analyses evaluating lenacapavir distribution to varying subgroups of males and females, price thresholds were considerably higher for scenarios of lenacapavir for FSWs only: $589 in South Africa, $39 in Zimbabwe, and $23; however HIV infections averted were lowest (6-7%) **(Table 3)**. HIV incidence reductions increased with expanding coverage to females with lower VOICE scores but price thresholds decreased. The effect of including males varied by scenario: in scenarios of lenacapavir provision to females with the highest HIV risk (e.g. FSWs and those with VOICE score >5), adding males lowered the price threshold considerably (up to 50%) but price thresholds were similar when adding males to scenarios of females with lower HIV risk (VOICE score>3 and VOICE score >1). In South Africa and Zimbabwe, including males was more efficient (in terms of price threshold and HIV infections averted) than expansion to next-highest-risk group of females across VOICE scores (**Supplemental Appendix II Figures S1-2**). However, in western Kenya, the opposite trend was observed with adding males being less efficient than expansion to the VOICE score across scenarios. Similar patterns were observed when evaluating the impact of adding male clients of FSWs to scenarios of females only instead of males >1 partner (**Supplemental Appendix II Tables S4-15**). Additional health impacts, volume of lenacapavir doses needed, and budget impact for all scenarios are available in Supplemental Appendix II (**Tables S4-15**).

**Table 3:** HIV incidence reduction and maximum price per dose for expanding Lenacapavir distribution scenarios*.

|  | South Africa |  | Western Kenya |  | Zimbabwe |  |
| --- | --- | --- | --- | --- | --- | --- |
|  | Price threshold | HIV incidence reduction (%) | Price threshold | HIV incidence reduction (%) | Price threshold | HIV incidence reduction (%) |
| FSWs |  |  |  |  |  |  |
| FSWs only | \$589.0<br>(454.0 – 726.0) | 6.0<br>(1.0 – 14.0) | \$23.0<br>(18.0 – 28.0) | 7.0<br>(1.7 – 12.8) | \$39.0<br>(28.0 – 54.0) | 5.7<br>(9.2 – 17.2) |
| FSWs and males >1 partner | \$87.0<br>(81.0 – 92.0) | 19.0<br>(13.0 – 25.0) | \$10.2<br>(9.6 – 10.7) | 31.0%<br>(27.4 – 35.0) | \$13.3<br>(13.0 – 15.8) | 21.7<br>(10.1 – 33.8) |
| VOICE Scores |  |  |  |  |  |  |
| Females VOICE > 5 only | \$92.0<br>(84.0 – 100.0) | 16.0<br>(10.0 – 22.0) | \$14.0<br>(13.2 – 7.4) | 35.0<br>(32.0 – 39.0) | \$4.9<br>(2.6 – 7.2) | 13.0<br>(0.2 – 25.4) |
| Females VOICE > 5, and males >1 partner | \$65.0<br>(61.0 – 69.0) | 25.0<br>(20.0 – 31.0) | \$7.5<br>(7.2 – 8.0) | 49.0<br>(49.0 – 52.0) | \$7.2<br>(5.9 – 8.9) | 28.8<br>(18.6 – 40.5) |
| Females VOICE > 3 only | \$43.0<br>(40.0 – 45.0) | 27.0<br>(20.0 – 34.0) | \$8.9<br>(8.5 – 9.3) | 54.4<br>(51.5 – 57.4) | \$6.0<br>(4.7 – 7.2) | 31.0<br>(19.0 – 40.0) |
| Females VOICE > 3, and males >1 partner | \$41.0<br>(39.0 – 42.0) | 35.0<br>(30.0 – 39.0) | \$5.4<br>(5.2 – 5.7) | 63.4<br>(60.9 – 66.0) | \$5.3<br>(4.5 – 6.2) | 40.0<br>(30.0 – 51.0) |
| Females VOICE > 1 only | \$33.0<br>(32.0 – 35.0) | 35.8<br>(30.2 – 41.2) | \$6.4<br>(6.1 – 6.7) | 63.0<br>(60.0 – 66.0) | \$4.3<br>(3.2 – 5.5) | 35.0<br>(26.0 – 44.0) |
| Females VOICE > 1, and males >1 partner | \$32.0<br>(31.0 – 33.0) | 42.5<br>(38.3 – 46.9) | \$4.2<br>(4.0 – 4.4) | 71.0<br>(69.0 – 73.0) | \$4.0<br>(3.3 – 4.9) | 45.0<br>(37.0 – 53.0) |

\* VOICE > 5 is inclusive of FSWs. HIV infections averted are compared to scenario of daily oral PrEP only among persons age 15-65 over 10 of years of Lenacapavir implementation. Prices are in 2021 USD. Values in parentheses show 95% uncertainty intervals representing the 2.5th and 97.5th percentiles across 100 parameter sets.

## DISCUSSION

In this modeling analysis, we evaluated the health and economic impact of lenacapavir scale up in western Kenya, Zimbabwe, and South Africa. We projected that lenacapavir provision at 1.4-3.5% population coverage can substantially decrease HIV incidence and can be cost-effectively implemented at a per-dose price of approximately $17 in western Kenya, $21 in Zimbabwe and $106 in South Africa. Although lenacapavir introduction can result in some cost-savings in the long-term from ART and hospitalizations averted, projected short-term budget impact was substantial, with costs increasing as lenacapavir is scaled up. In an era of shrinking donor funding for HIV programs, policymakers may weigh the impacts of lenacapavir implementation against other non-HIV health interventions which often have a lower cost-effectiveness threshold. Utilizing the threshold of $200/DALY averted reduced the price threshold for lenacapavir to $11 (western Kenya), $12, (Zimbabwe) and $75 (South Africa).

HIV incidence reductions and price thresholds were similar in western Kenya and Zimbabwe; however, the price threshold was 5-times higher in South Africa likely due to higher HIV prevalence and healthcare costs. The volume of lenacapavir doses needed is highest in South Africa (e.g. 7-times higher than Zimbabwe), due to the larger population size, which can inform policy negotiations, as South Africa can present a sizable market for lenacapavir that may sustain initially higher prices while production scales up. Utilizing a higher cost threshold of $1,175 per DALY averted increase the price threshold to $177, slightly higher than the main analysis. These findings also highlight the importance of country-specific analyses as budgetary impact, product volume, and price thresholds vary across settings due to differences in costs and HIV epidemics.

Health impacts and maximum price were sensitive to assumed lenacapavir uptake among subgroups with HIV risk indication. A strength of this analysis is that we conducted a comprehensive literature review of LA PrEP preferences among key groups and the general population in ESA to inform the inputs of our main analyses. In the higher uptake scenario, HIV infections averted increased by 10 percentage points across settings (compared to the main analysis), but per-dose price thresholds were 10-18% lower and doses required doubled. The relationships between health impacts, product volume and price threshold are illustrated by our sensitivity analysis exploring expansion by VOICE score. Lenacapavir provision to FSWs could sustain the highest price per-dose due to high HIV incidence in this group but population-level infections averted were low due to the relatively small number of FSWs. Price thresholds substantially decline with expanding lenacapavir coverage to populations with lower HIV risk and infections averted demonstrated diminishing marginal returns. Additionally, increased demand generation costs may be needed to reach broader coverage levels, which would increase program costs. Interestingly, including males with >1 partner was more efficient than expanding to the next highest risk female category in Zimbabwe and South Africa but not in western Kenya. This may be due to high coverage of voluntary medical male circumcision in western Kenya, which reduces HIV acquisition risk. At coverage of females with VOICE>1 and males with >1 partner, the price threshold decreased to $4 in western Kenya and Zimbabwe and $32 in South Africa; this is consistent with previous analyses that find that broad PrEP distribution in ESA is unlikely to be cost-effective unless PrEP costs are low(30,31). Taken together, these sensitivity analyses can provide insights to policy deliberations regarding the product volume that can be cost-effectively purchased at different price thresholds as countries may be able to commit to obtaining more doses of lenacapavir at lower prices.

To our knowledge this is the first modeling study to investigate the impact of lenacapavir scale up in ESA informed by data on PrEP preferences across subgroups. Our findings are within the range of a previous modeling study of CAB-LA implementation in a general ESA setting which projected that it would be cost-effective at 2.5% coverage at a cost of $57 per 6 months of use, which is lower than our estimate for South Africa but higher than that of Kenya and Zimbabwe (32). All other modeling studies of injectable LA PrEP we found in the published literature evaluated the setting of South Africa and demonstrated that LA PrEP could be cost-effective if provided to FSWs and young women with HIV risk indication (33). One study estimated that the threshold cost of CAB-LA in South Africa was $33-50 for 6 months of use, using the threshold of the same cost-effectiveness as oral PrEP (34). Our results contribute to the literature by estimating the health and economic impact of long-acting PrEP in ESA settings outside South Africa, which differs considerably from other regions in terms of health system costs and HIV epidemic.

We utilized PrEP provision costs from community settings such as pharmacies and assumed that making lenacapavir widely accessible will result in individuals aligning use with HIV risk. This is consistent with prior studies of oral PrEP in ESA which found that making PrEP easily accessible resulted in significant reductions in HIV incidence in the population despite low uptake rates, suggesting that individuals were effectively using PrEP during periods of HIV risk (25,35). We assumed a demand generation cost equivalent to 10% of the estimated per dose price to support educational campaigns to help individuals identify and target PrEP to times of HIV risk. If additional campaigns are needed to increase lenacapavir use among certain subgroups, or if lenacapavir is distributed through higher-resource home or mobile provision these costs may be underestimated. However, certain subgroups such as FSWs could sustain a higher price per-dose, indicating that more intensive outreach may be cost-effective due to higher incidence in this group. Future patterns of oral PrEP use in the context of LA products are uncertain; however our results were robust to scenarios of tripling oral PrEP use. This is likely due to the low levels of current use despite substantial efforts to increase coverage—tripling of low levels still resulted in a very small proportion of oral PrEP coverage.

Our analysis has several limitations. First, as lenacapavir is not yet available in ESA, we relied on stated preference literature to parameterize lenacapavir uptake, which may not align with observed behavior. However, evidence has shown that stated preferences are strongly correlated with actual user choices(36). Additionally, we conducted extensive sensitivity analyses varying lenacapavir uptake across populations with HIV risk indication. Second, we assumed lenacapavir efficacy was 95% in preventing HIV acquisitions among both sexes; however, clinical trial results are not yet available for cisgender men (13). If efficacy is lower than that observed in females, this analysis should be revisited. Third, we assume lenacapavir availability in ESA by 2026; delays in lenacapavir scale-up will likely result in lower price thresholds due to declining HIV incidence. Fourth, our model only simulates heterosexual mixing which accounts for the majority of the HIV epidemic in ESA; therefore, we cannot assess the impact of lenacapavir among men who have sex with men, or persons who inject drugs, who are important target groups for LA PrEP.

Strengths of this analysis include using an individual-based network model and evaluating uncertainty across 100 parameter sets in three ESA settings. We conducted a detailed literature review to inform cost assumptions. By including costs of HIV testing, lenacapavir delivery, demand generation, wastage, and HIV-related healthcare costs, we were able to isolate the product price of lenacapavir to better inform decision-making regarding lenacapavir purchasing and budgeting.

Overall, we found that lenacapavir can avert substantial HIV burden but prices will have to be affordable to ensure equitable and cost-effective distribution. Our findings are timely and can inform policy deliberations regarding price thresholds and product volume in an era of novel LA PrEP products.

## Data Availability

Data used for the modeling can be found in the Supplemental appendix. EMOD-HIV is open-source and publicly available online: www.idmod.org/idmdoc.

## ACKNOWLEDGEMENTS

This study was funded by the Bill and Melinda Gates Foundation. We would like to thank Dan Bridenbecker and Clark Kirkman IV from the Institute for Disease Modeling (Seattle, WA, USA) for providing computational support.

## AUTHOR CONTRIBUTIONS

All authors contributed to conceptualization of the modeling question, provided substantive input into the modelling scenarios and assumptions, interpreted results, and critically reviewed manuscript drafts. LW, DK, CA, NP, MS and AB contributed to model development, and coding. LW, CA, NP, AS and BP conducted analysis of model outputs. MS wrote the first draft of the manuscript. All authors read and approved the final manuscript.

