## appendix 1 for "Health and budget impact, and price threshold for cost-effectiveness of lenacapavir for PrEP in Eastern and Southern Africa: a modeling analysis"

**Accompanying the manuscript:**

### Additional results

#### Table S1: Five-year budget impact analysis - main scenario

| **Costs 2021 USD** | **ART** | **HIV testing** | **Illness** | **Lenacapavir** | **Oral PrEP** | **Total** |
| --- | --- | --- | --- | --- | --- | --- |
| **Kenya** |  |  |  |  |  |  |
| **2026** | 598  (-9,658-10,702) | -518  (-9,620-9,067) | -654  (-17,501-15,977) | 109,145  (104,823-113,221) | -72  (-2,275-2,113) | 108,500  (89,887-128,366) |
| **2027** | 1,269  (-33,174-10,702) | 739  (-19,543-9,067) | 1,295  (-32,465-15,977) | 511,609  (491,538-113,221) | -304  (-3,653-2,113) | 514,609  (463,967-128,366) |
| **2028** | -7,317  (-57,020-34,239) | -381  (-28,510-26,795) | -3,915  (-45,995-48,932) | 940,924  (905,327-976,905) | -564  (-4,377-2,370) | 928,747  (853,022- 992,976) |
| **2029** | -23,076  (-76,911-27,004) | -4,064  (-45,952-30,595) | -10,361  (-63,341-39,034) | 1,287,926  (1,239,114-1,340,491) | -750  (-3,639-2,384) | 1,249,675  (1,173,326-1,348,527) |
| **2030** | -52,352  (-139,170-8,826) | -588  (-31,439-41,121) | -14,002  (-65,397-39,552) | 1,352,654  (1,297,412-1,408,029) | -910  (-3,701-2,147) | 1,284,802  (1,185,136-1,372,277) |
| **Zimbabwe** |  |  |  |  |  |  |
| **2026** | 4,422  (-86,509-97,020) | 1,716  (-60,132-44,224) | -6,711  (-208,357-161,700) | 449,411  (372,081-577,388) | 2,023  (-62,663-78,781) | 450,861  (231,596-644,784) |
| **2027** | 3,899  (-308,356-97,020) | 6,228  (-135,723-44,224) | -7,500  (-362,517-161,700) | 2,091,999  (1,764,809-577,388) | -2,449  (-87,512-78,781) | 2,092,177  (1,574,311-644,784) |
| **2028** | -33,819  (-448,030-380,004) | 7,473  (-134,623-169,348) | -9,379  (-429,594-388,012) | 3,826,941  (3,222,764-4,950,398) | -4,430  (-101,463-95,259) | 3,786,785  (2,972,012-5,087,655) |
| **2029** | -81,827  (-608,861-433,635) | 11,428  (-220,070-204,198) | -12,384  (-570,417-417,110) | 5,255,650  (4,413,523-6,696,249) | -18,603  (-116,073-82,184) | 5,154,264  (4,115,489-7,057,242) |
| **2030** | -152,235  (-806,483-428,085) | 13,282  (-170,265-232,270) | -65,981  (-524,489-389,438) | 5,539,841  (4,679,231-7,141,145) | -15,492  (-99,465-83,356) | 5,319,415  (4,122,008- 7,370,707) |
| **South Africa** |  |  |  |  |  |  |
| **2026** | -191,240  (-3,687,130-3,221,022) | -37,102  (-882,803-855,072) | 136,001  (-4,041,431-5,001,473) | 22,555,962  (19,966,912-25,607,930) | -45,189  (-681,159-487,307) | 22,418,432  (17,244,933- 28,588,396) |
| **2027** | -867,145  (-9,134,705-3,221,022) | 21,202  (-1,354,212-855,072) | -2,742  (-7,328,791-5,001,473) | 71,379,936  (63,819,567-25,607,930) | -92,023  (-726,464-487,307) | 70,439,229  (56,677,761- 28,588,396) |
| **2028** | -2,478,365  (-13,915,291-9,311,474) | -11,287  (-1,630,872-1,542,747) | 153,445  (-8,558,129-8,828,681) | 122,949,797  (110,085,834-139,028,641) | -171,695  (-769,228-681,205) | 120,441,894  (102,823,061- 142,258,318) |
| **2029** | -5,345,799  (-19,354,290-11,086,700) | -8,836  (-1,543,391-1,576,058) | -549,516  (-12,035,809-9,826,002) | 153,237,728  (136,887,260-171,468,172) | -184,454  (-812,938-375,945) | 147,149,124  (126,802,110- 173,014,308) |
| **2030** | -9,058,861  (-24,774,120-9,020,046) | 46,389  (-1,712,121-1,926,140) | -1,360,296  (-15,431,644-10,455,926) | 157,403,032  (140,541,726-175,469,171) | -226,463  (-994,104-495,660) | 146,803,800  (120,657,269-171,380,117) |

#### Table S2: Five-year budget impact analysis - Higher lenacapavir coverage scenario

|  | **ART** | **HIV testing** | **Illness** | **Lenacapavir** | **Oral PrEP** | **Total** |
| --- | --- | --- | --- | --- | --- | --- |
| **Kenya** |  |  |  |  |  |  |
| **2026** | 172  (-9,663-9,414) | -276  (-11,986-9,717) | -1,640  (-17,950-15,960) | 197,904  (189,374-205,684) | -73  (-2,550-2,129) | 196,087  (176,033-212,656) |
| **2027** | -1,554  (-42,861-9,414) | -982  (-24,358-9,717) | -2,426  (-39,460-15,960) | 929,592  (891,481-205,684) | -378  (-3,317-2,129) | 924,252  (860,506- 212,656) |
| **2028** | -15,046  (-53,411-33,669) | -2,663  (-26,820-26,120) | -5,042  (-54,517-44,853) | 1,716,222  (1,645,233-1,783,237) | -801  (-4,101-2,352) | 1,692,670  (1,603,892-1,776,781) |
| **2029** | -46,814  (-99,645-9,614) | -4,644  (-45,410, 33,052) | -17,905  (-73,192-41,374) | 2,358,613  (2,269,345-2,454,948) | -1,876  (-5,306-1,943) | 2,287,374  (2,185,152-2,419,890) |
| **2030** | -101,109  (-157,669,-32,200) | -741  (-40,241-37,220) | -24,998  (-75,970-39,489) | 2,480,094  (2,382,858-2,581,250) | -1,745  (-5,379-1,865) | 2,351,502  (2,219,309-2,498,364) |
| **South Africa** |  |  |  |  |  |  |
| **2026** | 11,887  (-4,541,931-3,624,260) | 20,903  (-1,032,983-843,447) | 22,609  (-5,535,460-5,268,538) | 36,590,444  (31,866,990-41,203,503) | -44,793  (-561,176-658,553) | 36,601,049  (28,679,928-45,113,995) |
| **2027** | -876,205  (-9,028,238-3,624,260) | -84,211  (-1,184,761-843,447) | -337,786  (-7,428,363-5,268,538) | 117,063,847  (104,244,419-41,203,503) | -271,039  (-940,996-658,553) | 115,494,606  (97,929,171-45,113,995) |
| **2028** | -3,853,749  (-14,733,940-8,314,937) | 49,630  (-1,450,532-1,262,742) | -418,045  (-10,551,637-10,125,512) | 201,866,378  (180,817,995-227,079,565) | -414,574  (-1,057,122-292,494) | 197,229,640  (170,752,264- 229,561,525) |
| **2029** | -8,908,284  (-20,532,338-5,515,834) | 80,671  (-1,523,786-1,741,487) | -1,928,912  (-12,555,063-8,067,923) | 253,669,880  (225,891,187-284,381,258) | -524,453  (-1,303,585-193,905) | 242,388,901  (208,384,187- 280,324,980) |
| **2030** | -15,795,203  (-29,862,273-482,180) | 205,673  (-1,759,965-1,883,472) | -2,999,234  (-16,867,474-10,486,753) | 262,730,099  (232,963,491-295,973,542) | -563,109  (-1,160,032-135,606) | 243,578,225  (209,629,502- 283,128,586) |
| **Zimbabwe** |  |  |  |  |  |  |
| **2026** | 10,112  (-74,096-92,977) | 6,147  (-46,418-52,675) | -20,767  (-191,957-140,852) | 784,594  (650,762-1,031,338) | -5,568  (-68,808-70,004) | 774,517  (549,437-1,069,033) |
| **2027** | 24,421  (-210,204-92,977) | 1,028  (-147,777-52,675) | -497  (-325,594-140,852) | 3,653,027  (3,060,826-1,031,338) | -7,473  (-108,179-70,004) | 3,670,506  (2,907,375-1,069,033) |
| **2028** | -33,796  (-419,045-291,639) | -3,515  (-137,648-146,610) | -31,523  (-537,676-467,924) | 6,727,725  (5,601,747-8,839,838) | -26,595  (-119,304-68,221) | 6,632,296  (5,462,924-8,977,177) |
| **2029** | -129,656  (-565,982-238,431) | 14,891  (-193,424-198,210) | -49,393  (-506,257-299,973) | 9,239,225  (7,706,121-12,065,100) | -42,257  (-127,868-39,737) | 9,032,810  (7,485,084-11,876,617) |
| **2030** | -260,280  (-801,502-220,918) | 9,144  (-200,940-207,986) | -76,460  (-680,828-392,322) | 9,764,000  (8,159,272-12,751,418) | -46,159  (-145,942-30,976) | 9,390,244  (7,649,022-12,651,096) |

#### Table S3: Health and budget impact and maximum price threshold for South Africa*

|  | Main Scenario | Higher background oral PrEP scenario |
| --- | --- | --- |
|  | **South Africa** | **South Africa** |
| Lenacapavir coverage (%) | 1.4 (1.2-1.5) | 1.4 (1.2-1.5) |
| Infection averted (%) | 12.3 (5.4-19.5) | 12.5 (5.4-18.5) |
| Death averted (%) | 3.0 (-0.7-6.8) | 3.1 (-1.1-7.1) |
| Doses Required (5 years) | 3,942,509  (3,541,901-4,408,477) | 3,959,450  (3,515,260-4,437,445) |
| HIV infections averted per 1,000 doses | 28 (11-46) | 28 (12-45) |
| DALYs averted | 1,324,790  (-79,790-2,908,432) | 1,312,541  (-226,047-2,868797) |
| Max price per dose ($) ($500/DALY averted) | 106.28 (95.72-115.87) | 108.21 (99.34-117.20) |
| Max price per dose ($) ($200/DALY averted) | 74.87 (66.65- 82.33) | 77.14 (69.98-84.14) |
| 5-year budget impact ($) | 507,252,479  (436,138,529-585,420,059) | 522,058,105  (456,003,817-602,484,388) |
| Modeled population size  (2026) | 41,792,955  (41,519,588-42,030,162) | 41,797,010  (41,525,744-42,048,639) |

* DALYS: disability adjusted life years. Health impacts are compared to baseline scenario of daily oral PrEP only over a 35 year time horizon. Prices are in 2021 USD.

#### Figure S1: Sensitivity analyses: Expanding coverage to females only by VOICE score

Reduction in new HIV infections*

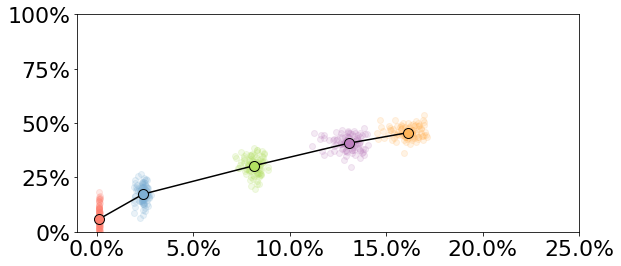

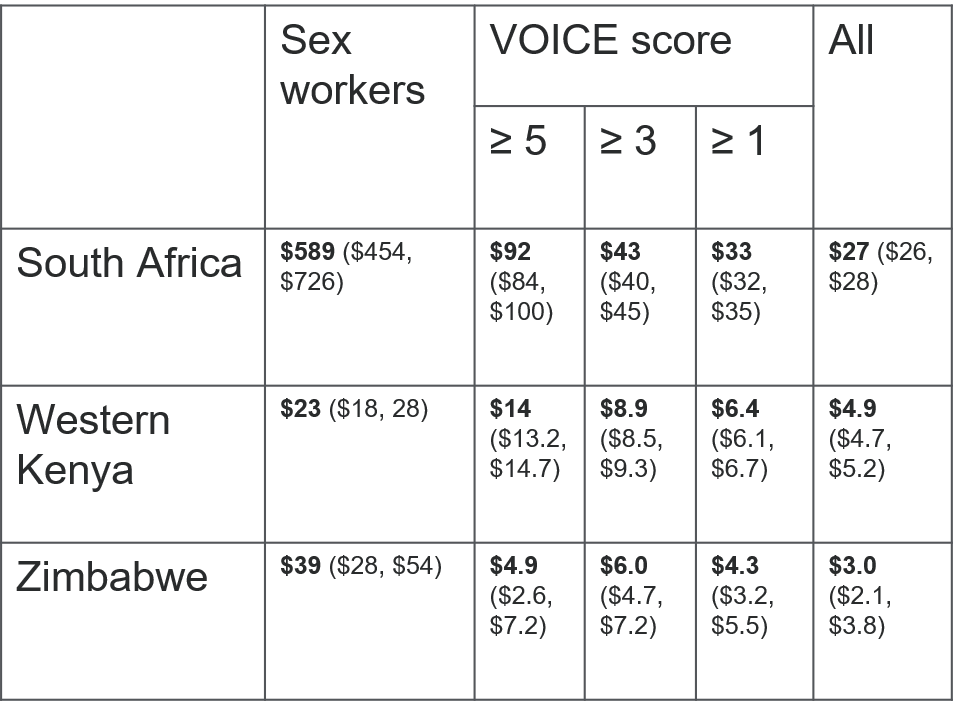

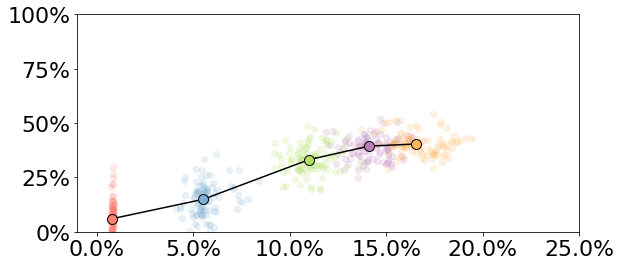

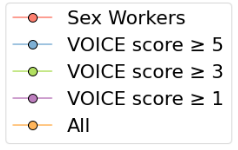

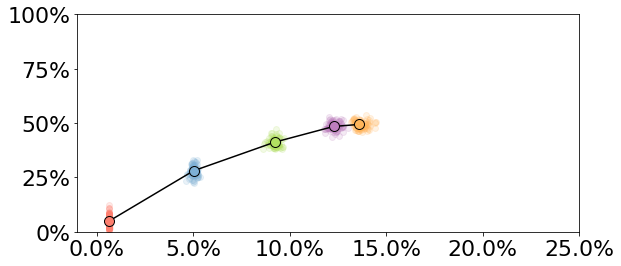

Per-dose price threshold

South Africa

Western Kenya

Zimbabwe

* Reductions in HIV infections over 10 years of Lenacapavir implementation.

** Average % of adults ages 15-49 using Lenacapavir over implementation period.

% of adult population using Lenacapavir*

#### Figure S2: Sensitivity analyses: Expanding lenacapavir coverage to females by VOICE score and males with >1 partner

Per-dose price threshold^¥^

Reduction in new HIV infections*

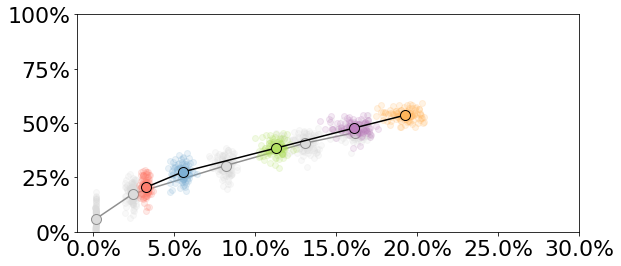

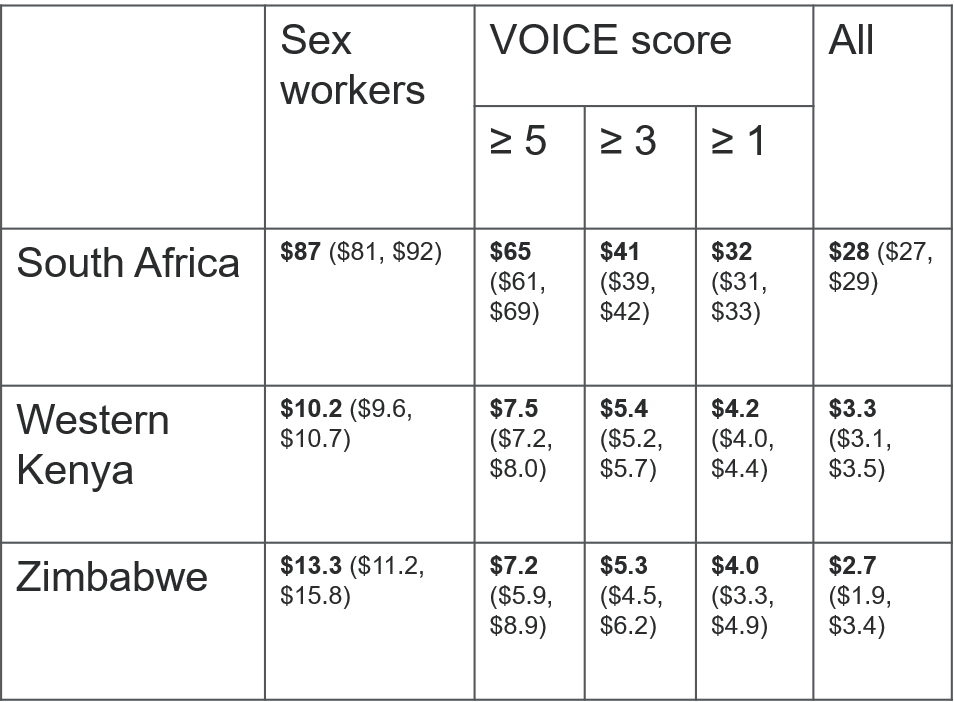

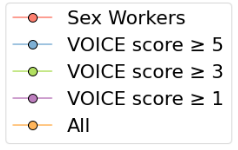

South Africa

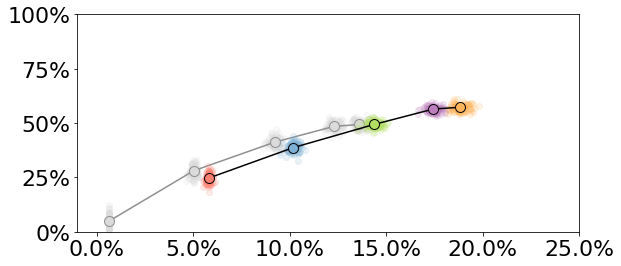

Females only in gray

Western Kenya

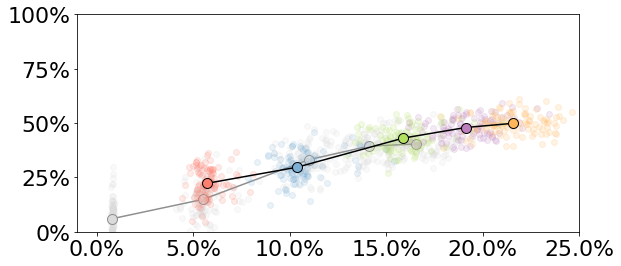

Zimbabwe

* Reductions in HIV infections over 10 years of Lenacapavir implementation.

** Average % of adults ages 15-49 using Lenacapavir over implementation period.

% of adult population using Lenacapavir*

¥ All scenarios include males with >1 partner

#### Western Kenya: lenacapavir distribution to FSWs and males

|  | FSWs | | | FSWs and Male Clients | | | FSW and Males >1 partner | | |
| --- | --- | --- | --- | --- | --- | --- | --- | --- | --- |
|  | **Mean** | **CI Low** | **CI High** | **Mean** | **CI Low** | **CI High** | **Mean** | **CI Low** | **CI High** |
| HIV Infections | 66,044 | 58,830 | 72,472 | 62,915 | 57,010 | 69,133 | 49,011 | 44,446 | 53,597 |
| HIV-related deaths | 66,382 | 60,430 | 71,172 | 65,545 | 60,503 | 70,403 | 61,399 | 57,172 | 66,127 |
| LEN doses (5 yrs) | 51,239 | 49,517 | 52,717 | 224,168 | 220,497 | 228,138 | 477,407 | 463,624 | 488,610 |
| LEN doses (10 yrs) | 116,189 | 113,627 | 118,445 | 511,014 | 504,350 | 518,005 | 1,090,121 | 1,061,123 | 1,118,122 |
| LEN PrEP coverage | 0.006 | 0.006 | 0.007 | 0.028 | 0.028 | 0.029 | 0.060 | 0.058 | 0.062 |
| Infections Averted (n) | 5,045 | 1,125 | 9,284 | 8,175 | 4,514 | 12,047 | 22,078 | 18,456 | 26,733 |
| Infections Averted (prop.) | 0.070 | 0.017 | 0.128 | 0.114 | 0.066 | 0.165 | 0.310 | 0.274 | 0.350 |
| Infections Averted per 1,000 doses | 43.4 | 10.0 | 80.0 | 16.0 | 8.7 | 23.7 | 20.3 | 16.8 | 24.9 |
| Deaths Averted (n) | 1,152 | 900 | 3,205 | 1,989 | 98 | 4,025 | 6,135 | 4,247 | 8,378 |
| Deaths Averted (prop.) | 0.017 | 0.013 | 0.050 | 0.029 | 0.001 | 0.058 | 0.091 | 0.065 | 0.118 |
| DALYs Averted | 20,974 | 5,693 | 49,030 | 35,608 | 11,368 | 61,086 | 103,533 | 78,963 | 127,316 |
| DALYs Averted per 1,000 doses | 181 | 49 | 415 | 70 | 22 | 120 | 95 | 71 | 119 |
| 5 Year Budget Impact (USD) | 103,926,394 | 98,565,584 | 108,118,224 | 105,288,515 | 99,787,498 | 109,539,248 | 109,853,294 | 104,469,256 | 114,145,585 |
| Max price per dose | 23.00 | 18.00 | 28.00 | 6.50 | 5.30 | 7.50 | 10.20 | 9.60 | 10.70 |

|  | Females only VOICE≥5 | | | Females VOICE≥5 & Male Clients of FSWs | | | VOICE≥5 and Males >1 partner | | |
| --- | --- | --- | --- | --- | --- | --- | --- | --- | --- |
|  | **Mean** | **CI Low** | **CI High** | **Mean** | **CI Low** | **CI High** | **Mean** | **CI Low** | **CI High** |
| HIV Infections | 45,998 | 42,509 | 51,014 | 43,851 | 40,620 | 47,904 | 36,581 | 33,495 | 39,982 |
| HIV-related deaths | 62,004 | 56,668 | 66,975 | 61,477 | 56,841 | 65,755 | 59,306 | 54,975 | 63,858 |
| 5-year LEN doses | 407,169 | 390,109 | 424,861 | 580,424 | 563,675 | 597,811 | 833,731 | 807,015 | 863,810 |
| Total LEN doses | 925,062 | 884,674 | 961,994 | 1,321,054 | 1,283,631 | 1,359,420 | 1,901,173 | 1,837,570 | 1,965,977 |
| LEN PrEP coverage | 0.05 | 0.05 | 0.05 | 0.07 | 0.07 | 0.08 | 0.11 | 0.10 | 0.11 |
| Infections Averted (n) | 25,092 | 20,631 | 30,057 | 27,238 | 22,910 | 32,629 | 34,508 | 29,563 | 40,556 |
| Infections Averted (prop.) | 0.35 | 0.32 | 0.39 | 0.38 | 0.35 | 0.42 | 0.49 | 0.46 | 0.52 |
| Infections Averted per 1,000 doses | 27.1 | 22.5 | 32.9 | 20.6 | 17.6 | 24.9 | 18.2 | 15.6 | 21.3 |
| Deaths Averted (n) | 5,530 | 3,617 | 7,854 | 6,057 | 3,562 | 8,384 | 8,228 | 6,163 | 10,554 |
| Deaths Averted (prop.) | 0.08 | 0.06 | 0.11 | 0.09 | 0.05 | 0.12 | 0.12 | 0.09 | 0.15 |
| DALYs Averted | 102,248 | 75,313 | 132,013 | 112,287 | 83,902 | 140,705 | 147,353 | 120,521 | 176,788 |
| DALYs Averted per 1,000 doses | 111 | 83 | 143 | 85 | 62 | 105 | 78 | 65 | 92 |
| 5 Year Budget Impact (USD) | 109,992,628 | 104,424,367 | 114,264,253 | 110,877,051 | 105,385,565 | 115,067,574 | 113,139,381 | 107,603,084 | 117,512,169 |
| Max price per dose | 14.00 | 13.20 | 14.70 | 9.40 | 8.90 | 9.90 | 7.50 | 7.20 | 8.00 |

#### Table S5: Western Kenya: lenacapavir distribution to females VOICE≥5 and males

#### Table S6: Western Kenya: lenacapavir distribution to females VOICE≥3 and males

|  | Females only VOICE≥3 | | | Females VOICE≥3 & Male Clients of FSWs | | | VOICE≥3 and Males >1 partner | | |
| --- | --- | --- | --- | --- | --- | --- | --- | --- | --- |
|  | **Mean** | **CI Low** | **CI High** | **Mean** | **CI Low** | **CI High** | **Mean** | **CI Low** | **CI High** |
| HIV Infections | 32,407 | 28,803 | 35,670 | 30,907 | 28,383 | 34,213 | 25,998 | 23,683 | 28,333 |
| HIV-related deaths | 59,251 | 54,725 | 63,548 | 58,878 | 54,356 | 62,987 | 57,360 | 53,116 | 61,684 |
| LEN doses (5 yrs) | 802,176 | 771,248 | 833,671 | 975,460 | 945,046 | 1,003,873 | 1,228,989 | 1,189,660 | 1,269,295 |
| LEN doses (10 yrs) | 1,759,759 | 1,695,024 | 1,828,006 | 2,155,367 | 2,087,678 | 2,222,918 | 2,736,088 | 2,647,115 | 2,822,821 |
| LEN PrEP coverage | 0.097 | 0.093 | 0.101 | 0.119 | 0.115 | 0.122 | 0.151 | 0.146 | 0.155 |
| Infections Averted (n) | 38,682 | 34,037 | 44,517 | 40,182 | 34,961 | 46,538 | 45,092 | 39,942 | 51,800 |
| Infections Averted (prop.) | 0.544 | 0.515 | 0.574 | 0.565 | 0.534 | 0.591 | 0.634 | 0.609 | 0.660 |
| Infections Averted per 1,000 doses | 22.0 | 18.9 | 25.6 | 18.6 | 16.1 | 21.6 | 16.5 | 14.5 | 18.8 |
| Deaths Averted (n) | 8,283 | 6,351 | 10,514 | 8,656 | 6,426 | 10,842 | 10,175 | 7,842 | 12,810 |
| Deaths Averted (prop.) | 0.122 | 0.097 | 0.152 | 0.128 | 0.100 | 0.155 | 0.150 | 0.120 | 0.180 |
| DALYs Averted | 155,815 | 129,252 | 182,116 | 161,488 | 128,912 | 186,784 | 185,448 | 156,188 | 214,665 |
| DALYs Averted per 1,000 doses | 89 | 75 | 104 | 75 | 59 | 87 | 68 | 57 | 79 |
| 5 Year Budget Impact (USD) | 113,737,660 | 108,129,304 | 117,962,809 | 114,039,696 | 108,424,167 | 118,150,567 | 115,951,299 | 110,360,448 | 120,211,433 |
| Max price per dose | 8.90 | 8.50 | 9.30 | 6.40 | 6.10 | 6.80 | 5.40 | 5.20 | 5.70 |

#### Table S7: Western Kenya: lenacapavir distribution to females VOICE≥1 and males

|  | Females only VOICE≥1 | | | Females VOICE≥1 & Male Clients of FSWs | | | VOICE≥1 and Males >1 partner | | |
| --- | --- | --- | --- | --- | --- | --- | --- | --- | --- |
|  | **Mean** | **CI Low** | **CI High** | **Mean** | **CI Low** | **CI High** | **Mean** | **CI Low** | **CI High** |
| HIV Infections | 26,424 | 24,135 | 29,202 | 24,988 | 22,788 | 27,352 | 20,433 | 18,734 | 22,412 |
| HIV-related deaths | 58,099 | 53,681 | 62,285 | 57,755 | 53,004 | 61,995 | 56,421 | 51,779 | 60,850 |
| LEN doses (5 yrs) | 1,088,735 | 1,056,519 | 1,126,587 | 1,261,919 | 1,228,483 | 1,299,674 | 1,515,401 | 1,469,903 | 1,560,595 |
| LEN doses (10 yrs) | 2,354,608 | 2,286,728 | 2,434,464 | 2,750,316 | 2,678,950 | 2,829,133 | 3,330,383 | 3,233,422 | 3,426,022 |
| LEN PrEP coverage | 0.13 | 0.13 | 0.13 | 0.15 | 0.15 | 0.16 | 0.18 | 0.18 | 0.19 |
| Infections Averted (n) | 44,666 | 39,053 | 51,456 | 46,101 | 41,037 | 52,736 | 50,656 | 44,845 | 57,681 |
| Infections Averted (prop.) | 0.63 | 0.60 | 0.66 | 0.65 | 0.63 | 0.67 | 0.71 | 0.69 | 0.73 |
| Infections Averted per 1,000 doses | 19 | 17 | 22 | 17 | 15 | 19 | 15 | 13 | 17 |
| Deaths Averted (n) | 9,435 | 7,110 | 11,808 | 9,779 | 7,574 | 12,776 | 11,113 | 9,186 | 13,782 |
| Deaths Averted (prop.) | 0.139 | 0.111 | 0.167 | 0.144 | 0.117 | 0.180 | 0.164 | 0.140 | 0.194 |
| DALYs Averted | 178,260 | 147,249 | 211,780 | 184,339 | 154,155 | 219,058 | 205,974 | 176,917 | 238,319 |
| DALYs Averted per 1,000 doses | 76 | 63 | 89 | 67 | 56 | 79 | 62 | 53 | 72 |
| 5 Year Budget Impact | 115,430,512 | 109,722,442 | 119,621,788 | 115,948,751 | 110,400,110 | 120,256,594 | 117,553,634 | 112,023,247 | 121,900,073 |
| Max price per dose | 6.40 | 6.10 | 6.70 | 5.00 | 4.70 | 5.20 | 4.20 | 4.00 | 4.40 |

* Reductions in HIV infections over 10 years of Lenacapavir implementation.

** Average % of adults ages 15-49 using Lenacapavir over implementation period.

% of adult population using Lenacapavir*

#### Table S8: Zimbabwe: lenacapavir distribution to female sex workers (FSWs) and males

|  | FSWs | | | FSWs and Male Clients | | | FSW and Males >1 partner | | |
| --- | --- | --- | --- | --- | --- | --- | --- | --- | --- |
|  | **Mean** | **CI Low** | **CI High** | **Mean** | **CI Low** | **CI High** | **Mean** | **CI Low** | **CI High** |
| HIV Infections | 262,357 | 192,000 | 355,126 | 254,991 | 186,070 | 333,603 | 217,454 | 157,349 | 281,978 |
| HIV-related deaths | 417,600 | 318,258 | 530,666 | 414,596 | 318,519 | 527,299 | 395,882 | 304,211 | 497,142 |
| LEN doses (5 yrs) | 153,780 | 141,632 | 166,463 | 467,175 | 441,311 | 494,855 | 1,056,361 | 905,258 | 1,320,277 |
| LEN doses (10 yrs) | 308,747 | 287,561 | 333,971 | 967,767 | 916,451 | 1,018,532 | 2,171,033 | 1,882,575 | 2,699,276 |
| LEN PrEP coverage | 0.007 | 0.006 | 0.007 | 0.021 | 0.020 | 0.023 | 0.048 | 0.042 | 0.059 |
| Infections Averted (n) | 17,660 | 20,775 | 51,764 | 25,026 | 10,989 | 68,697 | 62,563 | 22,260 | 112,284 |
| Infections Averted (prop.) | 0.057 | 0.092 | 0.172 | 0.082 | 0.050 | 0.203 | 0.217 | 0.101 | 0.338 |
| Infections Averted per 1,000 doses | 57 | 66 | 166 | 26 | 11 | 72 | 29 | 11 | 54 |
| Deaths Averted (n) | 10,579 | 18,232 | 47,968 | 13,583 | 17,954 | 46,882 | 32,297 | 2,023 | 71,764 |
| Deaths Averted (prop.) | 0.023 | 0.045 | 0.097 | 0.029 | 0.041 | 0.092 | 0.074 | 0.006 | 0.147 |
| DALYs Averted | 102,673 | 168,488 | 385,883 | 140,785 | 154,505 | 468,529 | 344,622 | 28,636 | 700,681 |
| DALYs Averted per 1,000 doses | 332 | 523 | 1,324 | 147 | 161 | 484 | 161 | 13 | 336 |
| 5 Year Budget Impact | 265,493,674 | 231,704,027 | 305,201,413 | 264,014,121 | 231,006,775 | 302,750,986 | 274,207,153 | 239,883,402 | 314,675,166 |
| Max price per dose | 39.00 | 28.00 | 54.00 | 13.00 | 8.00 | 17.00 | 13.30 | 11.20 | 15.80 |

#### Table S9: Zimbabwe: lenacapavir distribution to females VOICE≥5 and males

|  | Females only VOICE≥5 | | | Females VOICE≥5 & Male Clients of FSWs | | | VOICE≥5 and Males >1 partner | | |
| --- | --- | --- | --- | --- | --- | --- | --- | --- | --- |
|  | **Mean** | **CI Low** | **CI High** | **Mean** | **CI Low** | **CI High** | **Mean** | **CI Low** | **CI High** |
| HIV Infections | 241,012 | 181,591 | 312,530 | 228,744 | 176,010 | 297,428 | 197,439 | 144,274 | 248,734 |
| HIV-related deaths | 412,439 | 312,932 | 525,592 | 403,634 | 311,063 | 508,611 | 387,274 | 294,833 | 486,801 |
| LEN doses (5 yrs) | 1,032,353 | 819,176 | 1,346,094 | 1,346,429 | 1,135,391 | 1,666,492 | 1,936,905 | 1,630,874 | 2,496,153 |
| LEN doses (10 yrs) | 2,110,752 | 1,686,810 | 2,753,712 | 2,769,453 | 2,318,185 | 3,414,117 | 3,975,712 | 3,364,221 | 5,111,979 |
| LEN PrEP coverage | 0.047 | 0.038 | 0.061 | 0.061 | 0.051 | 0.075 | 0.088 | 0.074 | 0.113 |
| Infections Averted (n) | 39,005 | 354 | 85,738 | 51,273 | 9,229 | 98,448 | 82,578 | 38,845 | 136,333 |
| Infections Averted (prop.) | 0.130 | 0.002 | 0.254 | 0.174 | 0.051 | 0.301 | 0.288 | 0.186 | 0.405 |
| Infections Averted per 1,000 doses | 18.41 | 0.16 | 43.45 | 18.50 | 3.57 | 37.70 | 20.87 | 10.30 | 35.48 |
| Deaths Averted (n) | 15,740 | 18,514 | 50,286 | 24,546 | 16,451 | 63,291 | 40,905 | 3,540 | 94,950 |
| Deaths Averted (prop.) | 0.034 | 0.051 | 0.110 | 0.054 | 0.040 | 0.142 | 0.092 | 0.008 | 0.178 |
| DALYs Averted | 195,866 | 164,748 | 491,487 | 268,334 | 92,308 | 600,970 | 451,940 | 135,063 | 910,543 |
| DALYs Averted per 1,000 doses | 92 | 75 | 241 | 96 | 38 | 213 | 114 | 34 | 242 |
| 5 Year Budget Impact | 268,810,195 | 234,274,604 | 309,398,849 | 269,350,148 | 235,284,302 | 309,675,209 | 278,111,435 | 243,026,859 | 321,762,670 |
| Max price per dose | 4.90 | 2.60 | 7.20 | 5.60 | 4.00 | 7.40 | 7.20 | 5.90 | 8.90 |

#### Table S10: Zimbabwe: lenacapavir distribution to females VOICE≥3 and males

|  | Females only VOICE≥3 | | | Females VOICE≥3 & Male Clients of FSWs | | | VOICE≥3 and Males >1 partner | | |
| --- | --- | --- | --- | --- | --- | --- | --- | --- | --- |
|  | **Mean** | **CI Low** | **CI High** | **Mean** | **CI Low** | **CI High** | **Mean** | **CI Low** | **CI High** |
| HIV Infections | 192,955 | 139,641 | 257,589 | 186,405 | 140,772 | 240,798 | 165,253 | 117,137 | 219,305 |
| HIV-related deaths | 391,670 | 301,665 | 499,886 | 386,987 | 295,225 | 492,585 | 375,152 | 284,038 | 482,836 |
| LEN doses (5 yrs) | 2,014,993 | 1,737,804 | 2,316,826 | 2,330,340 | 2,034,209 | 2,639,880 | 2,919,646 | 2,455,323 | 3,450,648 |
| LEN doses (10 yrs) | 4,138,584 | 3,594,592 | 4,755,734 | 4,804,696 | 4,208,939 | 5,453,506 | 6,008,055 | 5,143,986 | 7,077,561 |
| LEN PrEP coverage | 0.092 | 0.079 | 0.106 | 0.106 | 0.094 | 0.121 | 0.133 | 0.113 | 0.157 |
| Infections Averted (n) | 87,062 | 42,968 | 146,755 | 93,612 | 53,041 | 148,263 | 114,764 | 67,839 | 173,897 |
| Infections Averted (prop.) | 0.31 | 0.19 | 0.40 | 0.33 | 0.24 | 0.42 | 0.40 | 0.30 | 0.51 |
| Infections Averted per 1,000 doses | 21.1 | 10.0 | 35.5 | 19.6 | 10.9 | 30.9 | 19.2 | 11.6 | 29.0 |
| Deaths Averted (n) | 36,509 | 5,377 | 82,574 | 41,192 | 11,269 | 78,879 | 53,027 | 17,392 | 93,377 |
| Deaths Averted (prop.) | 0.08 | 0.01 | 0.17 | 0.09 | 0.03 | 0.16 | 0.12 | 0.04 | 0.19 |
| DALYs Averted | 426,735 | 110,790 | 839,665 | 471,337 | 220,032 | 864,103 | 595,498 | 291,298 | 1,042,021 |
| DALYs Averted per 1,000 doses | 104 | 26 | 206 | 99 | 46 | 179 | 99 | 46 | 162 |
| 5 Year Budget Impact | 279,775,320 | 245,652,932 | 320,010,897 | 278,545,428 | 244,679,099 | 318,107,549 | 284,588,009 | 249,824,317 | 326,438,480 |
| Max price per dose | 6.00 | 4.70 | 7.20 | 5.30 | 4.30 | 6.30 | 5.30 | 4.50 | 6.20 |

#### Table S11: Zimbabwe: lenacapavir distribution to females VOICE≥1 and males

|  | Females only VOICE≥1 | | | Females VOICE≥1 & Male Clients of FSWs | | | VOICE≥1 and Males >1 partner | | |
| --- | --- | --- | --- | --- | --- | --- | --- | --- | --- |
|  | **Mean** | **CI Low** | **CI High** | **Mean** | **CI Low** | **CI High** | **Mean** | **CI Low** | **CI High** |
| HIV Infections | 180,509 | 137,823 | 242,395 | 171,387 | 127,090 | 229,617 | 154,006 | 111,462 | 204,547 |
| HIV-related deaths | 386,757 | 303,656 | 492,854 | 383,617 | 298,730 | 490,751 | 369,923 | 285,743 | 467,507 |
| LEN doses (5 yrs) | 2,620,621 | 2,292,096 | 2,922,073 | 2,937,429 | 2,626,878 | 3,240,703 | 3,528,274 | 3,047,249 | 3,969,289 |
| LEN doses (10 yrs) | 5,329,747 | 4,682,873 | 5,948,679 | 5,990,663 | 5,358,948 | 6,638,811 | 7,199,858 | 6,257,267 | 8,102,494 |
| LEN PrEP coverage | 0.12 | 0.10 | 0.13 | 0.13 | 0.12 | 0.15 | 0.16 | 0.14 | 0.18 |
| Infections Averted (n) | 99,508 | 56,547 | 155,833 | 108,630 | 61,758 | 159,341 | 126,011 | 81,182 | 176,241 |
| Infections Averted (prop.) | 0.35 | 0.26 | 0.44 | 0.38 | 0.28 | 0.48 | 0.45 | 0.37 | 0.53 |
| Infections Averted per 1,000 doses | 18.763 | 10.967 | 30.076 | 18.232 | 9.886 | 28.755 | 17.590 | 10.669 | 27.396 |
| Deaths Averted (n) | 41,422 | 6,435 | 82,421 | 44,562 | 12,759 | 83,036 | 58,256 | 24,537 | 101,067 |
| Deaths Averted (prop.) | 0.09 | 0.02 | 0.18 | 0.10 | 0.04 | 0.17 | 0.13 | 0.06 | 0.21 |
| DALYs Averted | 486,428 | 177,157 | 822,435 | 526,992 | 239,923 | 900,238 | 648,602 | 344,857 | 1,114,948 |
| DALYs Averted per 1,000 doses | 92 | 33 | 156 | 88 | 38 | 154 | 91 | 46 | 153 |
| 5 Year Budget Impact | 282,599,968 | 248,604,820 | 322,059,223 | 281,144,111 | 246,394,335 | 319,726,238 | 286,885,555 | 251,714,351 | 327,393,991 |
| Max price per dose | 4.30 | 3.20 | 5.50 | 3.90 | 3.00 | 4.70 | 4.00 | 3.30 | 4.90 |

#### Table S12: South Africa: lenacapavir distribution to female sex workers (FSWs) and males

|  | FSWs | | | FSWs and Male Clients | | | FSW and Males >1 partner | | |
| --- | --- | --- | --- | --- | --- | --- | --- | --- | --- |
|  | **Mean** | **CI Low** | **CI High** | **Mean** | **CI Low** | **CI High** | **Mean** | **CI Low** | **CI High** |
| HIV Infections | 2,289,509 | 1,983,132 | 2,556,430 | 2,118,620 | 1,880,365 | 2,392,208 | 1,969,688 | 1,721,410 | 2,269,778 |
| HIV-related deaths | 4,248,287 | 3,726,125 | 4,726,742 | 4,178,874 | 3,649,992 | 4,692,426 | 4,122,014 | 3,614,253 | 4,616,165 |
| LEN doses (5 yrs) | 479,916 | 402,361 | 565,211 | 5,872,186 | 5,513,199 | 6,338,869 | 11,163,581 | 10,157,411 | 12,127,878 |
| LEN doses (10 yrs) | 933,625 | 790,955 | 1,072,783 | 11,213,480 | 10,532,624 | 11,975,192 | 21,212,862 | 19,384,506 | 22,975,877 |
| LEN PrEP coverage | 0.001 | 0.001 | 0.001 | 0.014 | 0.013 | 0.015 | 0.026 | 0.024 | 0.028 |
| Infections Averted (n) | 136,637 | 25,386 | 339,713 | 307,526 | 142,327 | 479,928 | 456,458 | 283,293 | 615,728 |
| Infections Averted (prop.) | 0.06 | 0.01 | 0.14 | 0.13 | 0.06 | 0.19 | 0.19 | 0.13 | 0.25 |
| Infections Averted per 1,000 doses | 146.224 | 26.820 | 346.930 | 27.463 | 12.705 | 43.751 | 21.661 | 12.572 | 30.450 |
| Deaths Averted (n) | 57,214 | 104,572 | 214,430 | 126,627 | 16,073 | 276,126 | 183,487 | 53,787 | 307,764 |
| Deaths Averted (prop.) | 0.013 | 0.025 | 0.049 | 0.029 | 0.004 | 0.065 | 0.043 | 0.013 | 0.071 |
| DALYs Averted | 548,139 | 819,950 | 2,001,765 | 1,282,165 | 21,334 | 2,469,982 | 1,939,766 | 510,789 | 3,163,054 |
| DALYs Averted per 1,000 doses | 591.598 | 902.939 | 2,089.348 | 114.119 | 1.921 | 218.595 | 91.840 | 25.634 | 149.937 |
| 5 Year Budget Impact | 7,344,912,783 | 6,648,777,695 | 8,024,850,880 | 7,680,064,982 | 7,026,485,389 | 8,362,511,693 | 7,976,659,644 | 7,337,853,884 | 8,587,317,673 |
| Max price per dose | 589.00 | 454.00 | 726.00 | 112.00 | 102.00 | 123.00 | 87.00 | 81.00 | 92.00 |

#### Table S13: South Africa: lenacapavir distribution to females VOICE≥5 and males

|  | **Females only VOICE≥5** | | | **Females VOICE≥5 & Male Clients of FSWs** | | | **VOICE≥5 and Males >1 partner** | | |
| --- | --- | --- | --- | --- | --- | --- | --- | --- | --- |
|  | **Mean** | **CI Low** | **CI High** | **Mean** | **CI Low** | **CI High** | **Mean** | **CI Low** | **CI High** |
| HIV Infections | 2,047,511 | 1,835,757 | 2,284,787 | 1,934,154 | 1,715,490 | 2,152,971 | 1,826,668 | 1,603,269 | 2,094,663 |
| HIV-related deaths | 4,159,870 | 3,680,208 | 4,645,242 | 4,103,070 | 3,620,870 | 4,627,896 | 4,050,787 | 3,533,069 | 4,533,181 |
| LEN doses (5 yrs) | 8,314,922 | 6,909,900 | 9,320,727 | 13,742,716 | 12,130,735 | 15,036,659 | 19,029,719 | 16,775,709 | 20,819,387 |
| LEN doses (10 yrs) | 15,682,232 | 13,020,805 | 17,477,003 | 26,013,834 | 23,317,847 | 28,207,302 | 36,023,122 | 31,812,452 | 39,054,018 |
| LEN PrEP coverage | 0.019 | 0.016 | 0.022 | 0.032 | 0.029 | 0.035 | 0.044 | 0.039 | 0.048 |
| Infections Averted (n) | 378,635 | 224,297 | 548,156 | 491,992 | 326,064 | 699,832 | 599,478 | 446,637 | 779,684 |
| Infections Averted (prop.) | 0.16 | 0.10 | 0.22 | 0.20 | 0.14 | 0.26 | 0.25 | 0.20 | 0.31 |
| Infections Averted per 1,000 doses | 24.4 | 13.9 | 37.6 | 19.0 | 12.4 | 27.8 | 16.8 | 12.5 | 23.7 |
| Deaths Averted (n) | 145,631 | 4,926 | 271,762 | 202,431 | 60,344 | 353,439 | 254,714 | 109,699 | 399,816 |
| Deaths Averted (prop.) | 0.034 | 0.001 | 0.063 | 0.047 | 0.015 | 0.083 | 0.059 | 0.029 | 0.095 |
| DALYs Averted | 1,483,412 | 218,083 | 2,819,972 | 2,055,762 | 795,644 | 3,266,038 | 2,522,383 | 1,322,796 | 3,911,081 |
| DALYs Averted per 1,000 doses | 96 | 14 | 178 | 80 | 31 | 131 | 70 | 34 | 111 |
| 5 Year Budget Impact | 7,788,872,795 | 7,132,190,126 | 8,433,304,193 | 8,036,996,047 | 7,375,510,581 | 8,682,138,215 | 8,231,620,084 | 7,566,276,893 | 8,895,846,456 |
| Max price per dose | 92.00 | 84.00 | 100.00 | 76.00 | 71.00 | 81.00 | 65.00 | 61.00 | 69.00 |

#### Table S14: South Africa: lenacapavir distribution to females VOICE≥3 and males

|  | Females only VOICE≥3 | | | Females VOICE≥3 & Male Clients of FSWs | | | VOICE≥3 and Males >1 partner | | |
| --- | --- | --- | --- | --- | --- | --- | --- | --- | --- |
|  | **Mean** | **CI Low** | **CI High** | **Mean** | **CI Low** | **CI High** | **Mean** | **CI Low** | **CI High** |
| HIV Infections | 1,765,726 | 1,532,175 | 1,991,446 | 1,678,128 | 1,479,334 | 1,875,318 | 1,584,379 | 1,401,725 | 1,797,341 |
| HIV-related deaths | 4,055,255 | 3,601,778 | 4,476,272 | 4,016,094 | 3,532,696 | 4,496,736 | 3,956,652 | 3,433,242 | 4,398,056 |
| LEN doses (5 yrs) | 28,802,245 | 26,409,891 | 30,559,556 | 34,214,140 | 31,565,877 | 36,198,071 | 39,531,709 | 36,545,543 | 41,789,742 |
| LEN doses (10 yrs) | 53,545,933 | 49,328,410 | 56,819,452 | 63,867,163 | 59,612,147 | 67,705,384 | 73,967,440 | 68,541,172 | 78,260,816 |
| LEN PrEP coverage | 0.066 | 0.061 | 0.070 | 0.079 | 0.073 | 0.083 | 0.091 | 0.085 | 0.097 |
| Infections Averted (n) | 660,420 | 452,387 | 876,139 | 748,018 | 574,292 | 939,616 | 841,766 | 652,972 | 1,066,846 |
| Infections Averted (prop.) | 0.27 | 0.20 | 0.34 | 0.31 | 0.26 | 0.36 | 0.35 | 0.30 | 0.39 |
| Infections Averted per 1,000 doses | 12.4 | 8.5 | 16.6 | 11.7 | 8.8 | 15.3 | 11.4 | 8.9 | 14.6 |
| Deaths Averted (n) | 250,247 | 96,094 | 399,955 | 289,407 | 148,923 | 422,403 | 348,850 | 200,819 | 493,524 |
| Deaths Averted (prop.) | 0.058 | 0.023 | 0.088 | 0.067 | 0.032 | 0.098 | 0.081 | 0.050 | 0.115 |
| DALYs Averted | 2,578,975 | 1,297,933 | 3,777,709 | 2,968,781 | 1,772,623 | 4,213,474 | 3,554,900 | 2,307,312 | 4,913,849 |
| DALYs Averted per 1,000 doses | 48 | 24 | 73 | 47 | 27 | 67 | 48 | 32 | 65 |
| 5 Year Budget Impact | 8,319,881,476 | 7,690,791,407 | 8,979,908,912 | 8,469,077,577 | 7,847,043,197 | 9,145,051,645 | 8,687,490,286 | 8,056,025,146 | 9,348,067,578 |
| Max price per dose | 43.00 | 40.00 | 45.00 | 41.00 | 39.00 | 43.00 | 41.00 | 39.00 | 42.00 |

#### Table S15: South Africa: lenacapavir distribution to females VOICE≥1 and males

|  | Females only VOICE≥1 | | | Females VOICE≥1 & Male Clients of FSWs | | | VOICE≥1 and Males >1 partner | | |
| --- | --- | --- | --- | --- | --- | --- | --- | --- | --- |
|  | **Mean** | **CI Low** | **CI High** | **Mean** | **CI Low** | **CI High** | **Mean** | **CI Low** | **CI High** |
| HIV Infections | 1,553,773 | 1,384,915 | 1,739,361 | 1,459,923 | 1,290,115 | 1,625,904 | 1,393,791 | 1,235,376 | 1,553,787 |
| HIV-related deaths | 3,974,636 | 3,484,547 | 4,453,847 | 3,932,357 | 3,432,637 | 4,430,777 | 3,902,504 | 3,401,788 | 4,443,105 |
| LEN doses (5 yrs) | 46,014,102 | 41,543,544 | 48,925,754 | 51,417,577 | 46,922,192 | 54,458,722 | 56,772,999 | 51,740,681 | 60,220,576 |
| LEN doses (10 yrs) | 85,593,939 | 77,400,981 | 91,040,671 | 95,923,382 | 87,352,828 | 101,451,040 | 106,048,727 | 97,231,010 | 112,214,167 |
| LEN PrEP coverage | 0.105 | 0.095 | 0.112 | 0.118 | 0.108 | 0.126 | 0.130 | 0.119 | 0.138 |
| Infections Averted (n) | 872,373 | 682,130 | 1,080,884 | 966,223 | 763,067 | 1,162,424 | 1,032,355 | 834,267 | 1,232,363 |
| Infections Averted (prop.) | 0.358 | 0.302 | 0.412 | 0.398 | 0.351 | 0.435 | 0.425 | 0.383 | 0.469 |
| Infections Averted per 1,000 doses | 10.2 | 7.7 | 13.1 | 10.1 | 7.9 | 12.6 | 9.8 | 7.8 | 12.1 |
| Deaths Averted (n) | 330,865 | 194,439 | 482,674 | 373,145 | 243,365 | 545,430 | 402,997 | 259,268 | 553,254 |
| Deaths Averted (prop.) | 0.077 | 0.046 | 0.107 | 0.087 | 0.057 | 0.119 | 0.094 | 0.061 | 0.126 |
| DALYs Averted | 3,424,076 | 2,008,845 | 4,624,508 | 3,867,288 | 2,668,754 | 5,400,402 | 4,137,154 | 2,714,364 | 5,423,547 |
| DALYs Averted per 1,000 doses | 40 | 24 | 56 | 40 | 27 | 56 | 39 | 24 | 51 |
| 5 Year Budget Impact | 8,659,482,597 | 8,057,852,317 | 9,264,500,355 | 8,822,827,206 | 8,199,717,334 | 9,432,025,013 | 8,954,499,222 | 8,342,195,004 | 9,548,457,366 |
| Max price per dose | 33.00 | 32.00 | 35.00 | 33.00 | 32.00 | 34.00 | 32.00 | 31.00 | 33.00 |

**Table S16: Summary of HIV incidence reduction and maximum price per dose for expanding Lenacapavir distribution scenarios**

|  | **South Africa** | | **Western Kenya** | | **Zimbabwe** | |
| --- | --- | --- | --- | --- | --- | --- |
|  | Price threshold | HIV incidence reduction (%) | Price threshold | HIV incidence reduction (%) | Price threshold | HIV incidence reduction (%) |
| FSWs | | | | |  |  |
| FSWs only | $589.0  (454.0 – 726.0) | 6.0%  (1.0 – 14.0) | $23.0  (18.0 – 28.0) | 7.0%  (1.7 – 12.8) | $39.0  (28.0 – 54.0) | 5.7  (9.2 – 17.2) |
| FSWs and male clients of FSWs | $112.0  (102.0 – 123.0) | 13.0%  (6.0 – 19.0) | $6.5  (5.3 – 7.5) | 11.4%  (6.6 – 16.5) | $13.0  (8.0 – 17.0) | 8.2  (5.0 – 20.3) |
| FSWs and males >1 partner | $87.0  (81.0 – 92.0) | 19.0%  (13.0 – 25.0) | $10.2  (9.6 – 10.7) | 31.0%  (27.4 – 35.0) | $13.3  (13.0 – 15.8) | 21.7  (10.1 – 33.8) |
| VOICE Scores | | | | | | |
| Females VOICE > 5 only | $92.0  (84.0 – 100.0) | 16.0%  (10.0 – 22.0) | $14.0  (13.2 – 7.4) | 35.0%  (32.0 – 39.0) | $4.9  (2.6 – 7.2) | 13.0  (0.2 – 25.4) |
| Females VOICE > 5, and male clients of FSWs | $76.0  (71.0 – 81.0) | 20.0%  (14.0 – 26.0) | $9.4  (8.9 – 9.9) | 38.0%  (35.0 – 42.0) | $5.6  (4.0 – 7.4) | 17.4  (5.1 – 30.1) |
| Females VOICE > 5, and males >1 partner | $65.0  (61.0 – 69.0) | 25.0%  (20.0 – 31.0) | $7.5  (7.2 – 8.0) | 49.0%  (49.0 – 52.0) | $7.2  (5.9 – 8.9) | 28.8  (18.6 – 40.5) |
| Females VOICE > 3 only | $43.0  (40.0 – 45.0) | 27.0%  (20.0 – 34.0) | $8.9  (8.5 – 9.3) | 54.4%  (51.5 – 57.4) | $6.0  (4.7 – 7.2) | 31.0  (19.0 – 40.0) |
| Females VOICE > 3, and male clients of FSWs | $41.0  (39.0 – 43.0) | 31.0%  (26.0 – 36.0) | $6.4  (6.1 – 6.8) | 56.5%  (53.4 – 59.1) | $5.3  (4.3 – 6.3) | 33.0  (24.0 – 42.0) |
| Females VOICE > 3, and males >1 partner | $41.0  (39.0 – 42.0) | 35.0%  (30.0 – 39.0) | $5.4  (5.2 – 5.7) | 63.4%  (60.9 – 66.0) | $5.3  (4.5 – 6.2) | 40.0  (30.0 – 51.0) |
| Females VOICE > 1 only | $33.0  (32.0 – 35.0) | 35.8%  (30.2 – 41.2) | $6.4  (6.1 – 6.7) | 63.0%  (60.0 – 66.0) | $4.3  (3.2 – 5.5) | 35.0%  (26.0 – 44.0) |
| Females VOICE > 1, and male clients of FSWs | $33.0  (32.0 – 34.0) | 39.8%  (35.1 – 43.5) | $5.0  (4.7 – 5.2) | 65.0%  (63.0 – 67.0) | $3.9  (3.0 – 4.7) | 38.0%  (28.0 – 48.0) |
| Females VOICE > 1, and males >1 partner | $32.0  (31.0 – 33.0) | 42.5%  (38.3 – 46.9) | $4.2  (4.0 – 4.4) | 71.0%  (69.0 – 73.0) | $4.0  (3.3 – 4.9) | 45.0%  (37.0 – 53.0) |

### Figure S1: Five-Year Budget Impact for higher Lenacapavir coverage scenario: Western Kenya

Western Kenya

Mean cost ($)

|  | ART | LEN | Oral PrEP | HIV related illness | HIV testing |
| --- | --- | --- | --- | --- | --- |
| 2026 | 171.818 | 197904 | -72.856 | -1640.05 | -276 |
| 2027 | -1553.79 | 929592 | -377.783 | -2426.04 | -982 |
| 2028 | -15046.3 | 1716222 | -800.702 | -5042.03 | -2663.26 |
| 2029 | -46813.8 | 2358613 | -1876.23 | -17904.8 | -4644.12 |
| 2030 | -101109 | 2480094 | -1744.93 | -24997.7 | -740.85 |

### Figure S2: Five-Year Budget Impact for higher Lenacapavir coverage scenario: South Africa

South Africa

Mean cost ($)

|  | ART | LEN | Oral PrEP | HIV related illness | HIV testing |
| --- | --- | --- | --- | --- | --- |
| 2026 | 11886.9 | 36590444 | -44793.3 | 22608.6 | 20903 |
| 2027 | -876205 | 117063847 | -271039 | -337786 | -84210.6 |
| 2028 | -3853749 | 201866378 | -414574 | -418045 | 49629.7 |
| 2029 | -8908284 | 253669880 | -524453 | -1928912 | 80670.7 |
| 2030 | -15795203 | 262730099 | -563109 | -2999234 | 205673 |

### Figure S3: Five-Year Budget Impact for higher Lenacapavir coverage scenario: Zimbabwe

Mean cost ($)

|  | ART | LEN | Oral PrEP | HIV related illness | HIV testing |
| --- | --- | --- | --- | --- | --- |
| 2026 | 10111.7 | 784594 | -5568.12 | -20766.8 | 6146.86 |
| 2027 | 24421 | 3653027 | -7473.11 | -497.338 | 1028.09 |
| 2028 | -33796.3 | 6727725 | -26595.4 | -31522.5 | -3514.69 |
| 2029 | -129656 | 9239225 | -42257.2 | -49392.5 | 14891.4 |
| 2030 | -260280 | 9764000 | -46159 | -76460.2 | 9143.67 |

### Figure S4: Component Costs by Scenario in western Kenya

Cost ($Millions)

| Year | ART | LEN | Oral PrEP | HIV related illness | HIV testing |
| --- | --- | --- | --- | --- | --- |
| 2026 | 14.2546 M | 0.109145 M | 0.279512 M | 0.926655 M | 4.627 M |
| 2027 | 14.3914 M | 0.511609 M | 0.279546 M | 0.846324 M | 4.79396 M |
| 2028 | 14.4963 M | 0.940924 M | 0.279609 M | 0.77477 M | 4.96849 M |
| 2029 | 14.5708 M | 1.28793 M | 0.279852 M | 0.713262 M | 5.14625 M |
| 2030 | 14.6131 M | 1.35265 M | 0.280002 M | 0.66073 M | 5.32287 M |
| 2026 | 14.2542 M | 0.197904 M | 0.279511 M | 0.925668 M | 4.62724 M |
| 2027 | 14.3886 M | 0.929592 M | 0.279472 M | 0.842603 M | 4.79224 M |
| 2028 | 14.4885 M | 1.71622 M | 0.279372 M | 0.773643 M | 4.96621 M |
| 2029 | 14.547 M | 2.35861 M | 0.278727 M | 0.705718 M | 5.14567 M |
| 2030 | 14.5643 M | 2.48009 M | 0.279167 M | 0.649734 M | 5.32271 M |

### Figure S5: Component Costs by Scenario in western Kenya

Cost ($Millions)

| Year | ART | LEN | Oral PrEP | HIV related illness | HIV testing |
| --- | --- | --- | --- | --- | --- |
| 2026 | 997.856 M | 22.556 M | 15.3229 M | 256.665 M | 106.337 M |
| 2027 | 1001.78 M | 71.3799 M | 15.3639 M | 258.708 M | 108.335 M |
| 2028 | 1003.27 M | 122.95 M | 15.3417 M | 261.816 M | 110.232 M |
| 2029 | 1002.4 M | 153.238 M | 15.4065 M | 264.119 M | 112.179 M |
| 2030 | 999.889 M | 157.403 M | 15.502 M | 265.748 M | 114.039 M |
| 2026 | 998.059 M | 36.5904 M | 15.3233 M | 256.552 M | 106.395 M |
| 2027 | 1001.77 M | 117.064 M | 15.1849 M | 258.373 M | 108.229 M |
| 2028 | 1001.89 M | 201.866 M | 15.0988 M | 261.244 M | 110.293 M |
| 2029 | 998.84 M | 253.67 M | 15.0665 M | 262.74 M | 112.269 M |
| 2030 | 993.153 M | 262.73 M | 15.1654 M | 264.109 M | 114.198 M |

### Figure S6: Component Costs by Scenario in Zimbabwe

Cost ($Millions)

| Year | ART | LEN | Oral PrEP | HIV related illness | HIV testing |
| --- | --- | --- | --- | --- | --- |
| 2026 | 31.8088 M | 0.449411 M | 3.82349 M | 4.24207 M | 6.70519 M |
| 2027 | 32.0856 M | 2.092 M | 3.81879 M | 4.07882 M | 6.84436 M |
| 2028 | 32.262 M | 3.82694 M | 3.83064 M | 3.94169 M | 6.99545 M |
| 2029 | 32.3891 M | 5.25565 M | 3.82648 M | 3.78174 M | 7.15022 M |
| 2030 | 32.4316 M | 5.53984 M | 3.8388 M | 3.59501 M | 7.31111 M |
| 2026 | 31.8145 M | 0.784594 M | 3.8159 M | 4.22801 M | 6.70962 M |
| 2027 | 32.1061 M | 3.65303 M | 3.8159 M | 4.08582 M | 6.83916 M |
| 2028 | 32.262 M | 6.72773 M | 3.80847 M | 3.91955 M | 6.98447 M |
| 2029 | 32.3413 M | 9.23922 M | 3.80282 M | 3.74473 M | 7.15369 M |
| 2030 | 32.3235 M | 9.764 M | 3.80813 M | 3.58453 M | 7.30697 M |
