## appendix 2 for "Health and budget impact, and price threshold for cost-effectiveness of lenacapavir for PrEP in Eastern and Southern Africa: a modeling analysis"

**Accompanying the manuscript:**

### Lenacapavir uptake estimates by subgroup

We conducted a scoping literature review of user preferences for oral vs. long-acting PrEP in Eastern and Southern Africa. We identified 34 publications published between 2014-2024 that assessed demand for long-acting PrEP products among the general population, adolescent girls and young women, and female sex workers. The preprint review is available here:

Brian Pfau, Arden Saravis, Sarah N. Cox, Linxuan Wu, Rachel Wittenauer, Emily Callen, Cory Arrouzet, Monisha Sharma. User Preferences on Long-Acting Pre-Exposure Prophylaxis for HIV Prevention in Sub-Saharan Africa: A Scoping Review medRxiv 2024.04.01.24305173; doi: <https://doi.org/10.1101/2024.04.01.24305173> (1)

Of 34 articles identified,15 evaluated long-acting injectable (LAI) PrEP and were considered relevant to the present analysis **(Table S1)**. For each of the 15 studies, we estimated the proportion of participants who reported an interest in using LAI PrEP if it were to become available, along with upper and lower ranges of uncertainty. We then summarized proportions across studies for each subgroup of interest. Across subgroups, long-acting injectable PrEP was more commonly preferred over daily oral PrEP, although was not universally preferred, with a minority of participants reporting a desire to use daily oral PrEP over LA PrEP.

We assumed healthcare accessibility among the population was not 100%, therefore a proportion of individuals with a desire to use LAI PrEP would be unable to access it. We conducted a literature review on healthcare accessibility among sugroups in ESA, including female sex workers (FSW), male clients of FSW, adolescent girls and young women (AGYW), and the general population. We found 7 studies assessing healthcare accessibility in adults and female sex workers seeking facility- or community-based healthcare for any or a specific conditions in ESA **(Table S2)**. We found an overall healthcare utilization of 20-96% among adults in the general population and 40-80% of FSW in SSA. Based on the data, we assumed that healthcare was accessible by 90% of women in the general population and 80% of FSWs, males and adolescent girls and young women.

We multiplied healthcare accessibility by LAI demand to estimate the Lenacapavir uptake by subgroup in ESA **(Table S3).**

Table 1: Summaries of studies evaluating healthcare accessibility in Eastern and Southern Africa

| Study | Population and settings | Condition/Service | Relevant Findings |
| --- | --- | --- | --- |
| **Adults** | | | |
| Bigogo 2010 (2) | Individuals with symptoms of illness in rural western Kenya | Fever, respiratory infections, and diarrhea | Among 208,007 illness episodes identified from 27,171 individuals, 57% sought care at a healthcare facility or community-based organization. |
| Abuya 2007 (3) | Adults with recent illnesses (i.e., fever) in three districts in Kenya with different levels of malaria endemicity | Fever | 57% of adults with fever purchased over-the-counter medicines |
| Panzner 2016 (4) | Adults with fever <3 days seeking public or private care in 9 sub-Saharan African countries | Fever | 20-88% of the population sought healthcare for fever at a healthcare facility across different sites |
| Abaerei 2017 (5) | Adult residents in Gauteng province, South Africa | Non-specific | 96% reported utilizing any healthcare services from public or private facilities |
| **Female Sex Workers (FSW)** | | | |
| Lafort 2017 (6) | FSW in SA, Mozambique, and Kenya | Facility based sexual reproductive health | 26-41% of the participants utilized facility-based SRH service across different study sites |
| Pande 2019 (7) | FSW in Uganda | HIV test in the past 12 months in facility, community, or outreach events | 86% of the participants reported having an HIV test in the last 12 months |
| Richter 2014 (8) | FSW in Johannesburg, Rustenburg and Cape Town, SA | Received facility or community-based health service in the last month | 60% of participants interacted with healthcare services in the last month |

Table S2: Characteristics of studies assessing PrEP preference in Sub-Saharan Africa

| **Author (year)** | **Study design** | **Study population** | **Method/question for preference assessment** | **Location** | **Findings** | **Methods for adapting study estimate for model parameters** | **% of participants who report preference for LAI PrEP** | | | |
| --- | --- | --- | --- | --- | --- | --- | --- | --- | --- | --- |
|  |  |  |  |  |  |  | **Adult Women** | **AGYW** | **Adult Men** | **FSW** |
| Ngure et al. (2021) (9) | Cross-sectional survey among a subset of women existing a PrEP adherence support study after 2 years' follow up | AGYW aged 18-24 at high HIV risk after finishing an oral PrEP RCT | If all these methods (vaginal ring, injection [such as contraception Depo provera], implants [such as Norplant], oral pills) were available, which one would you prefer? Rank in order of preference with 1 being the most preferred | Kenya | 59 (36.0%) of participants preferred injectables, 56 (34.2%) preferred oral pills, 36 (22.0%) preferred implants, and 24 (14.6%) preferred vaginal rings | We took the proportion of participants who preferred injectables (36%) as lower bound and proportion of participants who preferred any long-acting PrEP as the upper bound (36%+22%+14.6%=73%) |  | 36-73% |  |  |
| Wara et al. (2023) (10) | Cross-sectional survey of women participating in the PrEP-PP or PrIMA-X and attending follow up visits | Pregnant/postpartum women with oral prior PrEP experience median age 27-29 yo | Would you prefer to switch to injectable PrEP over remaining on oral PrEP if it were available? | South Africa, Kenya | 75% participants preferred long-acting injectable over oral PrEP |  | 75% |  |  |  |
| Minnis et al. (2020) (11) | Discrete choice experiment | Female and male aged 18-24 yo with no prior exposure to product | Assessed preference weights and trade-offs for long-acting PrEP: injectable and implant | South Africa | Youth indicated strong interest in using a long-acting HIV prevention product; for 96% of the choice tasks (7032/7263), respondents stated they would be willing to use their chosen product if available. |  |  | 96% |  |  |
| Montgomery et al. (2021) (12) | discrete choice experiment | MSM and MSW aged 18-24 years old; MSW product naive; MSM with PrEP experience | Assessed 5 attributes of LA PrEP (injectable vs. implant) including the delivery form, duration, insertion location, soreness and deliveyr facility | South Africa | Duration-dominant decision-makers” (46%) were largest class, defined by significant preference for a longer-duration product. “Comprehensive decision-makers” (36%) had preferences shaped equally by multiple attributes, and preferred implants. “Injection-dominant decision-makers” (18%) had strong preference for injections (vs. implant) and were significantly more likely to be MSM. When estimating shares for a 2-month injection in the buttocks with mild soreness (HPTN regimen) vs. a 6-month implant (to arm) with moderate soreness (current target), 95% of “injection-dominant” would choose injections, whereas 79% and 63% of “duration-dominant” and “comprehensive”, would choose implant. | We calculated the proportion of participants who preferred LAI by using the proportion of decision-maker multiplied by the corresponding proportion of who preferred LAI, 0.46*0.95+0.36*0.21+0.18*0.37=58% |  |  | 58% |  |
| Cheng et al. (2019) (13) | Cross sectional survey from a discrete choice experiment | Adult men, mostly MSW | Rank from most to least preferred: LAI, oral, and condoms | South Africa | 48% (n= 85) of participants preferred LAI PrEP, while 33% (n= 58) and 20% (n= 35) chose oral PrEP and condoms. |  |  |  | 48% |  |
| Tolley et al. (2019) (14) | Cross sectional survey among women participating in phase RCT of LA PrEP (HPTN 076) at baseline | Low HIV risk women median age 31 (24-37) years old | Quantitative question: initial preferences for future HIV prevention product use | South Africa, Zimbabwe, US | Among African women at baseline, 81% prefer bi month injection, 11% for daily oral pill, 2% for vaginal ring , 0% for vaginal gel , and 6% for other options (e.g.,implant, sterilization, IUD) | We took proportion of participants who preferred injectables (81%) as the lower bound and proportion of participants who preferred any long acting PrEP as the upper bound (81+2=83%) | 81-83% |  |  |  |
| van der Straten et al (2018) (15) | Preference of product after trying placebo product (injectable, ring, and oral) | Sexually active women aged 18-30 years | what were women's preferences for TRIO products compared to each other and to male condoms? | South Africa, Zimbabwe, Kenya | Overall, 85% preferred a TRIO product over condoms, and all three products were chosen by a significant number of women (injections 64%, tablets 21%, vaginal ring 15%) | We calculated the proportion of participants who preferred LAI by using the proportion of women who preferred a TRIO product multiplied by the corresponding proportion who preferred LAI as the lower bound and any long-acting product as the upper bound.  % prefer LAI: 85%*64% =54%,  % prefer any long-acting PrEP 85%*(64%+15%)=67% | 54-67% |  |  |  |
| Jansen van Vuuren et al. (2023) (16) | Cross-sectional survey among women enrolled in PrEP implementation project | Sexually active women aged 18-30 years | participants were asked to choose one method which would be most suitable to incorporate into their lifestyle | South Africa | The %(n) for each PrEP modality is: Three-monthly injection 31.3 (133), two-monthly injection 5.4 (23), daily oral PrEP 37.2 (158), pill before and after sex 8.0 (34), six-monthly implant 10.6 (45), monthly vaginal rings 5.4 (23), no method chosen 1.6(7), and missing 0.5 (2) | We took the proportion of participants who preferred injectables (31.3+5.4=37%) as the lower bound and proportion of participants who preferred any long acting PrEP as the upper bound (31.3+5.4+10.6+5.4=52.7). | 37-53% |  |  |  |
| Kidman et al. (2020) (17) | cross sectional survey among a subset of children enrolled in a cohort study | Girls age 10-16 years | If participants were willing to get an injection, and if they would prefer a daily pill to an injection | Malawi | Willing to have injection among sexually active girls is 91%; 53-65% of girl prefer injectable vs. oral daily | We took willingness to have injection as upper bound and preference for injectable over oral as the lower bound |  | 53-91% |  |  |
| Mayanja et al. (2022) (18) | Cross sectional:  Assessed preference during first two weeks of enrollment in PrEP cohort study among PrEP naive women | Sexually active women aged 14-24 (high HIV risk, 94% had paid sex) | Ranking 5 methods (oral PrEP, LAI, ring, implant, and vaccine) on a 1 to 5 scale (1=most preferred, 5=least preferred); considered preferred is ranked the top two choices | Uganda | Participants were offered oral PrEP during the study; 47.6% preferred oral PrEP | Assume LA as 1-47.6%=52% |  |  |  | 52% |
| Harling et al. (2019) (19) | Cross sectional survey among female bar workers (FBW) | FBW aged 23-29 yo with low exposure to PrEP before (5% heard of PrEP) | how interested in taking PrEP as daily pill, injection every 2 mo, vaginal gel, and monthly ring; rank the four modalities from most to least preferred. | Tanzania | 79% of FBW interested in LAI; 42% of FBW listed LAI as their first choice and 18% listed LAI as their second choice | We took those who were interested in LAI as the upper bound (i.e., 80%) and those listed LAI as their first and second choices as the lower bound(i.e., 60%) |  |  |  | 60-80% |
| Luecke et al. (2016) (20) | Cross sectional survey among women finishing the VOICE trial | Women aged 21–41 | select what type of product formulation, if any, they would be interested to take and explain their choice(s). Women could select as many products as they wanted | South Africa, Uganda, Zimbabwe | A majority of women (81%) expressed a preference for injectable, implant, or vaginal ring; 40% preferred to use LAI | We took 80% as the upper bound (80%) and 40% as the lower bound. | 40-80% |  |  |  |
| Beckham et al. (2022) (21) | Cross sectional survey among FSW | FSW with little exposure to PrEP | If you personally were going to take ART to prevent HIV infection, would you prefer to take it in the form of a daily pill or an injection once every 3 months? Injection/pill | Tanzania | Most (88%) preferred LA vs oral PrEP |  |  |  |  | 88% |
| Siedner et al. (2018) (22) | Cross sectional survey among clients from public sector clinic | Clients from public sector primary care with 86% being female and median age of 28 years (IQR 22-35); at least 22% had exposure to oral PrEP | If in the future an injectable PrEP would become available, would you have a preference for either oral or injectable PrEP? | Eswatini | Seventy-five percent of respondents (95%CI 65–83%) expressed a preference for LAI every two months over daily oral PrEP. This preference was reported in both sexes (P=0.16) |  | 65-83% |  | 65-83% |  |
| Were et al. (2023) (23) | Cross sectional survey from PrEP delivery site | Individuals eligible for PrEP from PrEP delivery clinics | Participants were presented with all PrEP options and asked to rank from most preferred to least preferred of injection, implant, monthly oral, and ED-PrEP (men only); Particiapnts also asked willingness to use each product | Kenya | 50% of AGYW, 54% of FSW, and 45% of male most prefer LAI. Slightly more than half(3,026;50.3%) of the participants indicated willingness to use two-month injectable PrEP (cabotegravir), with more females 2,052(67.8%) expressing preference. |  | 68% | 50% | 45-50% | 54% |
| Rousseau (2024) (24) | prospective cohort study | Young people | Let participants to choose from the following: oral PrEP, dapivirine vaginal ring (DVR), and CAB-LA | South Africa | As of July 20th, among 717 enrolled AGYW, 550 (77%) choose to use CAB-LA, 13 (2%) choose to use DVR, and 154 (21%) choose to use oral PrEP. | We took the proportion who used CAB-LA and DVR as the upper bound (i.e., 79%) and proportion who used CAB-LA as the lower bound(i.e., 77%) |  | 77-79% |  |  |

### Empiric Data for Model Calibration and Validation

#### HIV prevalence in counties by age and gender

##### Table S1a. HIV prevalence in counties of the former Nyanza province (Homa Bay, Kisii, Kisumu, Migori, Nyamira, Siaya), by age and gender in Kenya^¥^

| **Age group** | **2003** | | **2007** | | **2008** | | **2012** | | **2018** | |
| --- | --- | --- | --- | --- | --- | --- | --- | --- | --- | --- |
|  | **Men** | **Women** | **Men** | **Women** | **Men** | **Women** | **Men** | **Women** | **Men** | **Women** |
| **15 - 19** | 0.0015 | 0.0459 | 0.0121 | 0.0773 | 0.0184 | 0.1078 | 0.0151 | 0.0486 | 0.0025 | 0.0294 |
| **20 - 24** | 0.0562 | 0.2997 | 0.0257 | 0.2056 | 0.0578 | 0.1201 | 0.0289 | 0.1411 | 0.0216 | 0.0910 |
| **25 - 29** | 0.2429 | 0.2301 | 0.1956 | 0.2454 | 0.2450 | 0.2228 | 0.2107 | 0.2454 | 0.0705 | 0.2334 |
| **30 - 34** | 0.1840 | 0.1632 | 0.2578 | 0.2576 | 0.1530 | 0.2593 | 0.2392 | 0.2047 | 0.0911 | 0.2696 |
| **35- 39** | 0.2064 | 0.1838 | 0.2384 | 0.2227 | 0.2275 | 0.2259 | 0.1955 | 0.2811 | 0.1675 | 0.2705 |
| **40 - 44** | 0.2533 | 0.3500 | 0.2024 | 0.1799 | 0.2501 | 0.0927 | 0.3132 | 0.1694 | 0.2075 | 0.2765 |
| **45 - 49** | 0.1624 | 0.1651 | 0.2103 | 0.1291 | 0.1331 | 0.1716 | 0.1623 | 0.2287 | 0.2796 | 0.1991 |
| **15 - 49** | 0.1160 | 0.1830 | 0.1140 | 0.1760 | 0.1140 | 0.1600 | 0.1340 | 0.1760 | 0.0826 | 0.1667 |

**^¥^Sources:** Kenya Demographic and Health Surveys, 2003 & 2008; Kenya AIDS Indicator Surveys, 2007 & 2012; Kenya Population-Based HIV Impact Assessment 2018.

##### Table S1b. HIV prevalence data by age and sex from population-based surveys for model calibration in South Africa^¥^

| **Sex** | **Age group** | **Year** | | | | |
| --- | --- | --- | --- | --- | --- | --- |
|  |  | **2002** | **2005** | **2008** | **2012** | **2017** |
| **Men** | 15-19 | 0.040 | 0.032 | 0.025 | 0.007 | 0.047 |
|  | 20-24 | 0.080 | 0.060 | 0.051 | 0.051 | 0.048 |
|  | 25-29 | 0.220 | 0.121 | 0.157 | 0.173 | 0.124 |
|  | 30-34 | 0.240 | 0.233 | 0.258 | 0.256 | 0.184 |
|  | 35-39 | 0.180 | 0.233 | 0.185 | 0.288 | 0.238 |
|  | 40-44 | 0.120 | 0.175 | 0.192 | 0.158 | 0.224 |
|  | 45-49 | 0.120 | 0.103 | 0.084 | 0.134 | 0.248 |
|  | 50-54 | 0.050 | 0.142 | 0.104 | 0.155 | 0.202 |
|  | 55-59 | 0.070 | 0.064 | 0.062 | 0.055 | 0.148 |
|  | 15-49 | 0.128 | 0.117 | 0.116 | 0.145 | 0.148 |
| **Women** | 15-19 | 0.070 | 0.094 | 0.067 | 0.056 | 0.058 |
|  | 20-24 | 0.170 | 0.239 | 0.211 | 0.174 | 0.156 |
|  | 25-29 | 0.320 | 0.333 | 0.327 | 0.284 | 0.275 |
|  | 30-34 | 0.240 | 0.260 | 0.291 | 0.360 | 0.347 |
|  | 35-39 | 0.140 | 0.193 | 0.248 | 0.316 | 0.394 |
|  | 40-44 | 0.190 | 0.124 | 0.163 | 0.280 | 0.359 |
|  | 45-49 | 0.110 | 0.087 | 0.141 | 0.197 | 0.303 |
|  | 50-54 | 0.080 | 0.075 | 0.102 | 0.148 | 0.222 |
|  | 55-59 | 0.070 | 0.030 | 0.077 | 0.097 | 0.176 |
|  | 15-49 | 0.177 | 0.202 | 0.213 | 0.232 | 0.263 |

^¥^Sources: South African National HIV Prevalence, Incidence and Behaviour Surveys (2002, 2005, 2008, 2012 and 2017) from the Human Sciences Research Council (HSRC)

##### Table S1c. HIV prevalence data by age and sex from population-based surveys for model calibration in Zimbabwe^¥^

| **Age group** | **2006** | | **2011** | | **2015** | | **2016** | |
| --- | --- | --- | --- | --- | --- | --- | --- | --- |
|  | **Men** | **Women** | **Men** | **Women** | **Men** | **Women** | **Men** | **Women** |
| **15 - 19** | 0.031 | 0.062 | 0.034 | 0.042 | 0.025 | 0.040 | 0.032 | 0.039 |
| **20 - 24** | 0.058 | 0.163 | 0.038 | 0.106 | 0.037 | 0.103 | 0.027 | 0.081 |
| **25 - 29** | 0.130 | 0.288 | 0.103 | 0.201 | 0.075 | 0.155 | 0.066 | 0.143 |
| **30 - 34** | 0.295 | 0.355 | 0.174 | 0.290 | 0.131 | 0.219 | 0.122 | 0.219 |
| **35- 39** | 0.321 | 0.345 | 0.251 | 0.291 | 0.180 | 0.280 | 0.194 | 0.266 |
| **40 - 44** | 0.329 | 0.257 | 0.262 | 0.257 | 0.270 | 0.313 | 0.254 | 0.296 |
| **45 - 49** | 0.260 | 0.180 | 0.299 | 0.225 | 0.232 | 0.243 | 0.281 | 0.289 |
| **15 - 49** | 0.145 | 0.211 | 0.123 | 0.177 | 0.105 | 0.167 | - | - |

Zimbabwe Demographic and Health Surveys 2005-06, 2010-22, 2015. Zimbabwe Population-Based HIV Impact Assessment 2016.

##### Table S2. HIV prevalence among men and women ages 15-49, by county and gender in Kenya^¥^

| **County** | **2003** | | **2007** | | **2008** | | **2012** | | **2018** | |
| --- | --- | --- | --- | --- | --- | --- | --- | --- | --- | --- |
|  | **Men** | **Women** | **Men** | **Women** | **Men** | **Women** | **Men** | **Women** | **Men** | **Women** |
| **Homa Bay** | 0.1097 | 0.2458 | 0.2514 | 0.3259 | 0.1737 | 0.2524 | 0.2217 | 0.2787 | 0.1279 | 0.2532 |
| **Kisii** | 0.0114 | 0.0853 | 0.0445 | 0.0693 | 0.0330 | 0.0573 | 0.0346 | 0.0368 | 0.0458 | 0.0684 |
| **Kisumu** | 0.1663 | 0.1914 | 0.1139 | 0.1847 | 0.1109 | 0.1810 | 0.1940 | 0.2022 | 0.0960 | 0.2096 |
| **Migori** | 0.1804 | 0.1860 | 0.1685 | 0.2181 | 0.1923 | 0.2228 | 0.1435 | 0.1925 | 0.0706 | 0.1758 |
| **Nyamira** | 0.0029 | 0.0742 | - | - | 0.0234 | 0.0544 | 0.0419 | 0.1045 | 0.0247 | 0.0432 |
| **Siaya** | 0.1824 | 0.2424 | 0.1445 | 0.2130 | 0.1526 | 0.1921 | 0.2596 | 0.2990 | 0.0961 | 0.1905 |

**^¥^Sources:** Kenya Demographic and Health Surveys, 2003 & 2008; Kenya AIDS Indicator Surveys, 2007 & 2012; Kenya Population-Based HIV Impact Assessment 2018.

#### Number of people on ART

##### Table S3a. Number of people on ART by county, gender, and age group in Kenya^¥^

| **Gender** | **County** | **Age group** | | **Year** | | | | | | | | | | | | | |
| --- | --- | --- | --- | --- | --- | --- | --- | --- | --- | --- | --- | --- | --- | --- | --- | --- | --- |
|  |  |  |  | **2004** | **2005** | **2006** | **2007** | **2008** | **2009** | **2010** | **2011** | **2012** | **2013** | **2014** | **2015** | **2016** | **2017** |
| **Men** | **Homa Bay** | 0 - 14 | | - | - | - | - | - | - | - | - | - | - | 2,945 | 3,583 | 4,109 | 4,192 |
|  |  | 15 - 99 | | 1,067 | 2,313 | 5,148 | 7,194 | 10,002 | 14,436 | 17,178 | 15,954 | 17,522 | 18,279 | 19,157 | 22,834 | 26,441 | 29,220 |
|  | **Kisii** | 0 - 14 | | - | - | - | - | - | - | - | - | - | - | 828 | 993 | 1,109 | 1,083 |
|  |  | 15 - 99 | | - | - | - | - | - | - | - | 2,972 | - | - | 4,614 | 5,451 | 6,604 | 7,169 |
|  | **Kisumu** | 0 - 14 | | - | - | - | - | - | - | - | - | - | - | 3,101 | 3,245 | 3,525 | 3,607 |
|  |  | 15 - 99 | | 945 | 2,047 | 4,557 | 6,368 | 8,853 | 12,779 | 15,206 | 14,122 | 15,511 | 16,180 | 21,216 | 24,550 | 28,082 | 31,021 |
|  | **Migori** | 0 - 14 | |  |  |  |  |  |  |  |  |  |  | 2,309 | 2,295 | 2,678 | 2,673 |
|  |  | 15 - 99 | | 711 | 1,541 | 3,430 | 4,793 | 6,664 | 9,619 | 11,446 | 10,630 | 11,675 | 12,179 | 13,929 | 15,165 | 17,438 | 18,455 |
|  | **Nyamira** | 0 - 14 | | - | - | - | - | - | - | - | - | - | - | 484 | 552 | 578 | 611 |
|  |  | 15 - 99 | | - | - | - | - | - | - | - | 1,362 | - | - | 2,120 | 2,585 | 3,142 | 3,474 |
|  | **Siaya** | 0 - 14 | | - | - | - | - | - | - | - | - | - | - | 2,645 | 2,950 | 3,017 | 3,197 |
|  |  | 15 - 99 | | 860 | 1,864 | 4,148 | 5,797 | 8,060 | 11,633 | 13,843 | 12,856 | 14,120 | 14,730 | 16,163 | 18,611 | 21,477 | 23,762 |
| **Women** | **Homa Bay** | 0 - 14 | | - | - | - | - | - | - | - | - | - | - | 3,431 | 3,835 | 4,426 | 4,535 |
|  |  | 15 - 99 | | 1,359 | 2,944 | 6,551 | 9,155 | 12,522 | 17,202 | 21,798 | 31,454 | 35,182 | 38,819 | 40,118 | 49,956 | 57,286 | 61,811 |
|  | **Kisii** | 0 - 14 | | - | - | - | - | - | - | - | - | - | - | 906 | 1,079 | 1,200 | 1,146 |
|  |  | 15 - 99 | | - | - | - | - | - | - | - | 7,902 | - | - | 11,691 | 14,350 | 17,274 | 19,044 |
|  | **Kisumu** | 0 - 14 | | - | - | - | - | - | - | - | - | - | - | 3,241 | 3,393 | 3,810 | 3,831 |
|  |  | 15 - 99 | | 1,203 | 2,606 | 5,799 | 8,104 | 11,084 | 15,227 | 19,296 | 27,843 | 31,143 | 34,362 | 41,230 | 48,424 | 56,384 | 60,789 |
|  | **Migori** | 0 - 14 | | - | - | - | - | - | - | - | - | - | - | 2,526 | 2,448 | 2,868 | 2,884 |
|  |  | 15 - 99 | | 905 | 1,961 | 4,365 | 6,100 | 8,343 | 11,461 | 14,524 | 20,958 | 23,442 | 25,865 | 27,896 | 31,964 | 38,637 | 40,891 |
|  | **Nyamira** | 0 - 14 | | - | - | - | - | - | - | - | - | - | - | 506 | 567 | 601 | 622 |
|  |  | 15 - 99 | | - | - | - | - | - | - | - | 3,766 | - | - | 5,964 | 7,210 | 8,258 | 8,654 |
|  | **Siaya** | 0 - 14 | | - | - | - | - | - | - | - | - | - | - | 2,778 | 3,136 | 3,299 | 3,569 |
|  |  | 15 - 99 | | 1,095 | 2,372 | 5,279 | 7,377 | 10,090 | 13,862 | 17,566 | 25,347 | 28,351 | 31,281 | 33,911 | 39,853 | 44,892 | 48,808 |
| **Both** | **All** | | 15-99 | 8,954 | 19,404 | 43,185 | 60,350 | 83,142 | 116,787 | 143,877 | 175,003 | 194,552 | 210,769 | 246,293 | 301,029 | 333,333 | 389,159 |

**^¥^**Source: Kenya Ministry of Health

##### Table S3b: Number of people on ART by sex (ages 15-49 years) in South Africa ^¥^

| **Year** | **Male** | **Female** |
| --- | --- | --- |
| 2001 | 2713 | 3543 |
| 2002 | 5768 | 7586 |
| 2003 | 9321 | 12313 |
| 2004 | 17717 | 24423 |
| 2005 | 34874 | 59240 |
| 2006 | 66629 | 123308 |
| 2007 | 118977 | 228753 |
| 2008 | 186564 | 367389 |
| 2009 | 277931 | 548206 |
| 2010 | 402000 | 781477 |
| 2011 | 558131 | 1095411 |
| 2012 | 713294 | 1415016 |
| 2013 | 876749 | 1732515 |
| 2014 | 1013499 | 2003014 |
| 2015 | 1130063 | 2239669 |
| 2016 | 1238815 | 2518238 |
| 2017 | 1403702 | 2998170 |

^¥^Source: South Africa Department of Health Surveys

##### Table S3c: Number of people on ART by year and sex (ages 15-49 years) in Zimbabwe ^¥^

| Year | Male | Female |
| --- | --- | --- |
| 2004 | 4854 | 6146 |
| 2005 | 10049 | 15818 |
| 2006 | 22323 | 31309 |
| 2007 | 37527 | 52562 |
| 2008 | 57603 | 77120 |
| 2009 | 82266 | 117364 |
| 2010 | 119235 | 211144 |
| 2011 | 148551 | 296112 |
| 2012 | 178027 | 340774 |
| 2013 | 245690 | 373190 |
| 2014 | 264614 | 468305 |
| 2016 | 361399 | 579378 |
| 2017 | 388658 | 663132 |

^¥^Zimbabwe Ministry of Health

#### Population size by age and gender

##### Table S4a. Population size by gender, county, and age group in 2009 in western Kenya^¥^

| **Age Group** | **Men** | | | | | | **Women** | | | | | |
| --- | --- | --- | --- | --- | --- | --- | --- | --- | --- | --- | --- | --- |
|  | **Homa Bay** | **Kisii** | **Kisumu** | **Migori** | **Nyamira** | **Siaya** | **Homa Bay** | **Kisii** | **Kisumu** | **Migori** | **Nyamira** | **Siaya** |
| **0 - < 1** | 18,335 | 18,236 | 17,457 | 19,265 | 10,313 | 15,093 | 18,354 | 17,993 | 16,926 | 19,309 | 10,263 | 14,860 |
| **1 - 4** | 69,799 | 69,529 | 63,054 | 69,921 | 41,165 | 56,269 | 69,250 | 69,023 | 63,172 | 69,519 | 40,396 | 55,901 |
| **5 - 9** | 75,926 | 76,757 | 67,083 | 73,872 | 46,450 | 60,966 | 75,973 | 75,778 | 67,779 | 74,333 | 46,867 | 60,710 |
| **10 - 14** | 68,689 | 68,473 | 62,706 | 64,300 | 42,590 | 58,296 | 67,159 | 68,072 | 63,359 | 63,249 | 42,198 | 56,248 |
| **15 - 19** | 57,430 | 59,228 | 55,597 | 53,075 | 36,604 | 49,220 | 54,119 | 60,776 | 56,741 | 52,238 | 36,786 | 47,825 |
| **20 - 24** | 39,573 | 41,898 | 47,281 | 38,690 | 24,409 | 32,725 | 50,309 | 58,225 | 57,649 | 48,004 | 34,184 | 41,443 |
| **25 - 29** | 30,437 | 32,792 | 40,964 | 30,727 | 19,515 | 25,961 | 36,016 | 42,878 | 40,614 | 34,670 | 27,273 | 30,135 |
| **30 - 34** | 23,259 | 26,678 | 30,412 | 23,344 | 16,605 | 20,359 | 26,342 | 30,031 | 27,515 | 25,630 | 19,487 | 22,328 |
| **34 - 39** | 16,013 | 21,766 | 21,251 | 17,024 | 14,039 | 14,793 | 20,010 | 26,051 | 20,611 | 19,313 | 17,106 | 17,932 |
| **40 - 44** | 11,914 | 15,718 | 15,145 | 12,170 | 10,470 | 11,118 | 16,513 | 18,360 | 16,894 | 14,773 | 11,377 | 16,082 |
| **45 - 49** | 11,124 | 16,797 | 13,361 | 10,549 | 11,318 | 10,390 | 15,248 | 19,181 | 15,298 | 12,888 | 11,886 | 15,486 |
| **50 - 54** | 9,705 | 12,789 | 11,251 | 8,565 | 8,379 | 9,079 | 12,942 | 14,136 | 12,504 | 10,314 | 8,703 | 14,541 |
| **55 - 59** | 8,159 | 9,527 | 8,718 | 6,399 | 5,999 | 8,414 | 9,833 | 9,528 | 9,175 | 7,692 | 5,819 | 12,265 |
| **60 - 64** | 6,989 | 7,395 | 7,054 | 5,250 | 5,026 | 7,712 | 8,587 | 7,654 | 7,597 | 6,000 | 5,107 | 11,081 |
| **65 - 69** | 4,325 | 4,637 | 4,163 | 3,382 | 3,094 | 5,107 | 5,957 | 5,320 | 5,402 | 4,508 | 3,322 | 7,732 |
| **70 - 74** | 4,029 | 3,945 | 3,777 | 2,907 | 2,753 | 5,175 | 5,355 | 5,017 | 4,757 | 3,524 | 3,153 | 7,173 |
| **75 - 79** | 2,835 | 2,743 | 2,392 | 2,033 | 1,778 | 3,549 | 3,891 | 3,338 | 3,356 | 2,968 | 1,919 | 5,464 |
| **80 - 99** | 3,726 | 3,701 | 2,821 | 2,624 | 2,393 | 4,159 | 5,316 | 5,891 | 4,615 | 3,636 | 3,475 | 6,155 |

**^¥^**Source: Kenya National Bureau of Statistics, 2009 Census

##### Table S4b. Population of South Africa by age and sex ^¥^

| **Sex** | **Age group** | **Year** | | | | |
| --- | --- | --- | --- | --- | --- | --- |
|  |  | **2002** | **2005** | **2008** | **2012** | **2017** |
| **Men** | 0 - 4 | 2634839 | 2651819 | 2692300 | 2784372 | 2886299 |
|  | 5 - 9 | 2559246 | 2570683 | 2592577 | 2644720 | 2767111 |
|  | 10 - 14 | 2582620 | 2565499 | 2557928 | 2578751 | 2636981 |
|  | 15 - 19 | 2546968 | 2596517 | 2584945 | 2572032 | 2587023 |
|  | 20 - 24 | 2348165 | 2488613 | 2572307 | 2606447 | 2594656 |
|  | 25 - 29 | 2069939 | 2215800 | 2362377 | 2530034 | 2612453 |
|  | 30 - 34 | 1759391 | 1899279 | 2018461 | 2227862 | 2488823 |
|  | 35 - 39 | 1488894 | 1568198 | 1664587 | 1845019 | 2131687 |
|  | 40 - 44 | 1270053 | 1324166 | 1367150 | 1493652 | 1731088 |
|  | 45 - 49 | 1077752 | 1122009 | 1159644 | 1234887 | 1398602 |
|  | 50 - 54 | 831061 | 925655 | 973184 | 1038560 | 1147992 |
|  | 55 - 59 | 636945 | 677128 | 760379 | 856728 | 947123 |
| **Women** | 0 - 4 | 2587465 | 2602664 | 2640997 | 2729586 | 2825703 |
|  | 5 - 9 | 2523770 | 2535299 | 2555462 | 2604517 | 2719700 |
|  | 10 - 14 | 2557891 | 2533039 | 2526253 | 2545219 | 2603685 |
|  | 15 - 19 | 2527828 | 2568409 | 2552976 | 2535920 | 2555420 |
|  | 20 - 24 | 2323512 | 2451570 | 2523985 | 2560705 | 2550570 |
|  | 25 - 29 | 2064262 | 2167428 | 2282514 | 2447992 | 2550351 |
|  | 30 - 34 | 1807453 | 1906808 | 1967958 | 2123170 | 2394266 |
|  | 35 - 39 | 1577255 | 1648612 | 1717873 | 1821711 | 2039776 |
|  | 40 - 44 | 1370835 | 1434647 | 1491937 | 1594914 | 1745050 |
|  | 45 - 49 | 1184477 | 1243314 | 1302936 | 1396666 | 1541175 |
|  | 50 - 54 | 924873 | 1054059 | 1124243 | 1214556 | 1348848 |
|  | 55 - 59 | 727478 | 783876 | 898804 | 1035986 | 1160340 |

^¥^Source: United Nations Population Database

##### Table S4c. Population of Zimbabwe by age and sex in 2010^¥^

| Age Group (years) | Male | Female |
| --- | --- | --- |
| 0-4 | 1003778 | 993503 |
| 5-9 | 849773 | 849884 |
| 10-14 | 791114 | 795446 |
| 15-19 | 729897 | 766954 |
| 20-24 | 646316 | 730145 |
| 25-29 | 511221 | 596614 |
| 30-34 | 407730 | 471291 |
| 35-39 | 296577 | 333524 |
| 40-44 | 220200 | 245985 |
| 45-49 | 163800 | 200252 |
| 50-54 | 125947 | 167229 |
| 55-59 | 99658 | 142147 |
| 60-64 | 68299 | 100376 |
| 65-69 | 60276 | 94053 |
| 70-74 | 39694 | 70357 |
| 75-79 | 21954 | 48407 |
| 80-84 | 9792 | 28377 |
| 85-89 | 2574 | 10891 |
| 90-94 | -- | 2858 |

Source: Zimbabwe National Statistics Agency.

#### Voluntary medical male circumcision

##### Table S5a. Circumcision status quo by county, age group, and year in Kenya ^¥^

|  | **Homa Bay** | | | **Kisii** | | | **Kisumu** | | | | **Migori** | | | | **Nyamira** | | | | **Siaya** | | |
| --- | --- | --- | --- | --- | --- | --- | --- | --- | --- | --- | --- | --- | --- | --- | --- | --- | --- | --- | --- | --- | --- |
| **Year** | **10-14** | **15-24** | **25-49** | **10-14** | **15-24** | **25-49** | **10-14** | **15-24** | **25-49** | **10-14** | | **15-24** | **25-49** | **10-14** | | **15-24** | **25-49** | **10-14** | | **15-24** | **25-49** |
| **Pre-2008** | 0.249 | 0.249 | 0.249 | 0.948 | 0.948 | 0.948 | 0.322 | 0.322 | 0.322 | 0.410 | | 0.410 | 0.410 | 0.965 | | 0.965 | 0.965 | 0.252 | | 0.252 | 0.252 |
| **2008** | 0.118 | 0.235 | 0.275 | 0.948 | 0.948 | 0.948 | 0.152 | 0.303 | 0.333 | 0.194 | | 0.385 | 0.451 | 0.965 | | 0.965 | 0.965 | 0.119 | | 0.243 | 0.270 |
| **2009** | 0.122 | 0.240 | 0.283 | 0.948 | 0.948 | 0.948 | 0.178 | 0.315 | 0.339 | 0.199 | | 0.388 | 0.463 | 0.965 | | 0.965 | 0.965 | 0.127 | | 0.250 | 0.277 |
| **2010** | 0.166 | 0.285 | 0.299 | 0.948 | 0.948 | 0.948 | 0.324 | 0.389 | 0.362 | 0.214 | | 0.395 | 0.478 | 0.965 | | 0.965 | 0.965 | 0.222 | | 0.310 | 0.293 |
| **2011** | 0.226 | 0.355 | 0.313 | 0.948 | 0.948 | 0.948 | 0.450 | 0.479 | 0.385 | 0.280 | | 0.423 | 0.486 | 0.965 | | 0.965 | 0.965 | 0.293 | | 0.375 | 0.304 |
| **2012** | 0.337 | 0.486 | 0.339 | 0.948 | 0.948 | 0.948 | 0.516 | 0.559 | 0.409 | 0.516 | | 0.530 | 0.509 | 0.965 | | 0.965 | 0.965 | 0.417 | | 0.483 | 0.324 |
| **2013** | 0.368 | 0.565 | 0.361 | 0.948 | 0.948 | 0.948 | 0.564 | 0.634 | 0.436 | 0.587 | | 0.614 | 0.528 | 0.965 | | 0.965 | 0.965 | 0.435 | | 0.549 | 0.340 |
| **2014** | 0.471 | 0.710 | 0.399 | 0.948 | 0.948 | 0.948 | 0.620 | 0.712 | 0.467 | 0.717 | | 0.726 | 0.554 | 0.965 | | 0.965 | 0.965 | 0.537 | | 0.659 | 0.368 |
| **2015** | 0.506 | 0.811 | 0.438 | 0.948 | 0.948 | 0.948 | 0.672 | 0.790 | 0.502 | 0.742 | | 0.813 | 0.580 | 0.965 | | 0.965 | 0.965 | 0.562 | | 0.739 | 0.395 |
| **2016** | 0.476 | 0.894 | 0.488 | 0.948 | 0.948 | 0.948 | 0.756 | 0.844 | 0.538 | 0.697 | | 0.891 | 0.613 | 0.965 | | 0.965 | 0.965 | 0.628 | | 0.841 | 0.425 |
| **2017** | 0.489 | 0.937 | 0.537 | 0.948 | 0.948 | 0.948 | 0.863 | 0.889 | 0.572 | 0.680 | | 0.949 | 0.648 | 0.965 | | 0.965 | 0.965 | 0.759 | | 0.917 | 0.457 |
| **2018** | 0.589 | 0.925 | 0.637 | 0.948 | 0.948 | 0.948 | 0.944 | 0.925 | 0.651 | 0.775 | | 0.976 | 0.744 | 0.965 | | 0.965 | 0.965 | 0.844 | | 0.959 | 0.562 |
| **2019** | 0.741 | 0.904 | 0.678 |  |  |  | 0.784 | 0.943 | 0.677 | 0.785 | |  | 0.775 |  | |  |  | 0.790 | | 0.976 | 0.596 |
| **2020** | 0.802 | 0.901 | 0.717 |  |  |  | 0.787 | 0.925 | 0.703 | 0.800 | |  | 0.808 |  | |  |  | 0.800 | |  | 0.633 |
| **2021** | 0.802 | 0.909 | 0.749 |  |  |  | 0.836 | 0.913 | 0.727 |  | |  | 0.836 |  | |  |  |  | |  | 0.668 |
| **2022** |  | 0.917 | 0.779 |  |  |  | 0.804 | 0.947 | 0.750 |  | |  | 0.865 |  | |  |  |  | |  | 0.704 |
| **2023** |  | 0.926 | 0.807 |  |  |  | 0.804 |  | 0.773 |  | |  | 0.892 |  | |  |  |  | |  | 0.738 |
| **2024** |  | 0.935 | 0.833 |  |  |  | 0.804 |  | 0.795 |  | |  | 0.918 |  | |  |  |  | |  | 0.772 |
| **2025** |  | 0.944 | 0.858 |  |  |  | 0.804 |  | 0.815 |  | |  | 0.942 |  | |  |  |  | |  | 0.803 |
| **2026** |  | 0.949 | 0.879 |  |  |  | 0.801 |  | 0.832 |  | |  | 0.961 |  | |  |  |  | |  | 0.832 |
| **2027** |  | 0.953 | 0.898 |  |  |  | 0.801 |  | 0.848 |  | |  | 0.976 |  | |  |  |  | |  | 0.976 |
| **2028** |  | 0.955 | 0.915 |  |  |  | 0.801 |  | 0.862 |  | |  |  |  | |  |  |  | |  |  |
| **2029** |  |  | 0.931 |  |  |  | 0.801 |  | 0.874 |  | |  |  |  | |  |  |  | |  |  |
| **2030** |  |  | 0.945 |  |  |  | 0.801 |  | 0.885 |  | |  |  |  | |  |  |  | |  |  |
| **2031** |  |  | 0.955 |  |  |  | 0.802 |  | 0.897 |  | |  |  |  | |  |  |  | |  |  |
| **2032** |  |  |  |  |  |  |  |  | 0.907 |  | |  |  |  | |  |  |  | |  |  |
| **2033** |  |  |  |  |  |  |  |  | 0.917 |  | |  |  |  | |  |  |  | |  |  |
| **2034** |  |  |  |  |  |  |  |  | 0.924 |  | |  |  |  | |  |  |  | |  |  |
| **2035** |  |  |  |  |  |  |  |  | 0.931 |  | |  |  |  | |  |  |  | |  |  |

**^¥^Source:** Circumcision prevalence prior to 2008 is obtained from the Kenya Demographic and Health Survey, 2003. Prevalence of circumcision from 2008 onward combines prevalence of traditional male circumcision and voluntary medical male circumcision estimates obtained from the Decision-Makers' Program Planning Toolkit 2.

##### Table S5b. Number of voluntary medical male circumcisions conducted in South Africa by age group^*^

| **Year** | **Age Group** | | | | | |
| --- | --- | --- | --- | --- | --- | --- |
|  | **10 - 14** | **15 - 19** | **20 - 24** | **25 - 34** | **35 - 49** | **≥50** |
| **2010** | 55,431 | 30,552 | 15,856 | 18,047 | 7,864 | 1,160 |
| **2011** | 137,648 | 75,866 | 39,374 | 44,816 | 19,527 | 2,881 |
| **2012** | 175,060 | 96,487 | 50,075 | 56,996 | 24,834 | 3,664 |
| **2013** | 156,496 | 86,255 | 44,765 | 50,952 | 22,201 | 3,276 |
| **2014** | 199,750 | 110,095 | 57,138 | 65,035 | 28,337 | 4,181 |
| **2015** | 199,535 | 109,976 | 57,076 | 64,965 | 28,306 | 4,176 |
| **2016** | 165,672 | 91,312 | 47,390 | 53,940 | 23,502 | 3,468 |
| **2017** | 203,960 | 112,415 | 58,342 | 66,406 | 28,934 | 4,269 |
| **2018** | 279,500 | 154,050 | 79,950 | 91,000 | 39,650 | 5,850 |
| **2019** | 258,000 | 142,200 | 73,800 | 84,000 | 36,600 | 5,400 |
| **2020** | 236,500 | 130,350 | 67,650 | 77,000 | 33,550 | 4,950 |
| **2021** | 107,500 | 59,250 | 30,750 | 35,000 | 15,250 | 2,250 |
| **2022 onwards** | 43,000 | 23,700 | 12,300 | 14,000 | 6,100 | 900 |

^*^Source: South Africa Department of Health (unpublished data) and South Africa National Strategic Plan for HIV, TB, and STIs 2017-2022(25).

##### Table S5c. Number of voluntary medical male circumcisions conducted in Zimbabwe among male aged 15-25 years old^*^

| **Year** | **Target # of  males**  **(15-25 years old)** |
| --- | --- |
| **2008.5** | 2784 |
| **2009.5** | 9381 |
| **2010.5** | 27973 |
| **2011.5** | 29321 |
| **2012.5** | 65679 |
| **2013.5** | 110163 |
| **2014.5** | 133012 |
| **2015.5** | 136986 |
| **2016.5** | 159476 |
| **2017.5** | 197705 |
| **2018.5** | 207375 |
| **2019.5** | 53743 |
| **2020.5** | 53743 |
| **2021.5-2040.5** | 4288000 |

Source: McGillen JB, Stover J, Klein DJ, Xaba Sinokuthemba, et al. The emerging health impact of voluntary medical male circumcision in Zimbabwe: An evaluation using three epidemiological models. Jul 2018 PLOS One. https://doi.org/10.1371/journal.pone.0199453

#### Age-specific population fertility rates

##### Table S5a. Age-specific population fertility rates in Kenya 1950-2044^¥^

|  | **Age-specific fertility rates (births per 1,000 women)** | | | | | | |
| --- | --- | --- | --- | --- | --- | --- | --- |
| **Year** | **15-19** | **20-24** | **25-29** | **30-34** | **35-39** | **40-44** | **45-49** |
| **1950-1955** | 169.1 | 351.6 | 338.1 | 284.3 | 203.5 | 110.7 | 38.9 |
| **1955-1960** | 175.9 | 365.9 | 351.9 | 295.8 | 211.8 | 115.2 | 40.5 |
| **1960-1965** | 182.3 | 379.1 | 364.5 | 306.5 | 219.4 | 119.4 | 41.9 |
| **1965-1970** | 183.3 | 381.2 | 366.6 | 308.2 | 220.6 | 120 | 42.2 |
| **1970-1975** | 180.6 | 375.5 | 361.1 | 303.6 | 217.3 | 118.3 | 41.5 |
| **1975-1980** | 172.7 | 359.1 | 345.3 | 290.3 | 207.8 | 113.1 | 39.7 |
| **1980-1985** | 163.1 | 339.2 | 326.2 | 274.2 | 196.3 | 106.8 | 37.5 |
| **1985-1990** | 147.8 | 307.3 | 295.5 | 248.4 | 177.8 | 96.8 | 34.0 |
| **1990-1995** | 115.3 | 268.9 | 252.0 | 206.8 | 161.6 | 73.4 | 52.0 |
| **1995-2000** | 111.5 | 260.7 | 253.3 | 196.2 | 143.3 | 62.4 | 42.7 |
| **2000-2005** | 104.2 | 243.6 | 236.7 | 183.4 | 133.9 | 58.3 | 39.9 |
| **2005-2010** | 97.1 | 227.1 | 221.4 | 170.6 | 123.7 | 53.6 | 36.5 |
| **2010-2015** | 86.2 | 201.9 | 202.3 | 149.2 | 102.1 | 42.4 | 27.9 |
| **2015-2020** | 75.1 | 176.5 | 179.8 | 129.6 | 85.8 | 34.8 | 22.4 |
| **2020-2024** | 69.9 | 165.1 | 171.8 | 120.2 | 76.2 | 29.9 | 18.6 |
| **2025-2029** | 65.0 | 154.7 | 164.8 | 112.6 | 68.5 | 26.0 | 15.5 |
| **2030-2034** | 60.6 | 145.7 | 159.1 | 106.7 | 62.5 | 22.9 | 13.1 |
| **2035-2039** | 56.2 | 137.2 | 153.6 | 101.8 | 57.6 | 20.4 | 11.0 |
| **2040-2044** | 52.2 | 129.7 | 149.3 | 98.2 | 53.8 | 18.5 | 9.4 |

**^¥^Source**: 2019 World Population Prospects

##### Table S5b. Age-specific population fertility rates in South Africa 1950-2044^¥^

|  | **Age-specific fertility rates (births per 1,000 women)** | | | | | | |
| --- | --- | --- | --- | --- | --- | --- | --- |
| **Year** | **15-19** | **20-24** | **25-29** | **30-34** | **35-39** | **40-44** | **45-49** |
| **1950-1955** | 66.8 | 265 | 291.9 | 242.2 | 189.8 | 132 | 72.3 |
| **1955-1960** | 65.7 | 260.8 | 287.3 | 238.3 | 186.7 | 130 | 71.2 |
| **1960-1965** | 64.7 | 256.6 | 282.7 | 234.5 | 183.7 | 127.9 | 70 |
| **1965-1970** | 60.4 | 239.7 | 264.1 | 219.1 | 171.7 | 119.5 | 65.4 |
| **1970-1975** | 76.1 | 233.9 | 253.6 | 211 | 160.2 | 105.1 | 54 |
| **1975-1980** | 86.1 | 217.3 | 231.8 | 193.6 | 142.3 | 87.4 | 41.5 |
| **1980-1985** | 93.6 | 201.1 | 211.3 | 177 | 125.9 | 71.7 | 30.5 |
| **1985-1990** | 95.4 | 179.4 | 185.7 | 155.9 | 107.3 | 56 | 20.4 |
| **1990-1995** | 90.8 | 152.2 | 155.2 | 130.8 | 86.9 | 41 | 11.7 |
| **1995-2000** | 80.6 | 140.5 | 142.5 | 111.5 | 74.4 | 31 | 10.3 |
| **2000-2005** | 70.7 | 139 | 141.8 | 105.6 | 67.4 | 27.1 | 8.8 |
| **2005-2010** | 59.2 | 131.7 | 135.1 | 95.9 | 58.4 | 22.6 | 7.1 |
| **2010-2015** | 50.9 | 129 | 133.1 | 90.2 | 52.3 | 19.4 | 5.9 |
| **2015-2020** | 43.6 | 127 | 131.6 | 85.3 | 47 | 16.6 | 4.8 |
| **2020-2024** | 37.2 | 125.7 | 130.8 | 81.3 | 42.4 | 14.1 | 3.8 |
| **2025-2029** | 31.4 | 124.8 | 130.4 | 77.8 | 38.3 | 11.9 | 2.9 |
| **2030-2034** | 26.2 | 124.5 | 130.4 | 74.8 | 34.6 | 9.8 | 2.1 |
| **2035-2039** | 21.5 | 124.8 | 131.3 | 72.5 | 31.4 | 7.9 | 1.4 |
| **2040-2044** | 17 | 125.7 | 132.6 | 70.6 | 28.5 | 6.2 | 0.6 |

**^¥^Source**: 2012 World Population Prospects

##### Table S5c. Age-specific population fertility rates in Zimbabwe 1950-2044^¥^

|  | **Age-specific fertility rates (births per 1,000 women)** | | | | | | |
| --- | --- | --- | --- | --- | --- | --- | --- |
| **Year** | **15-19** | **20-24** | **25-29** | **30-34** | **35-39** | **40-44** | **45-49** |
| **1950-1955** | 159.9 | 296.1 | 289.3 | 261.5 | 188.6 | 124.2 | 40.4 |
| **1955-1960** | 164.6 | 304.8 | 297.8 | 269.2 | 194.2 | 127.8 | 41.6 |
| **1960-1965** | 171.7 | 317.8 | 310.5 | 280.8 | 202.5 | 133.3 | 43.4 |
| **1965-1970** | 174.0 | 322.2 | 314.8 | 284.6 | 205.3 | 135.1 | 44.0 |
| **1970-1975** | 174.0 | 322.2 | 314.8 | 284.6 | 205.3 | 135.1 | 44.0 |
| **1975-1980** | 171.7 | 317.8 | 310.5 | 280.8 | 202.5 | 133.3 | 43.4 |
| **1980-1985** | 127.9 | 280.5 | 280.7 | 249.2 | 188.7 | 98.5 | 34.9 |
| **1985-1990** | 110.6 | 241.6 | 240.0 | 214.4 | 161.6 | 81.1 | 25.2 |
| **1990-1995** | 102.7 | 207.0 | 195.0 | 171.9 | 126.7 | 60.5 | 19.0 |
| **1995-2000** | 100.1 | 190.7 | 178.9 | 143.9 | 103.2 | 46.1 | 14.1 |
| **2000-2005** | 100.4 | 190.2 | 171.8 | 136.8 | 92.8 | 40.8 | 11.3 |
| **2005-2010** | 111.1 | 202.9 | 178.2 | 142.7 | 94.8 | 38.3 | 9.1 |
| **2010-2015** | 108.8 | 209.3 | 197.8 | 153.3 | 104.0 | 37.5 | 7.1 |
| **2015-2020** | 86.1 | 184.4 | 174.0 | 149.1 | 90.3 | 35.9 | 5.3 |
| **2020-2024** | 71.7 | 166.2 | 162.5 | 143.9 | 83.5 | 32.8 | 3.9 |
| **2025-2029** | 60.2 | 150.7 | 152.8 | 139.5 | 77.8 | 30.3 | 2.9 |
| **2030-2034** | 51.0 | 137.6 | 144.7 | 135.9 | 73.2 | 28.1 | 2.2 |
| **2035-2039** | 43.6 | 126.2 | 137.7 | 132.9 | 69.4 | 26.2 | 1.7 |
| **2040-2044** | 37.4 | 115.9 | 131.3 | 129.9 | 66.0 | 24.5 | 1.4 |

**^¥^Source**: 2019 World Population Prospects

##### HIV-deleted mortality rates calculations

In EMOD, we modelled HIV cause-deleted mortality rates in the background and HIV transmission and related morality rates in the foreground. To calculate the HIV deleted mortality, we first investigated all-cause mortality trends between 1960 and 2000 between countries with and without widespread HIV-AIDS crises. Countries without the epidemic demonstrated an exponential decline in mortality, while those grappling with the crisis experienced an exponential decrease interrupted by a sudden spike in the 1980s. We assumed the difference between these two curves (i.e., the spike) is due to the impact of the HIV-AIDS epidemic. Taking Kenya as an example, we fitted an exponential curve from 1970 to 1980 to represent the cause-deleted mortality. We then conducted a check on the population demographics generated by EMOD, ensuring that the age structure of the population simulated through both cause-deleted mortality and simulated HIV transmission aligns with the UN WPP’s population projections post-1980.

##### Table S6a. Age-specific HIV deleted mortality rates in Kenya 1950-2049 by gender ^¥^

|  |  | **Age-specific mortality rates (%)** | | | | | | |
| --- | --- | --- | --- | --- | --- | --- | --- | --- |
| **Sex** | **Year** | **15-19** | **20-24** | **25-29** | **30-34** | **35-39** | **40-44** | **45-49** |
| **Women** | **1997.5** | 0.136 | 0.191 | 0.244 | 0.294 | 0.361 | 0.453 | 0.542 |
|  | **2002.5** | 0.116 | 0.167 | 0.215 | 0.259 | 0.321 | 0.408 | 0.493 |
|  | **2007.5** | 0.100 | 0.145 | 0.188 | 0.229 | 0.286 | 0.367 | 0.449 |
|  | **2012.5** | 0.085 | 0.127 | 0.166 | 0.202 | 0.254 | 0.331 | 0.408 |
|  | **2017.5** | 0.073 | 0.110 | 0.145 | 0.178 | 0.226 | 0.298 | 0.371 |
|  | **2022.5** | 0.063 | 0.096 | 0.128 | 0.158 | 0.201 | 0.268 | 0.338 |
|  | **2027.5** | 0.054 | 0.084 | 0.112 | 0.139 | 0.179 | 0.241 | 0.307 |
|  | **2032.5** | 0.046 | 0.073 | 0.099 | 0.123 | 0.160 | 0.217 | 0.280 |
|  | **2037.5** | 0.039 | 0.064 | 0.087 | 0.108 | 0.142 | 0.196 | 0.254 |
|  | **2042.5** | 0.034 | 0.055 | 0.076 | 0.096 | 0.126 | 0.176 | 0.231 |
| **Men** | **1997.5** | 0.165 | 0.253 | 0.288 | 0.345 | 0.433 | 0.551 | 0.713 |
|  | **2002.5** | 0.142 | 0.219 | 0.251 | 0.304 | 0.385 | 0.494 | 0.648 |
|  | **2007.5** | 0.121 | 0.189 | 0.219 | 0.267 | 0.343 | 0.443 | 0.589 |
|  | **2012.5** | 0.104 | 0.163 | 0.191 | 0.235 | 0.305 | 0.398 | 0.535 |
|  | **2017.5** | 0.089 | 0.141 | 0.166 | 0.207 | 0.271 | 0.357 | 0.486 |
|  | **2022.5** | 0.076 | 0.122 | 0.145 | 0.182 | 0.241 | 0.320 | 0.441 |
|  | **2027.5** | 0.065 | 0.106 | 0.126 | 0.160 | 0.215 | 0.287 | 0.401 |
|  | **2032.5** | 0.056 | 0.091 | 0.110 | 0.141 | 0.191 | 0.258 | 0.364 |
|  | **2037.5** | 0.048 | 0.079 | 0.096 | 0.124 | 0.170 | 0.231 | 0.331 |
|  | **2042.5** | 0.041 | 0.068 | 0.083 | 0.109 | 0.151 | 0.208 | 0.300 |

**^¥^Source**: 2019 World Population Prospects

##### Table S6b. Age-specific mortality rates in South Africa 1950-2049 by gender^¥^

|  |  | **Age-specific mortality rates (%)** | | | | | | |
| --- | --- | --- | --- | --- | --- | --- | --- | --- |
| **Sex** | **Year** | **15-19** | **20-24** | **25-29** | **30-34** | **35-39** | **40-44** | **45-49** |
| **Women** | **1997.5** | 0.116 | 0.163 | 0.215 | 0.263 | 0.351 | 0.482 | 0.702 |
|  | **2002.5** | 0.096 | 0.134 | 0.179 | 0.222 | 0.302 | 0.424 | 0.629 |
|  | **2007.5** | 0.079 | 0.111 | 0.150 | 0.187 | 0.260 | 0.373 | 0.563 |
|  | **2012.5** | 0.066 | 0.092 | 0.125 | 0.158 | 0.223 | 0.328 | 0.504 |
|  | **2017.5** | 0.054 | 0.076 | 0.105 | 0.133 | 0.192 | 0.288 | 0.451 |
|  | **2022.5** | 0.045 | 0.063 | 0.087 | 0.113 | 0.165 | 0.254 | 0.404 |
|  | **2027.5** | 0.037 | 0.052 | 0.073 | 0.095 | 0.142 | 0.223 | 0.362 |
|  | **2032.5** | 0.031 | 0.043 | 0.061 | 0.080 | 0.122 | 0.196 | 0.324 |
|  | **2037.5** | 0.025 | 0.035 | 0.051 | 0.068 | 0.105 | 0.172 | 0.290 |
|  | **2042.5** | 0.021 | 0.029 | 0.042 | 0.057 | 0.090 | 0.152 | 0.260 |
| **Men** | **1997.5** | 0.177 | 0.312 | 0.361 | 0.437 | 0.594 | 0.872 | 1.264 |
|  | **2002.5** | 0.169 | 0.281 | 0.327 | 0.395 | 0.541 | 0.803 | 1.177 |
|  | **2007.5** | 0.173 | 0.254 | 0.296 | 0.358 | 0.494 | 0.739 | 1.095 |
|  | **2012.5** | 0.156 | 0.230 | 0.268 | 0.324 | 0.451 | 0.681 | 1.019 |
|  | **2017.5** | 0.141 | 0.207 | 0.243 | 0.294 | 0.411 | 0.627 | 0.949 |
|  | **2022.5** | 0.127 | 0.187 | 0.220 | 0.266 | 0.375 | 0.578 | 0.883 |
|  | **2027.5** | 0.115 | 0.169 | 0.199 | 0.241 | 0.342 | 0.532 | 0.822 |
|  | **2032.5** | 0.103 | 0.153 | 0.180 | 0.218 | 0.312 | 0.490 | 0.765 |
|  | **2037.5** | 0.093 | 0.138 | 0.163 | 0.197 | 0.285 | 0.451 | 0.713 |
|  | **2042.5** | 0.082 | 0.124 | 0.148 | 0.179 | 0.260 | 0.415 | 0.663 |

**^¥^Source**: 2012 World Population Prospects

##### Table S6C. Age-specific HIV-deleted mortality rates in Zimbabwe 1950-2049 by gender

|  |  | **Age-specific mortality rates (%)** | | | | | | |
| --- | --- | --- | --- | --- | --- | --- | --- | --- |
| **Sex** | **Year** | **15-19** | **20-24** | **25-29** | **30-34** | **35-39** | **40-44** | **45-49** |
| **Women** | **1997.5** | 0.130 | 0.205 | 0.293 | 0.369 | 0.470 | 0.580 | 0.692 |
|  | **2002.5** | 0.104 | 0.172 | 0.251 | 0.321 | 0.416 | 0.521 | 0.631 |
|  | **2007.5** | 0.084 | 0.144 | 0.216 | 0.279 | 0.368 | 0.467 | 0.574 |
|  | **2012.5** | 0.068 | 0.121 | 0.186 | 0.243 | 0.325 | 0.420 | 0.523 |
|  | **2017.5** | 0.055 | 0.102 | 0.159 | 0.212 | 0.288 | 0.377 | 0.477 |
|  | **2022.5** | 0.044 | 0.085 | 0.137 | 0.184 | 0.254 | 0.338 | 0.434 |
|  | **2027.5** | 0.035 | 0.071 | 0.118 | 0.160 | 0.225 | 0.303 | 0.395 |
|  | **2032.5** | 0.029 | 0.060 | 0.101 | 0.140 | 0.199 | 0.272 | 0.360 |
|  | **2037.5** | 0.023 | 0.050 | 0.087 | 0.121 | 0.176 | 0.244 | 0.328 |
|  | **2042.5** | 0.018 | 0.042 | 0.075 | 0.106 | 0.156 | 0.219 | 0.299 |
| **Men** | **1997.5** | 0.185 | 0.285 | 0.333 | 0.407 | 0.521 | 0.666 | 0.869 |
|  | **2002.5** | 0.154 | 0.240 | 0.283 | 0.351 | 0.456 | 0.590 | 0.784 |
|  | **2007.5** | 0.128 | 0.203 | 0.241 | 0.302 | 0.399 | 0.522 | 0.708 |
|  | **2012.5** | 0.107 | 0.171 | 0.205 | 0.261 | 0.349 | 0.463 | 0.638 |
|  | **2017.5** | 0.089 | 0.144 | 0.174 | 0.225 | 0.305 | 0.410 | 0.576 |
|  | **2022.5** | 0.074 | 0.122 | 0.148 | 0.194 | 0.267 | 0.363 | 0.520 |
|  | **2027.5** | 0.062 | 0.103 | 0.126 | 0.167 | 0.234 | 0.321 | 0.469 |
|  | **2032.5** | 0.052 | 0.087 | 0.107 | 0.144 | 0.204 | 0.285 | 0.423 |
|  | **2037.5** | 0.043 | 0.073 | 0.091 | 0.124 | 0.179 | 0.252 | 0.381 |
|  | **2042.5** | 0.036 | 0.062 | 0.078 | 0.107 | 0.156 | 0.223 | 0.344 |

**^¥^Source**: 2019 World Population Prospects

#### Model fit to age-specific and overall prevalence from population-based surveys by sex

##### Figure S1a Model fit to age-specific and overall prevalence from population-based surveys by sex in Kenya

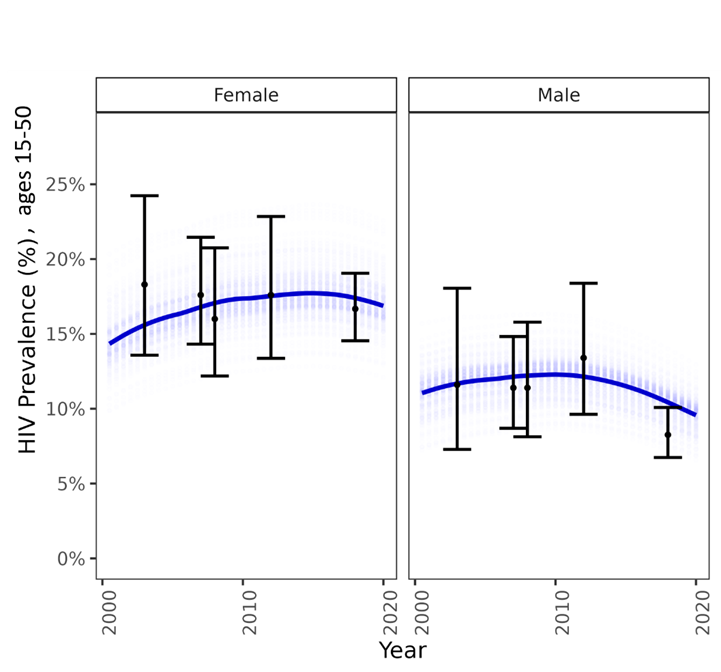

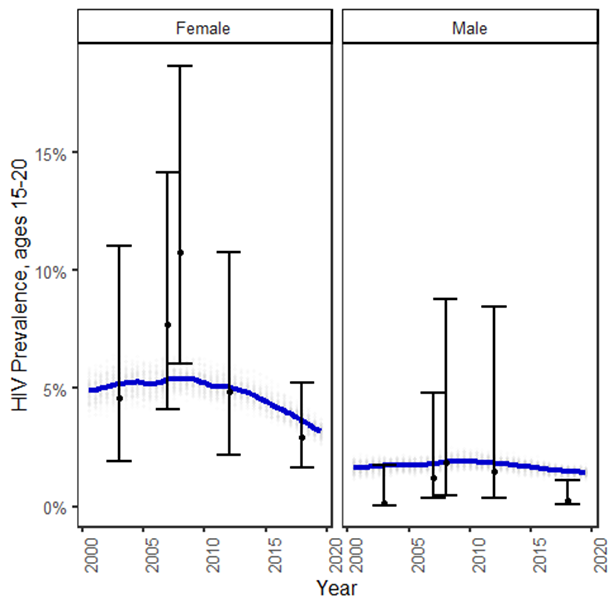

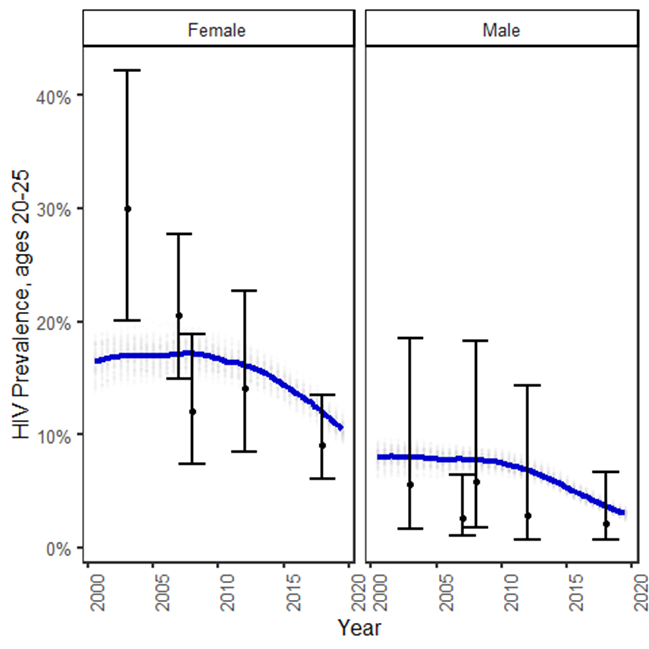

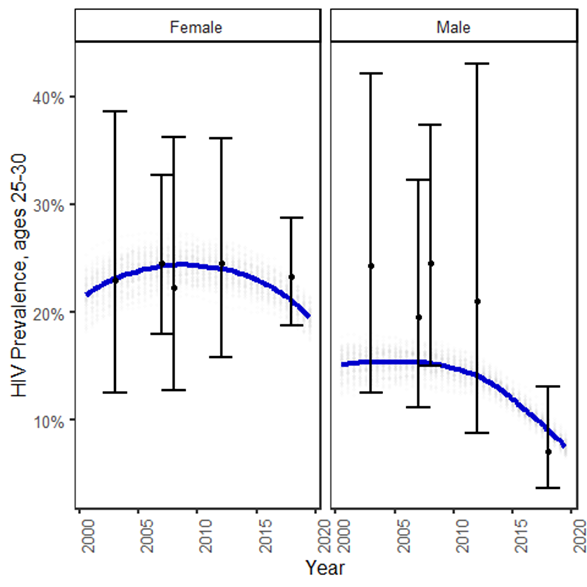

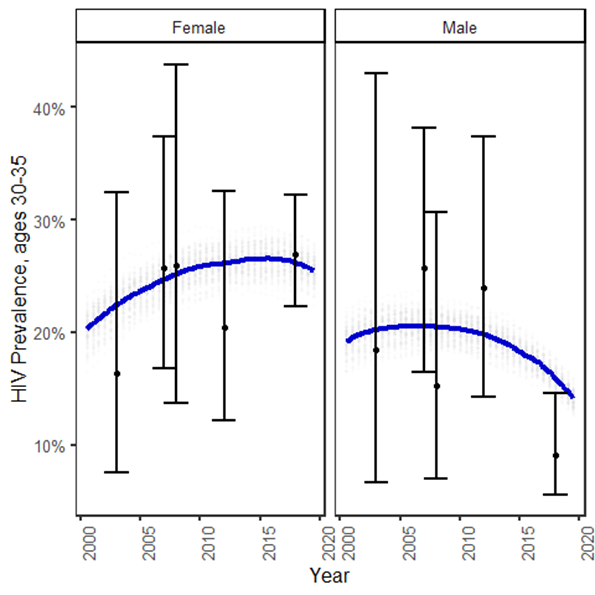

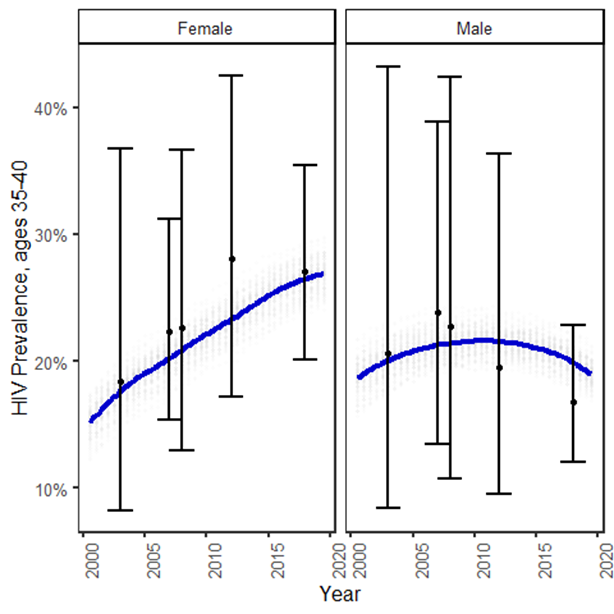

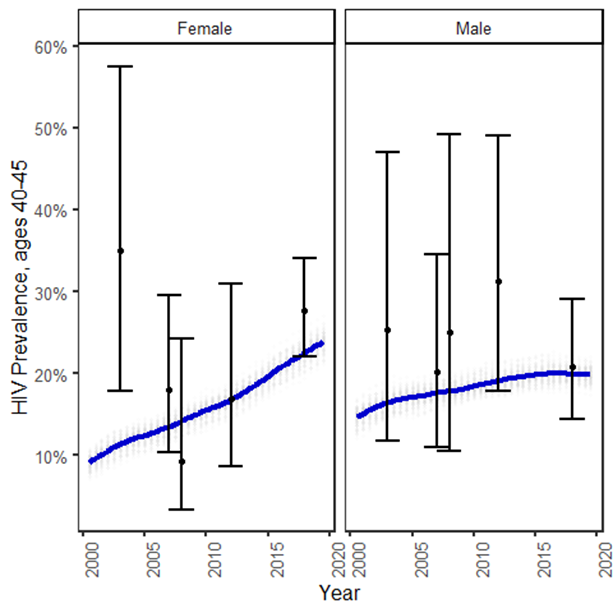

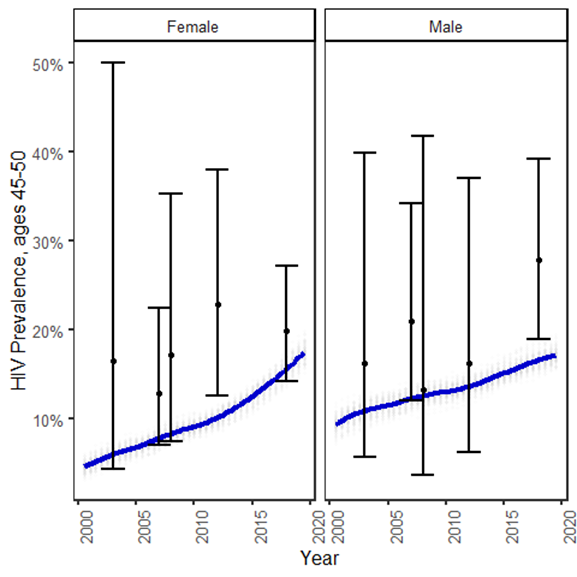

* Blue curves refer to the LOESS line fitting to the 100 simulations; the error bars refer to the empirical estimates and 95% confidence intervals for HIV prevalence obtained from Kenya Demographic and Health Surveys and Kenya AIDS Indicator Surveys.

##### Figure S1b Model fit to age-specific and overall prevalence from population-based surveys by sex in South Africa

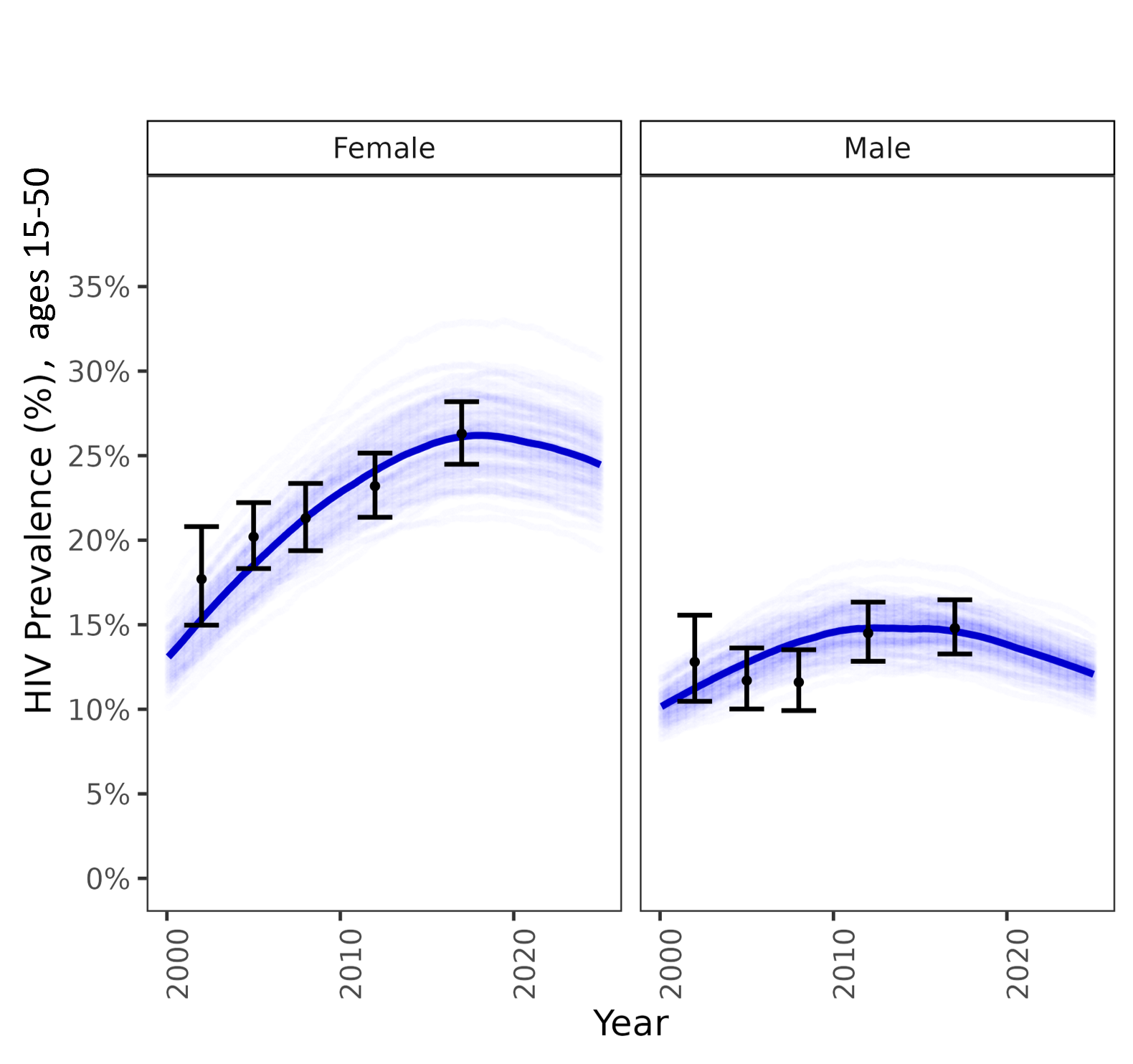

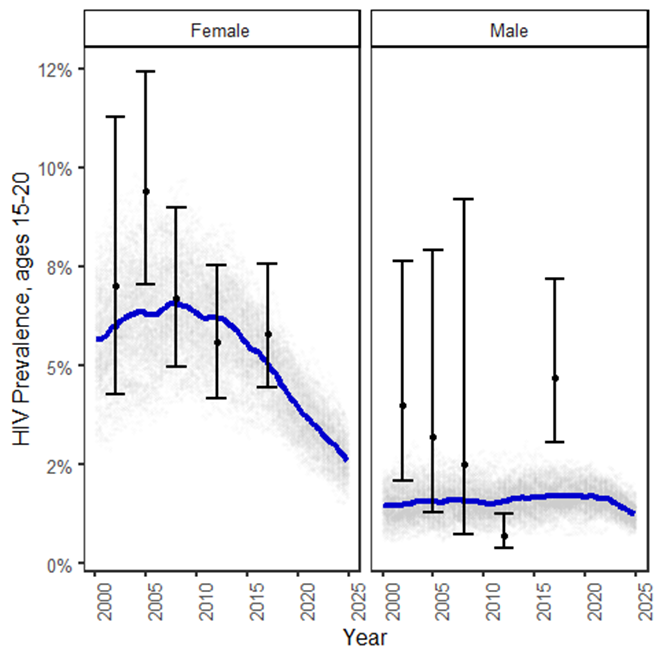

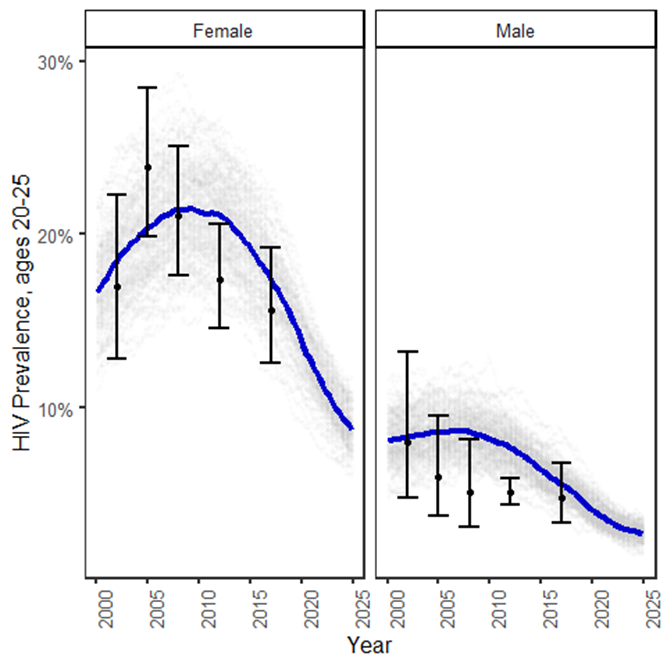

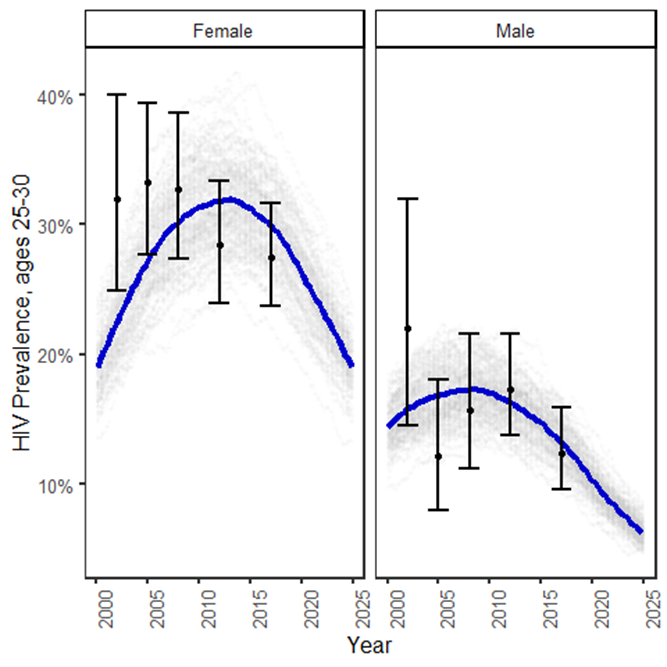

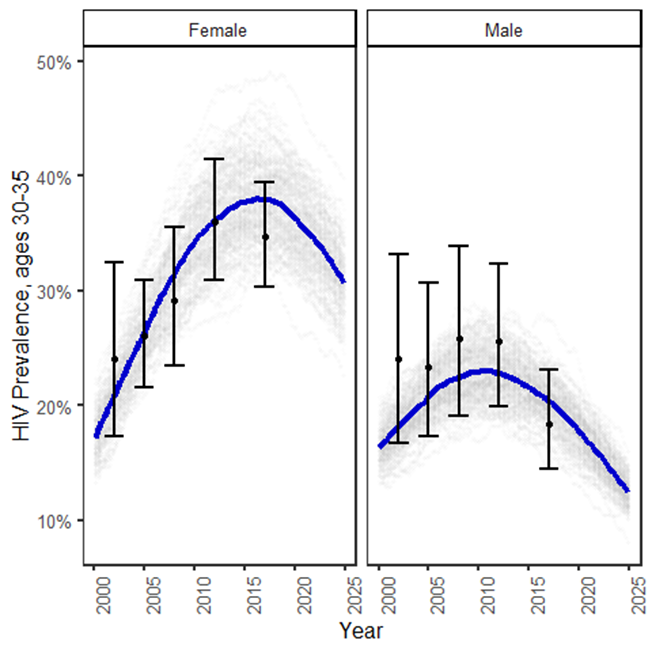

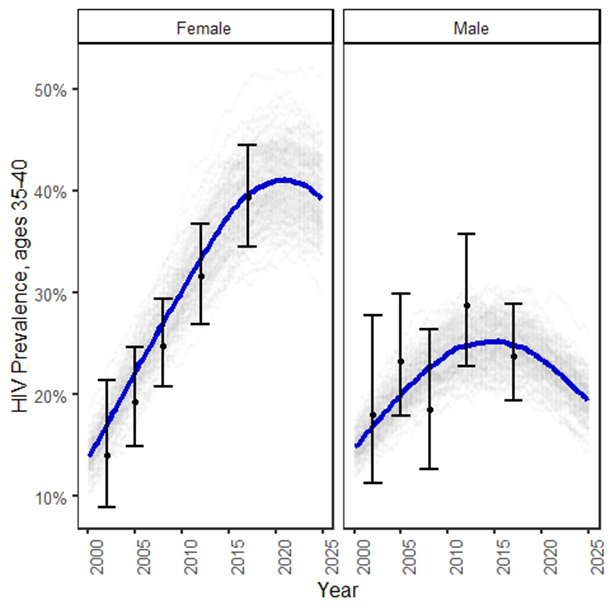

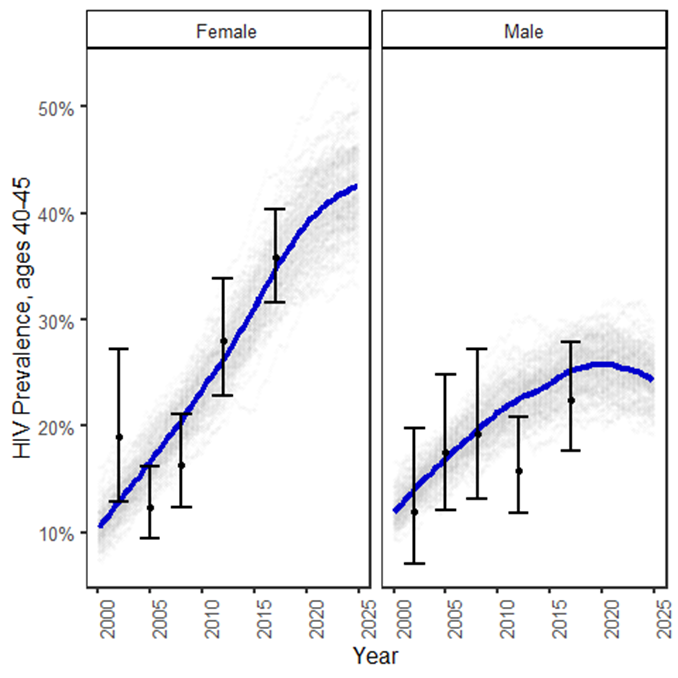

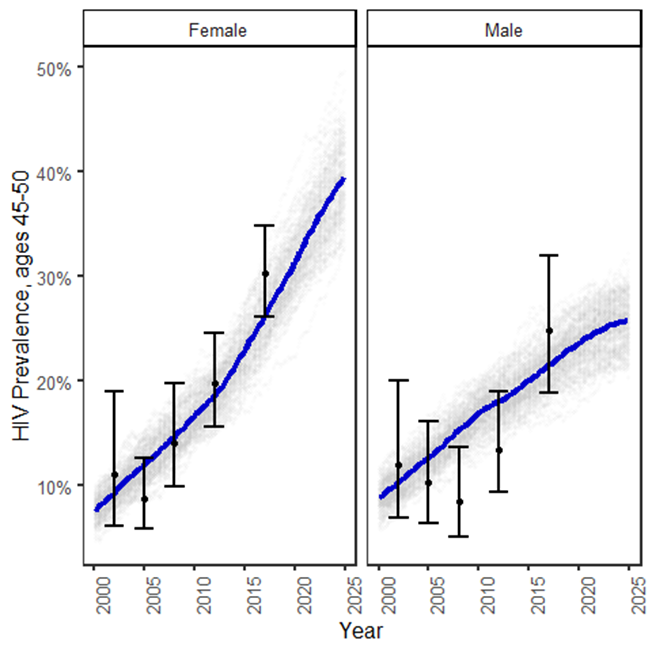

* Blue curves refer to the LOESS average fitting to the 100 simulations; the error bars refer to the empirical estimates and 95% confidence intervals for HIV prevalence obtained from South African National HIV Prevalence, Incidence and Behaviour Surveys (2002, 2005, 2008, 2012 and 2017) from the Human Sciences Research Council (HSRC)

##### Figure S1c. Model fit to age-specific and overall prevalence from population-based surveys by sex in Zimbabwe

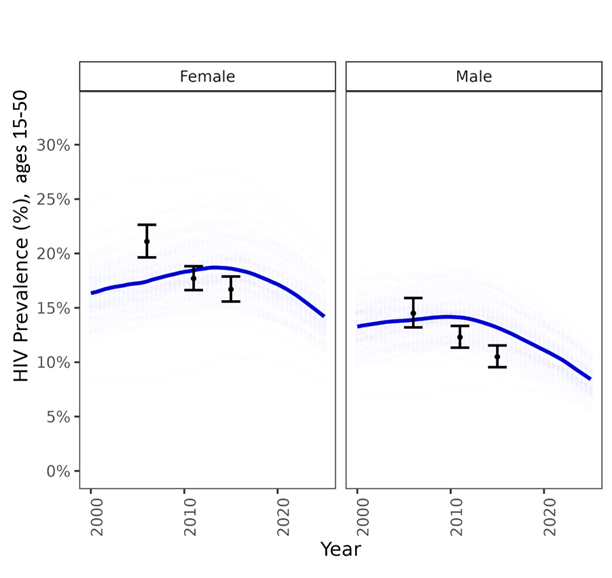

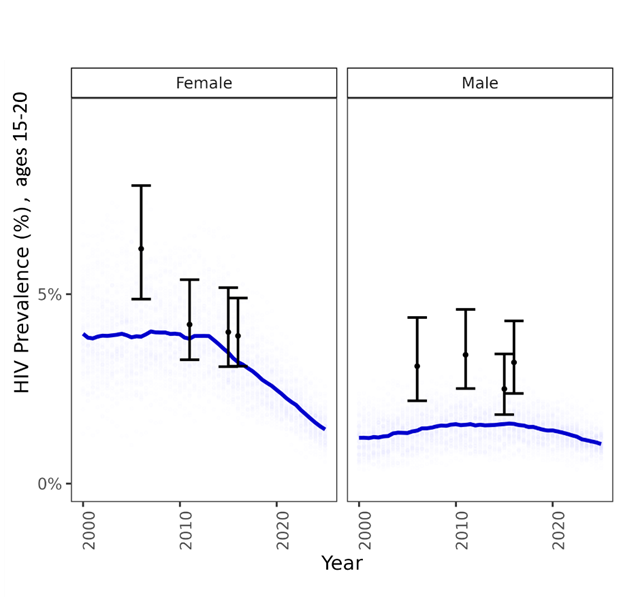

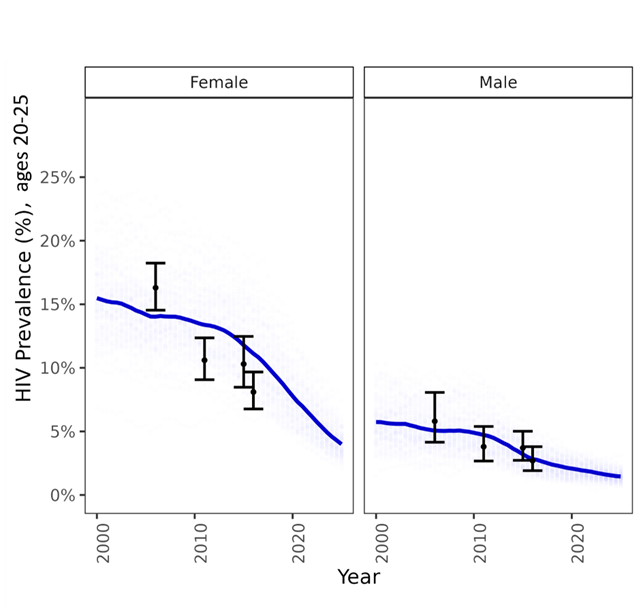

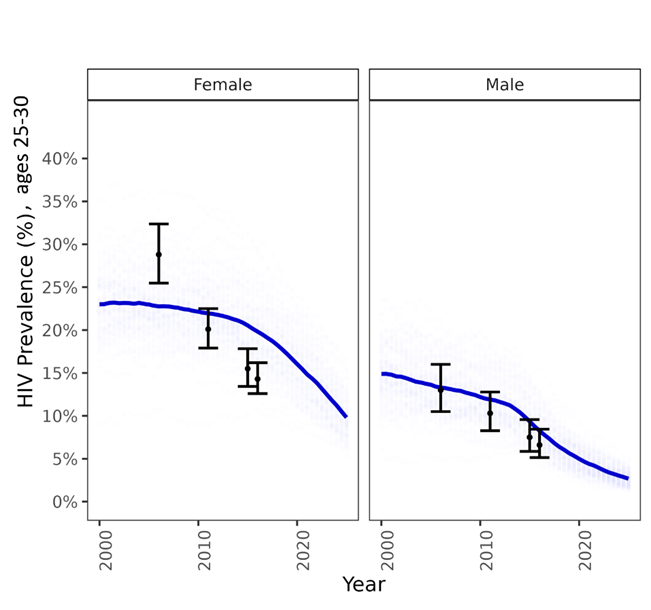

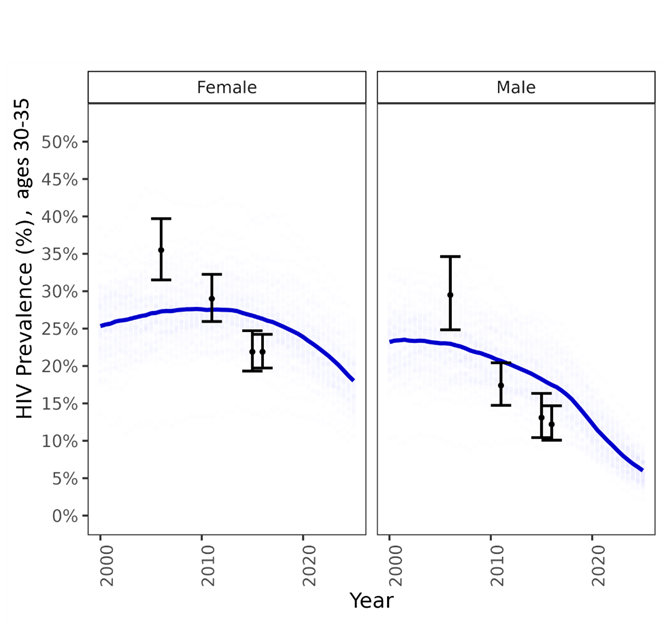

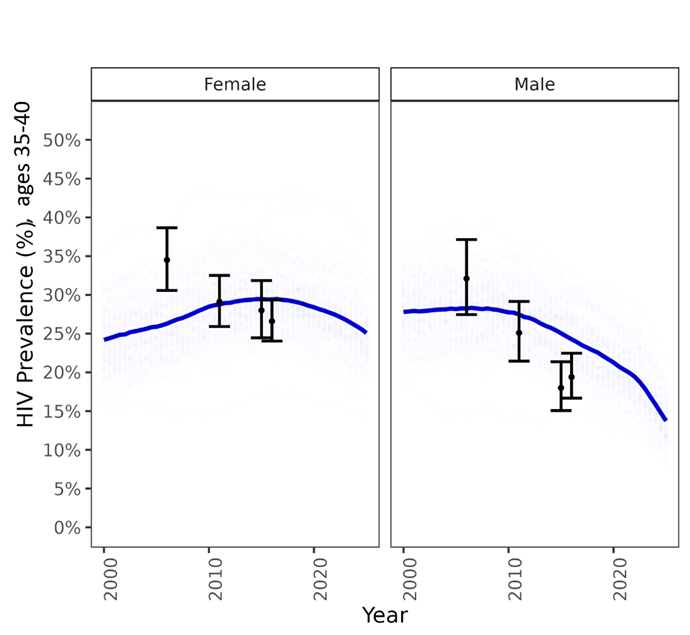

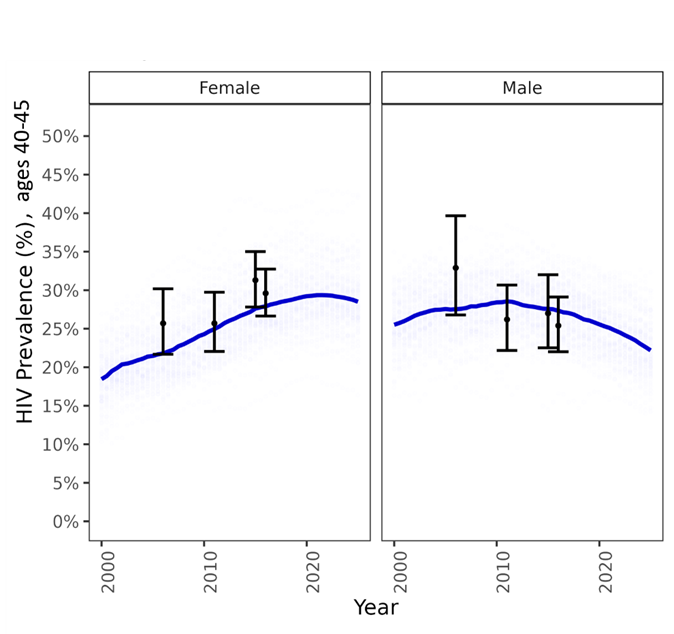

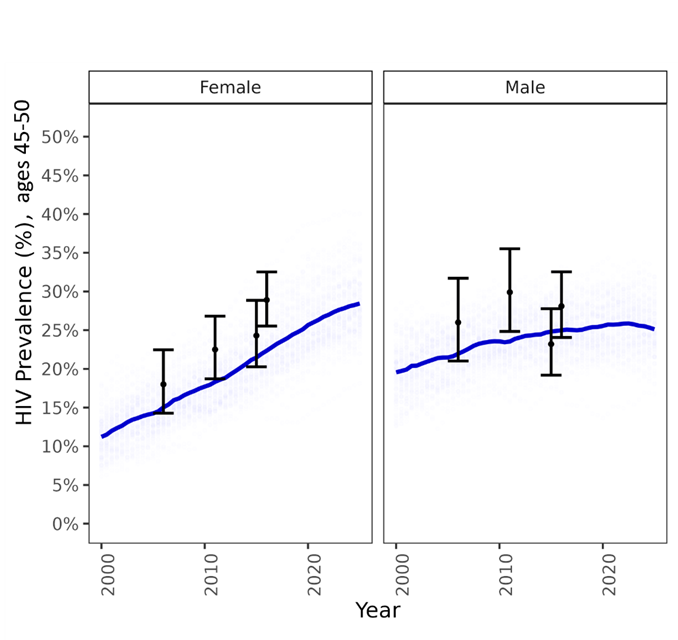

* Blue curves refer to the LOESS average fitting to the 100 simulations; the error bars refer to the empirical estimates and 95% confidence intervals for HIV prevalence obtained from PHIA Surveys

#### Model fit to age-specific and overall prevalence from population-based surveys by sex

##### Figure S2a Model fit to age-specific and overall ART coverage from population-based surveys by sex in Kenya

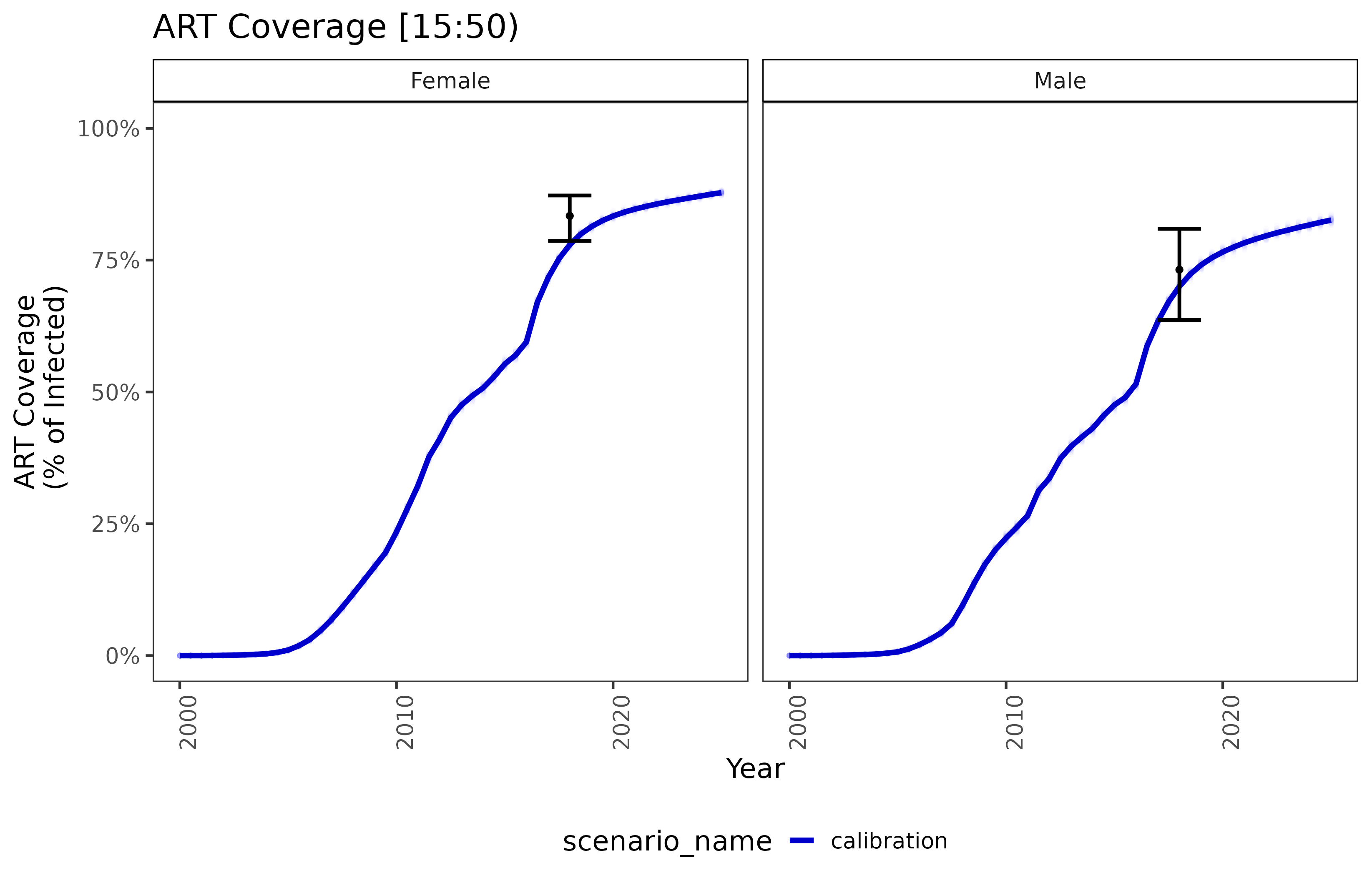

##### Figure S2b Model fit to age-specific and overall ART coverage from population-based surveys by sex in South Africa

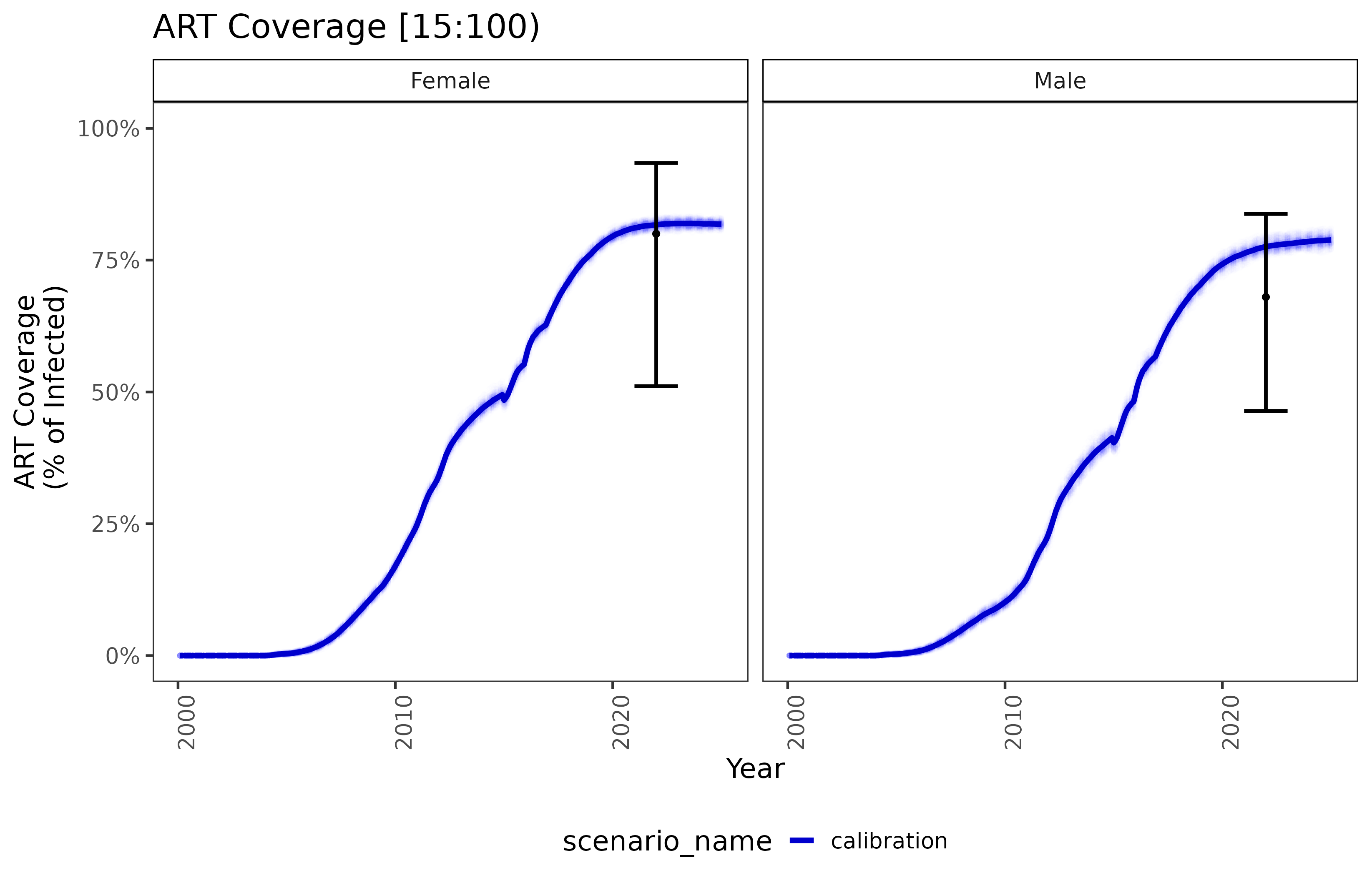

##### Figure S2c Model fit to age-specific and overall ART coverage from population-based surveys by sex in Zimbabwe

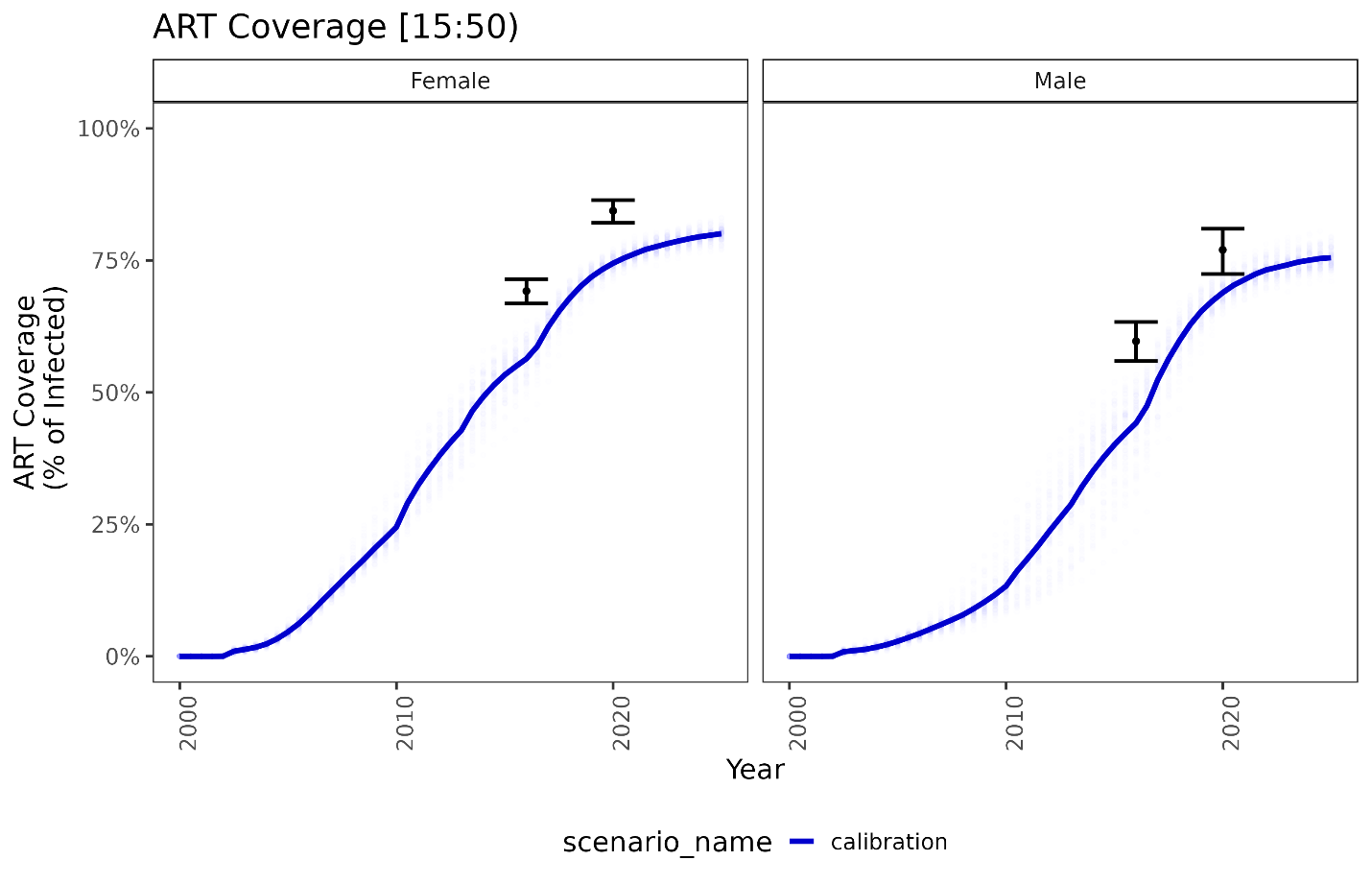

### Model Overview and Parameters

*Model initialization*

The simulations begin prior to the start of infection in the year 1960 to allow sufficient time for the epidemic to burn-in. During this time, individuals with demographic properties specified in the demographics file begin forming relationships; relationship formation rates for each gender and relationship type are updated daily using a relationship flow algorithm. Adjustment of pair formation entry rates is terminated at a specific timepoint (e.g., 1975) and the rates are fixed at that value for the remainder of the simulation. The age profile of the population is initialized using demographic data including population age distribution or age-specific fertility rates. A prior analysis of our model evaluated the age/sex pairings, partnership length and other sexual network characteristics and confirmed that these outputs reached equilibrium within 20 years, prior to the introduction of HIV into the model: <https://ieeexplore.ieee.org/abstract/document/6426573>. (25) HIV infections are seeded in 1980 and affect a certain proportion of the population based on the age and gender distribution reported from historical data. ART intervention is introduced in the year 2012. Eligible individuals enroll in ART based on historical eligibility criteria based on CD4 count, which changes over time based on WHO guidelines for ART initiation until the implementation of universal ART, which is assumed to remain the same until the end of the simulations (year 2050). Calibration and validation processes are performed to refine the initialization and ensure that the model aligns with observed HIV dynamics in the target population (26,27).

The following provides a more detailed description of the model initialization process: <https://docs.idmod.org/projects/emod-hiv/en/latest/sti-model-relationships.html>. (26)

*Modelled time step*

The model is implemented using a monthly time step, aggregating events and changes over the course of each month. Monthly updated information can then be ascertained regarding a range of activities and occurrences, including sexual mixing, relationship formation, stages of HIV infection, HIV testing and its results, and PrEP initiation and discontinuation.

*PrEP cascade*

HIV-negative individuals aged 18 to 49 years were eligible for PrEP if testing negative at the time of initiation or continuation, and if sexually active with at least one partner. PrEP discontinuation occurred if individuals are lost to follow-up or no longer met the eligibility criteria (i.e., an individual turned 50 years old, all partnerships end, or tested HIV-positive). Individuals who stopped PrEP can re-start at any time if they meet the eligibility criteria (i.e. start a new partnership).

#### Model Parameters

##### Table S7a. Select model parameters used to fit the EMOD-HIV transmission model to survey data on prevalence and ART coverage from Kenya.

| **Parameter** | **Parameter Description** | **Fitted median** | **(IQR)** |
| --- | --- | --- | --- |
| ARTLinkMax | Maximum probability of linkage to ART | 0.999 | (0.978, 1.000) |
| ARTLinkMid | Year of ART linkage (given eligibility), that is, time of the inflection point in the sigmoid trend. | 2003.008 | (2002.920, 2003.837) |
| AcuteDurationMonths | The time since infection, in months, over which the Acute_Stage_Infectivity_Multiplier is applied to coital acts occurring in that time period. | 1.000 | (1.000, 1.331) |
| CircumcisionReducedAcquire | The reduction of susceptibility to STI by voluntary male medical circumcision (VMMC). | 0.600 | (0.598, 0.600) |
| Homa_BayInfrmlCondomsMax | Maximum rate of condom use in informal relationships in Homa Bay | 0.231 | (0.230, 0.233) |
| Homa_BayLOWRisk | Proportion of the population that is low-risk in Homa Bay | 0.557 | (0.549, 0.559) |
| Homa_BayTrnsCondomsMax | Maximum rate of condom use in transitory relationships in Homa Bay | 0.244 | (0.232, 0.245) |
| InfmrlFormRate | Informal relationship formation rate | 0.000 | (0.000, 0.000) |
| InfrmlCondomMid | Year midpoint of logistic scale-up of condom use in informal relationships | 1993.007 | (1991.629, 1998.680) |
| InfrmlCondomRate | Rate of logistic scale-up of condom use in informal relationships | 2.661 | (1.953, 2.909) |
| InfrmlCondomsMax | Maximum rate of condom use in informal relationships | 0.216 | (0.215, 0.217) |
| InfrmlDurHet | Heterogeneity in duration of informal relationships | 0.750 | (0.750, 0.750) |
| KisiiInfrmlCondomsMax | Maximum rate of condom use in informal relationships in Kisii | 0.231 | (0.226, 0.232) |
| KisiiLOWRisk | Proportion of the population that is low-risk in Kisii | 0.938 | (0.935, 0.939) |
| KisiiTrnsCondomsMax | Maximum rate of condom use in transitory relationships in Kisii | 0.375 | (0.369, 0.376) |
| KisumuInfrmlCondomsMax | Maximum rate of condom use in informal relationships in Kisumu | 0.182 | (0.181, 0.183) |
| KisumuLOWRisk | Proportion of the population that is low-risk in Kisumu | 0.764 | (0.762, 0.765) |
| KisumuTrnsCondomsMax | Maximum rate of condom use in transitory relationships in Kisumu | 0.338 | (0.337, 0.347) |
| LogBaseInfectivity | The probability of transmission when none of the transmission multipliers apply to a particular coital act. | 0.002 | (0.002, 0.002) |
| MaleToFemaleOld | Male-to-female relative risk of infection among older individuals | 2.012 | (1.506, 2.132) |
| MaleToFemaleYoung | Male-to-female relative risk of infection among young individuals | 1.116 | (1.000, 1.242) |
| MaxInfrmlFLOW | Maximum number of informal relationships among low-risk females | 1.311 | (1.255, 1.315) |
| MaxInfrmlFMED | Maximum number of informal relationships among medium-risk females | 2.050 | (2.002, 2.351) |
| MaxInfrmlMLOW | Maximum number of informal relationships among low-risk males | 1.165 | (1.159, 1.203) |
| MaxInfrmlMMED | Maximum number of informal relationships among medium-risk males | 2.530 | (2.442, 2.735) |
| MaxMrtlFMED | Maximum number of marital relationship among medium-risk females | 1.139 | (1.136, 1.159) |
| MaxMrtlMMED | Maximum number of marital relationship among medium-risk males | 1.302 | (1.268, 1.316) |
| MaxTrnsFLOW | Maximum number of transitory relationships among low-risk females | 1.599 | (1.580, 1.625) |
| MaxTrnsFMED | Maximum number of transitory relationships among medium-risk females | 2.943 | (2.891, 3.000) |
| MaxTrnsMLOW | Maximum number of transitory relationships among low-risk males | 1.599 | (1.588, 1.677) |
| MaxTrnsMMED | Maximum number of transitory relationships among medium-risk males | 2.557 | (2.475, 2.879) |
| MigoriInfrmlCondomsMax | Maximum rate of condom use in informal relationships in Migori | 0.186 | (0.185, 0.189) |
| MigoriLOWRisk | Proportion of the population that is low-risk in Migori | 0.795 | (0.793, 0.797) |
| MigoriTrnsCondomsMax | Maximum rate of condom use in transitory relationships in Migori | 0.249 | (0.248, 0.257) |
| MrtlCondomMax | Maximum rate of condom use in marital relationships | 0.192 | (0.191, 0.193) |
| MrtlCondomMid | Year midpoint of logistic scale-up of condom use in marital relationships | 2003.131 | (1999.017, 2005.000) |
| MrtlCondomRate | Rate of logistic scale-up of condom use in marital relationships | 2.971 | (2.595, 3.000) |
| MrtlFormRate | Marital relationship formation rate | 0.000 | (0.000, 0.000) |
| NyamiraInfrmlCondomsMax | Maximum rate of condom use in informal relationships in Nyamira | 0.096 | (0.093, 0.097) |
| NyamiraLOWRisk | Proportion of the population that is low-risk in Nyamira | 0.902 | (0.900, 0.910) |
| NyamiraTrnsCondomsMax | Maximum rate of condom use in transitory relationships in Nyamira | 0.309 | (0.304, 0.313) |
| PrExInfrmlFemLOW | Probability of potential for extra-relational informal relationship among low-risk females | 0.366 | (0.364, 0.371) |
| PrExInfrmlFemMED | Probability of potential for extra-relational informal relationship among medium-risk females | 0.401 | (0.401, 0.407) |
| PrExInfrmlMaleLOW | Probability of potential for extra-relational informal relationship among low-risk males | 0.244 | (0.222, 0.249) |
| PrExInfrmlMaleMED | Probability of potential for extra-relational informal relationship among medium-risk males | 0.382 | (0.379, 0.389) |
| PrExTrnsFemLOW | Probability of potential for extra-relational transitory relationship among low-risk females | 0.033 | (0.033, 0.033) |
| PrExTrnsFemMED | Probability of potential for extra-relational transitory relationship among medium-risk females | 0.460 | (0.450, 0.468) |
| PrExTrnsMaleLOW | Probability of potential for extra-relational transitory relationship among low-risk males | 0.330 | (0.328, 0.332) |
| PrExTrnsMaleMED | Probability of potential for extra-relational transitory relationship among medium-risk males | 0.593 | (0.589, 0.611) |
| PreARTLinkMax | Maximum probability of linkage to pre-ART care | 0.715 | (0.708, 0.737) |
| PreARTLinkMid | Year midpoint of logistic scale-up of pre-ART linkage | 1996.709 | (1995.685, 1998.896) |
| PreARTLinkMin | Minimum probability of linkage to pre-ART care | 0.435 | (0.416, 0.442) |
| RiskAssortivity | Risk assortivity | 0.663 | (0.648, 0.673) |
| SeedYrHigh | Seed year | 1986.562 | (1982.000, 1988.000) |
| SexualDebutAgeFemaleWeibullHeterogeneity | Heterogeneity parameter of Weibull distribution of female age of sexual debut | 0.086 | (0.083, 0.086) |
| SexualDebutAgeFemaleWeibullScale | Scale parameter of Weibull distribution of female age of sexual debut | 16.013 | (15.540, 16.038) |
| SexualDebutAgeMaleWeibullHeterogeneity | Heterogeneity parameter of Weibull distribution of male age of sexual debut | 0.040 | (0.040, 0.040) |
| SexualDebutAgeMaleWeibullScale | Scale parameter of Weibull distribution of male age of sexual debut | 15.708 | (15.155, 16.090) |
| SiayaInfrmlCondomsMax | Maximum rate of condom use in informal relationships in Siaya | 0.160 | (0.148, 0.161) |
| SiayaLOWRisk | Proportion of the population that is low-risk in Siaya | 0.729 | (0.722, 0.731) |
| SiayaTrnsCondomsMax | Maximum rate of condom use in transitory relationships in Siaya | 0.300 | (0.293, 0.304) |
| TrnsCondomMax | Maximum rate of condom use in transitory relationships | 0.243 | (0.242, 0.253) |
| TrnsCondomMid | Year midpoint of logistic scale-up of condom use in transitory relationships | 1997.960 | (1996.962, 1999.273) |
| TrnsCondomRate | Rate of logistic scale-up of condom use in transitory relationships | 0.999 | (0.978, 1.000) |
| TrnsFormRate | Transitory relationship formation rate | 2003.008 | (2002.920, 2003.837) |

* Median and interquartile ranges (IQRs) reported for all dynamic parameters used in the calibration process from 100 best-fitting parameter sets. †

##### Table S7b. Select model parameters used to fit the EMOD-HIV transmission model to survey data on prevalence and ART coverage from South Africa.

| **Parameter** | **Description** | **Fitted Median** | **(IQR)** |
| --- | --- | --- | --- |
| ART Link Max | Maximum probability of linkage to ART | 1.000 | (0.997, 1.000) |
| ART Link Mid | Year of ART linkage (given eligibility), that is, time of the inflection point in the sigmoid trend. | 2,005.96 | (2,005.86, 2,006.08) |
| All: Infmrl Condom | Modern condom usage rate in informal relationships across all locations | 0.61 | (0.58, 0.64) |
| All: LOW Risk | Proportion of the population that is low-risk | 0.936 | (0.930, 0.942) |
| All: Trns Condom | Modern condom usage rate in transitory relationships across all locations | 0.2 | (0.16, 0.23) |
| Base Infectivity | The probability of transmission when none of the transmission multipliers apply to a coital act (or when all multipliers are set to 1). | 0.0015 | (0.0015, 0.0016) |
| Circumcision Reduced Acquire | The reduction of susceptibility to STI by voluntary male medical circumcision (VMMC). | 0.6 |  |
| Infmrl Condoms Late | Modern condom usage rate in informal relationships, by county | 0.37 | (0.35, 0.39) |
| Infmrl Form Rate | Informal relationship formation rate | 0.0009 | (0.0008, 0.0009) |
| Infrml Condom Mid | Year midpoint of logistic scale-up of condom use in informal relationships | 1,998.27 | (1,997.81, 1,999.03) |
| Infrml Condom Rate | Rate of logistic scale-up of condom use in informal relationships | 2.03 | (1.89, 2.18) |
| Infrml Dur Het | Heterogeneity in duration of informal relationships | 0.693 | (0.675, 0.711) |
| Male To Female Old | Male-to-female relative risk of infection among older individuals | 2.33 | (2.22, 2.49) |
| Male To Female Young | Male-to-female relative risk of infection among young individuals | 2.97 | (2.74, 3.37) |
| Max Infmrl F LOW | Maximum number of informal relationships among low-risk females | 1.67 | (1.61, 1.72) |
| Max Infmrl F MED | Maximum number of informal relationships among medium-risk females | 0.92 | (0.89, 0.94) |
| Max Infmrl M LOW | Maximum number of informal relationships among low-risk males | 1.73 | (1.69, 1.76) |
| Max Infmrl M MED | Maximum number of informal relationships among medium-risk males | 0.75 | (0.70, 0.79) |
| Max Mrtl F ME | Maximum number of marital relationships among medium-risk females | 1.23 | (1.16, 1.27) |
| Max Mrtl M ME | Maximum number of marital relationships among medium-risk males | 0.93 | (0.91, 0.96) |
| Max Trns F LOW | Maximum number of transitory relationships among low-risk females | 1.5 | (1.48, 1.52) |
| Max Trns F MED | Maximum number of transitory relationships among medium-risk females | 3.1 | (3.06, 3.14) |
| Max Trns M LOW | Maximum number of transitory relationships among low-risk males | 1.49 | (1.46, 1.53) |
| Max Trns M MED | Maximum number of transitory relationships among medium-risk males | 3.02 | (2.88, 3.10) |
| Mrtl Condom Max | Maximum rate of condom use in marital relationships | 0.191 | (0.184, 0.211) |
| Mrtl Condom Mid | Year midpoint of logistic scale-up of condom use in marital relationships | 1,994.92 | (1,994.19, 1,995.27) |
| Mrtl Condom Rate | Rate of logistic scale-up of condom use in marital relationships | 3.6 | (3.49, 3.69) |
| Mrtl Form Rate | Marital relationship formation rate | 0.0001 | (0.0001, 0.0001) |
| Pr Ex Infmrl Fem LOW | Probability of potential for extra-relational informal relationship among low-risk females | 0.08 | (0.06, 0.10) |
| Pr Ex Infmrl Fem MED | Probability of potential for extra-relational informal relationship among medium-risk females | 0.391 | (0.381, 0.404) |
| Pr Ex Infmrl Male LOW | Probability of potential for extra-relational informal relationship among low-risk males | 0.46 | (0.42, 0.50) |
| Pr Ex Infmrl Male MED | Probability of potential for extra-relational informal relationship among medium-risk males | 0.379 | (0.370, 0.390) |
| Pr Ex Trns Fem LOW | Probability of potential for extra-relational transitory relationship among low-risk females | 0.066 | (0.058, 0.081) |
| Pr Ex Trns Fem MED | Probability of potential for extra-relational transitory relationship among medium-risk females | 0.58 | (0.56, 0.62) |
| Pr Ex Trns Male LOW | Probability of potential for extra-relational transitory relationship among low-risk males | 0.17 | (0.12, 0.19) |
| Pr Ex Trns Male MED | Probability of potential for extra-relational transitory relationship among medium-risk males | 0.59 | (0.58, 0.62) |
| PreART Link Max | Maximum probability of linkage to pre-ART care | 0.89 | (0.85, 0.96) |
| PreART Link Mid | Year midpoint of logistic scale-up of pre-ART linkage | 1,998.28 | (1,997.76, 1,998.72) |
| PreART Link Min | Minimum probability of linkage to pre-ART care | 0.63 | (0.62, 0.65) |
| Risk Assortivity | Risk assortivity | 0.47 | (0.44, 0.49) |
| SeedYr HIGH | Seed year | 1991 |  |
| Sexual Debut Age Female Weibull Heterogeneity | Heterogeneity parameter of Weibull distribution of female age of sexual debut | 0.062 | (0.051, 0.067) |
| Sexual Debut Age Female Weibull Scale | Scale parameter of Weibull distribution of female age of sexual debut | 16.49 | (16.34, 16.62) |
| Sexual Debut Age Male Weibull Heterogeneity | Heterogeneity parameter of Weibull distribution of male age of sexual debut | 0.043 | (0.038, 0.050) |
| Sexual Debut Age Male Weibull Scale | Scale parameter of Weibull distribution of male age of sexual debut | 16.47 | (16.24, 16.62) |
| Trns Condom Late | Modern condom usage rate in transitory relationships | 0.61 | (0.59, 0.64) |
| Trns Condom Mid | Year midpoint of logistic scale-up of condom use in transitory relationships | 2,006.77 | (2,006.08, 2,007.21) |
| Trns Condom Rate | Rate of logistic scale-up of condom use in transitory relationships | 1.94 | (1.84, 2.01) |
| Trns Form Rate | Transitory relationship formation rate | 0.0013 | (0.0013, 0.0014) |

* Median and interquartile ranges (IQRs) reported for all dynamic parameters used in the calibration process from 100 best-fitting parameter sets. †

##### Table S7c. Select model parameters used to fit the EMOD-HIV transmission model to survey data on prevalence and ART coverage from Zimbabwe.

| **Parameter** | **Parameter Description** | **Fitted Median** | **(IQR)** |
| --- | --- | --- | --- |
| All: LOW Risk | Proportion of the population that is low-risk | 0.523 | (0.500, 0.540) |
| Base Infectivity | The probability of transmission when none of the transmission multipliers apply to a coital act | 0.0038 | (0.0035, 0.0040) |
| Demographic Coverage | Demographic coverage of the intervention | 0.193 | (0.185, 0.205) |
| HCT Uptake Post Debut Max | Maximum rate of HIV counseling and testing (HCT) post sexual debut | 0.34 | (0.28, 0.38) |
| HCT Uptake Post Debut Mid | Year midpoint of logistic scale-up of HIV counseling and testing (HCT) post sexual debut | 2,008.86 | (2,008.32, 2,009.70) |
| Informal Form Rate | Informal relationship formation rate | 0.0012 | (0.0011, 0.0013) |
| Infrml Condom Mid | Year midpoint of logistic scale-up of condom use in informal relationships | 1,998.13 | (1,997.41, 1,998.67) |
| Infrml Condom Rate | Rate of logistic scale-up of condom use in informal relationships | 3.85 | (3.73, 4.00) |
| Infrml Condoms Max | Maximum rate of condom use in informal relationships | 0.21 | (0.19, 0.26) |
| Male To Female Old | Male-to-female relative risk of infection among older individuals | 1.44 | (1.33, 1.66) |
| Male To Female Young | Male-to-female relative risk of infection among young individuals | 2.66 | (2.26, 2.83) |
| Marital Form Rate | Marital relationship formation rate | 0.00011 | (0.00009, 0.00014) |
| Max Infrml F LOW | Maximum number of informal relationships among low-risk females | 1.25 | (1.18, 1.32) |
| Max Infrml F MED | Maximum number of informal relationships among medium-risk females | 2.83 | (2.35, 3.15) |
| Max Infrml M LOW | Maximum number of informal relationships among low-risk males | 1.17 | (1.13, 1.21) |
| Max Infrml M MED | Maximum number of informal relationships among medium-risk males | 2.27 | (2.13, 2.46) |
| Max Mrtl F MED | Maximum number of marital relationships among medium-risk females | 1.18 | (1.12, 1.24) |
| Max Mrtl M MED | Maximum number of marital relationships among medium-risk males | 0.93 | (0.90, 0.98) |
| Max Trns F LOW | Maximum number of transitory relationships among low-risk females | 1.7 | (1.65, 1.73) |
| Max Trns F MED | Maximum number of transitory relationships among medium-risk females | 2.78 | (2.55, 3.03) |
| Max Trns M LOW | Maximum number of transitory relationships among low-risk males | 1.95 | (1.91, 2.00) |
| Max Trns M MED | Maximum number of transitory relationships among medium-risk males | 2.52 | (2.41, 2.60) |
| Mrtl Condom Max | Maximum rate of condom use in marital relationships | 0.18 | (0.18, 0.21) |
| Mrtl Condom Mid | Year midpoint of logistic scale-up of condom use in marital relationships | 2,000.76 | (2,000.28, 2,002.48) |
| Mrtl Condom Rate | Rate of logistic scale-up of condom use in marital relationships | 3.49 | (3.20, 3.61) |
| Pr Ex Infrml Fem LOW | Probability of potential for extra-relational informal relationship among low-risk females | 0.28 | (0.26, 0.32) |
| Pr Ex Infrml Fem MED | Probability of potential for extra-relational informal relationship among medium-risk females | 0.34 | (0.24, 0.37) |
| Pr Ex Infrml Male LOW | Probability of potential for extra-relational informal relationship among low-risk males | 0.49 | (0.46, 0.56) |
| Pr Ex Infrml Male MED | Probability of potential for extra-relational informal relationship among medium-risk males | 0.24 | (0.20, 0.28) |
| Pr Ex Trns Fem LOW | Probability of potential for extra-relational transitory relationship among low-risk females | 0.098 | (0.086, 0.118) |
| Pr Ex Trns Fem MED | Probability of potential for extra-relational transitory relationship among medium-risk females | 0.72 | (0.66, 0.78) |
| Pr Ex Trns Male LOW | Probability of potential for extra-relational transitory relationship among low-risk males | 0.2 | (0.14, 0.23) |
| Pr Ex Trns Male MED | Probability of potential for extra-relational transitory relationship among medium-risk males | 0.69 | (0.68, 0.75) |
| Risk Assortivity | Risk assortivity | 0.62 | (0.61, 0.65) |
| SeedYr HIGH | Seed year | 1,976.87 | (1,975.63, 1,977.77) |
| Trans Form Rate | Transitory relationship formation rate | 0.0003 | (0.0001, 0.0004) |
| Trns Condom Mid | Year midpoint of logistic scale-up of condom use in transitory relationships | 2,001.80 | (1,999.58, 2,003.89) |
| Trns Condom Rate | Rate of logistic scale-up of condom use in transitory relationships | 3.23 | (2.88, 3.48) |
| Trns Condoms Max | Maximum rate of condom use in transitory relationships | 0.22 | (0.19, 0.25) |

* Median and interquartile ranges (IQRs) reported for all dynamic parameters used in the calibration process from 100 best-fitting parameter sets. †

##### Table S8. Utility weights for estimating disability-adjusted life-years averted

| Health State | DALY Weight | Reference |
| --- | --- | --- |
| HIV-negative | 0 | Vos *et al (28)* |
| HIV and not on ART | 0.274 |  |
| HIV and on ART | 0.078 |  |

#### Costing parameters

##### Table S9 Kenya costing parameter calculations

| **Cost parameter** | **Estimate (USD)** | **Year** | **Data source** | **Calculation detials, notes** |
| --- | --- | --- | --- | --- |
| **HIV costs** |  |  |  |  |
| Annual health care costs (among those not on ART) |  |  |  |  |
| HIV-positive CD4 < 200 | 110.3 | 2021 | Eaton 2014 (29) | Adjusted for inflation and GDP/capita ratio using this approach: **Step 1:** Adjust South Africa value in 2012 USD for inflation to be in 2021 USD ($374.08 = $167*2.24)  Cost of health care use, CD4 count <200 cells per μL, not in HIV care (per person-year) in South Africa= $167 USD Inflation Rate between time of costing (2012) and 2021: 2.24 =4.7/2.1 **Step 2:** Adjust South Africa 2021 USD value by multiplying by the Kenya GDP/cap ratio ($374.08*0.295) South Africa 2021 GDP per capita in $USD = 7,055; Kenya $USD = 2,082 Kenya GDP/ ZA GDP ratio adjustment: (2,082/7,055)= 0.295 |
| HIV-positive CD4 200 - 349 | 30.38 | 2021 | Eaton 2014 (29) | Adjusted for inflation and GDP/capita ratio (see steps above) |
| HIV-positive CD4 > 350 | 8.59 | 2021 | Eaton 2014 (29) | Adjusted for inflation and GDP/capita ratio (see steps above) |
| End of life care | 105.68 | 2021 | Eaton 2014 (29) | Adjusted for inflation and GDP/capita ratio (see steps above) |
| Annual ART provision costs (30) | 196.85 | 2020 | Long et al. (2010) (30) | 1st line ART delivery cost: $121 in 2016 USD → $131 in 2020 USD includes labs + staff encounters Cost of 1st line ART: $131 USD (delivery cost in 2020 USD) + $43.20 (ART)*1.2(additional 20% suplly chain)=$ 182.84; The delivery cost ratio between 2nd and 1st ART is 2.4;  Cost of 2nd line ART: $131*2.4 USD (delivery cost in 2020 USD) + $279.60*1.2=$649.92; weighted average of 1st adn 2nd lines ART cost assuming 3% on 2nd line ART: $182.84*0.97 + $649.92*0.03=$196.85 |
| **Oral PrEP costs** |  |  |  |  |
| Oral PrEP per person month, facility | 10.88 | 2019 | Wanga et al. 2019 (31) | Data from micro-costing study, including viable (personnel, drug, lab and HIV testing, and other) and fixed (traning, demand creation, personnel, capital, overhead) costs |
| Facility-based HIV-positive test | 3.68 | 2017 | Meisner et al. (2021) (32) | Data from micro-costing study. Inputs include screening test kit, other supply costs, and personnel costs. |
| Facility-based HIV-negative test | 2.64 | 2017 | Meisner et al. (2021) (32) | Data from micro-costing study. Inputs include screening test kit, other supply costs, and personnel costs. |
| **Lenacapavir costs** |  |  |  |  |
| Demand generation | 10% |  |  | Assumption |
| Delivery | $8.55: $2.50 for HIV test, $0.05 for syringe, $6 for delivery overhead (facilities, staff, etc.) |  | Galactionova et al. (2015) (33)  Ojal et al. (2019) (34)  Mvundura et al. (2015) (35)  Mangale et al. (2022) (36)  UNICEF (2024) (37) | Syringe estimates were derived from UNICEF vaccine program estimates. Delivery overhead estimates were derived from the oral PrEP literature; provider time or overhead that is above and beyond resources needed to dispense oral PrEP are not included. |
| Product wastage | 5% of the product goes to waste |  | Ojal et al. (2019) (34) |  |

##### Table S10 South Africa costing parameter calculations

| **South Africa** |  |  |  |  |
| --- | --- | --- | --- | --- |
| **Cost parameter** | **Estimate (USD)** | **Year** | **Data source** | **Calculation detials, notes** |
| **HIV costs** |  |  |  |  |
| Annual health care costs  (among those not on ART) |  |  |  |  |
| HIV-positive CD4 < 200 | 374.08 | 2021 | Eaton 2014 (29) | Adjusted for inflation and GDP/capita ratio using this approach: Adjust South Africa value in 2012 USD for inflation to be in 2021 USD ($374.08 = $167*2.24)  Cost of health care use, CD4 count <200 cells per μL, not in HIV care (per person-year) in South Africa= $167 USD Inflation Rate between time of costing (2012) and 2021: 2.24 =4.7/2.1 |
| HIV-positive CD4 200 - 349 | 102.95 | 2021 | Eaton 2014 (29) | Adjusted for inflation and GDP/capita ratio (see above) |
| HIV-positive CD4 > 350 | 29.10 | 2021 | Eaton 2014 (29) | Adjusted for inflation and GDP/capita ratio (see above) |
| End of life care | 358.10 | 2021 | Eaton 2014 (29) | Adjust for inflation using World Bank CPI: the cost in 2021 USD for SA $160*4.7/2.1=$358.1 2021 USD. |
| Annual ART provision costs | 189.56 | 2020 | Long et al. (2010) (30) | 1st line ART delivery cost: $ 119 (including personnel, building, equipment, etc) in 2018 USD → $124 in 2020 USD  Cost of 1st line ART: $124 USD (delivery cost in 2020 USD) + $43.20 (ART)*1.2(additional 20% supply chain)=$ 175.84; The delivery cost ratio between 2nd and 1st ART is 2.4;  Cost of 2nd line ART: $124*2.4 USD (delivery cost in 2020 USD) + $279.60*1.2=$633.12 per year; weighted average of 1st and 2nd lines ART cost assuming 3% on 2nd line ART: $175.84*0.97 + $633.12*0.03=$189.56 |
| **Oral PrEP costs** |  |  |  |  |
| Oral PrEP per person month, facility | 15.20 | 2021 | Jamieson et al. (2022) (38)  Jamieson et al. (2020) (38) | Ingredients-based approach, including rapid HIV testing, counselling, provision of condoms, syndromic screening with treatment referral, adherence counselling, traning, outreach, mobilization, monitoring and evaluation costs; Average cost of oral PrEP per person initiated:$76-78 across FSW, AGYW, and heterosexual men with a duration of 5 months use. This translates to a monthly cost of ~76/5=$15.20 |
| Facility-based HIV-positive test | 5.62 | 2018 | Meyer-Rath et al. (2019) (39) | Including supply, furniture, and staff salaries |
| Facility-based HIV-negative test | 3.62 | 2018 | Meyer-Rath et al. (2019) (39) | Including supply, furniture, and staff salaries |

##### Table S11 Zimbabwe costing parameter calculations

|  | **Kenya** |  |  |  |
| --- | --- | --- | --- | --- |
| **Cost parameter** | **Estimate** | **Year** | **Data source** | **Calculation details, notes** |
| **HIV costs** |  |  |  |  |
| Annual health care costs  (among those not on ART) |  |  |  |  |
| HIV-positive CD4 < 200 | 93.98 | 2021 | Eaton et al. (2014) (29) | Adjusted for inflation and GDP/capita ratio using this approach: **Step 1:** Adjust South Africa value in 2012 USD for inflation to be in 2021 USD ($374.08 = $167*2.24)  Cost of health care use, CD4 count <200 cells per μL, not in HIV care (per person-year) in South Africa= $167 USD Inflation Rate between time of costing (2012) and 2021: 2.24 =4.7/2.1 **Step 2:** Adjust South Africa 2021 USD value by multiplying by the Zimbabwe GDP/cap ratio ($374.08*0.251) South Africa 2021 GDP per capita in $USD = 7,055; ZA $USD = 1,774 Kenya GDP/ ZA GDP ratio adjustment: (1,774/7,055)= 0.251 |
| HIV-positive CD4 200 - 349 | 25.89 | 2021 | Eaton et al. (2014) (29) | Adjusted for inflation and GDP/capita ratio (see steps above) |
| HIV-positive CD4 > 350 | 7.32 | 2021 | Eaton et al. (2014) (29) | Adjusted for inflation and GDP/capita ratio (see steps above) |
| End of life care | 89.83 | 2021 | Eaton et al. (2014) (29) | Adjusted for inflation and GDP/capita ratio (see steps above) |
| Annual ART provision costs | 176.01 | 2020 | Long et al. (2010) (30) | 1st line ART delivery cost, use ratio of Kenya:Zim GDPs (1774/2082=0.85): $131*0.85 =$111 in 2020 USD includes labs + staff encounters Cost of 1st line ART: $111 USD (delivery cost in 2020 USD) + $43.20 (ART)*1.2(additional 20% suplly chain)=$ 162.84; The delivery cost ratio between 2nd and 1st ART is 2.4;  Cost of 2nd line ART: $111*2.4 USD (delivery cost in 2020 USD) + $279.60*1.2=$601.92; weighted average of 1st adn 2nd lines ART cost assuming 3% on 2nd line ART: $162.84*0.97 + $601.92*0.03=$176.01 |
| **Oral PrEP costs** |  |  |  |  |
| Oral PrEP per person month, facility | 9.25 | 2019 | Wanga et al. (2019) (31) | Translated from the cost from Kenya using ratio of Kenya:Zim GDPs (1774/2082=0.85) |
| Facility-based HIV-positive test | 3.13 | 2017 | Meisner et al. 2021 (32) | same with above |
| Facility-based HIV-negative test | 2.24 | 2017 | Meisner et al. 2021 (32) | same with above |

#### Number of Oral PrEP initiations in Western Kenya, South Africa, and Zimbabwe

##### Table S12 Number of PrEP initiations in Western Kenya, South Africa, and Zimbabwe

|  | Western Kenya | South Africa | Zimbabwe |
| --- | --- | --- | --- |
| 2016 | 655.2 | 722 | 288 |
| 2017 | 4376.4 | 3362 | 2677 |
| 2018 | 13208.4 | 8476 | 6131 |
| 2019 | 3370.8 | 45576 | 9302 |
| 2020 | 11543.6 | 106402 | 9501 |
| 2021 | 18007.2 | 205657 | 32019 |
| 2022 | 68220.4 | 422239 | 83580 |

Source: Global PrEP tracker, AVAC (40,41)

### VOICE Risk Score

The VOICE risk score is an empirical HIV risk score derived from the VOICE trial to predict HIV acquisition risk in SSA with a higher score indicating a higher risk of HIV acquisition (42). The VOICE score can be calculated based on summing the point values that correspond to the following factors: age<25 years (2 points), unmarried or not living with a primary partner(2 points), partner does not provide financial or material support (1 point); primary partner has other partners (yes or don’t know, 2 points); alcohol use in the past 3 months (1 point); having a curable sexually transmitted infection (1 point); herpes simplex virus type 2 (HSV-2) status (2 points). Due to several of these factors are not available in EMOD, we adapted the VOICE score calculation and only took the sum of the point values for the following factors: younger age (i.e., 15-24, 2 points), unmarried status (2 points), ≥ 1 male sexual partner who has other partners (2 points) and being identified as median sexual risk by EMOD (2 points). The modeled population in EMOD is stratified into three levels of sexual risk behavior groups: high-risk group representing sex workers and clients, medium risk group with short-term partnerships and increased propensity to form multiple concurrent partnerships, and low-risk group with fewer, lower-term partnerships (43). The median sexual risk category in EMOD was used as a proxy for other risk factors included in the original VOICE score calculation but not available in EMOD.
